# Development and validation of treatment-decision algorithms for children evaluated for pulmonary tuberculosis: an individual participant data meta-analysis

**DOI:** 10.1101/2022.09.13.22279911

**Authors:** Kenneth S. Gunasekera, Olivier Marcy, Johanna Muñoz, Elisa Lopez-Varela, Moorine P. Sekadde, Molly F. Franke, Maryline Bonnet, Shakil Ahmed, Farhana Amanullah, Aliya Anwar, Orvalho Augusto, Rafaela Baroni Aurilio, Sayera Banu, Iraj Batool, Annemieke Brands, Kevin P. Cain, Lucía Carratalá-Castro, Maxine Caws, Eleanor S. Click, Lisa M. Cranmer, Alberto L. García-Basteiro, Anneke C. Hesseling, Julie Huynh, Senjuti Kabir, Leonid Lecca, Anna Mandalakas, Farai Mavhunga, Aye Aye Myint, Kyaw Myo, Dorah Nampijja, Mark P. Nicol, Patrick Orikiriza, Megan Palmer, Clemax Couto Sant’Anna, Sara Ahmed Siddiqui, Jonathan P. Smith, Rinn Song, Nguyen Thuy Thuong Thuong, Vibol Ung, Marieke M. van der Zalm, Sabine Verkuijl, Kerri Viney, Elisabetta G. Walters, Joshua L. Warren, Heather J. Zar, Ben J. Marais, Stephen M. Graham, Thomas P. A. Debray, Ted Cohen, James A. Seddon

**Affiliations:** Department of Epidemiology of Microbial Diseases, Yale School of Public Health, New Haven, USA; University of Bordeaux, Inserm UMR1219, Institut de Recherche pour le Développement (IRD) EMR 271, Bordeaux France; Julius Center for Health Sciences and Primary Care, University Medical Center Utrecht, Utrecht University, Utrecht, The Netherlands; Barcelona Institute for Global Health (ISGlobal), Hospital Clínic - Universitat de Barcelona, Barcelona, Spain; National Tuberculosis and Leprosy Program, Kampala, Uganda; Department of Global Health and Social Medicine, Harvard Medical School, Boston, USA; University of Montpellier, Inserm, Institut de Recherche pour le Développement, TransVIHMI, Montpellier, France; Epicentre, Mbarara, Uganda; Department of Paediatrics, Dhaka Medical College Hospital, Dhaka, Bangladesh; Indus Hospital & Health Network, Karachi, Pakistan; The Aga Khan University Hospital, Karachi, Pakistan; Centro de Investigação em Saúde de Manhiça, Maputo, Mozambique; Instituto de Puericultura e Pediatria Martagao Gesteira. Universidade Federal do Rio de Janeiro, Rio de Janeiro, Brazil; Programme on Emerging Infections, Infectious Disease Division, icddr,b, Dhaka, Bangladesh; Global Tuberculosis Programme, World Health Organization, Geneva, Switzerland; US Centers for Disease Control and Prevention, Atlanta, USA; Manhiça Health Research Centre, Maputo, Mozambique; Barcelona Institute for Global Health, Barcelona, Spain; Liverpool School of Tropical Medicine, Liverpool, UK; Emory School of Medicine, Atlanta, USA; Emory Rollins School of Public Health, Atlanta, USA; Children’s Healthcare of Atlanta, Atlanta, USA; Centro de Investigação em Saúde de Manhiça, CISM,e; Maputo, Mozambique; ISGlobal, Hospital Clínic - Universitat de Barcelona, Barcelona, Spain; Centro de Investigación Biomédica en Red de Enfermedades Infecciosas (CIBERINFEC), Barcelona, Spain; Desmond Tutu TB Centre, Department of Paediatrics and Child Health, Faculty of Medicine and Health Sciences, Stellenbosch University, Tygerberg, South Africa; Oxford University Clinical Research Unit, Centre for Tropical Diseases, Ho Chi Minh City, Vietnam; Nuffield Department of Medicine, Oxford University, UK; Socios En Salud Surcursal Perú, Lima, Perú; Global TB Program, Baylor College of Medicine and Texas Children’s Hospital, Houston, USA; Clinical Infectious Disease Group, German Center for Infectious Research (DZIF), Clinical TB Unit, Research Center Borstel, Borstel, Germany; Department of Paediatrics, University of Medicine, Mandalay, Myanmar; Department of Paediatrics, University of Medicine, Magway, Myanmar; Mbarara University of Science and Technology, Mbarara, Uganda; Division of Infection and Immunity, Department of Biomedical Sciences, University of Western Australia, Perth, Australia; Department of Microbiology, Division of Basic Medical Sciences, School of Medicine, University of Global Health Equity, Kigali, Rwanda; Faculty of Medicine, Universidade Federal do Rio de Janeiro, Rio de Janeiro, Brazil; Department of Health Policy and Management, Yale School of Public Health, New Haven, USA; Oxford Vaccine Group, Department of Paediatrics, University of Oxford, Oxford, UK; Division of Infectious Diseases, Boston Children’s Hospital, Boston, Massachusetts; Department of Pediatrics, Harvard Medical School, Boston, USA; Oxford University Clinical Research Unit, Ho Chi Minh City, Vietnam; Nuffield Department of Medicine, Centre for TropicalMedicine and Global Health, University of Oxford, Oxford, UK; University of Health Sciences, Phnom Penh, Cambodia; National Pediatric Hospital, Phnom Penh, Cambodia; School of Public Health, University of Sydney, Sydney, Australia; Directorate of Integrated Laboratory Medicine, Institute of Human Genetics, Newcastle upon Tyne NHS Foundation Trust, Newcastle upon Tyne, United Kingdom; Department of Biostatistics, Yale School of Public Health, New Haven, USA; Department of Paediatrics & Child Health, Red Cross Children’s Hospital, Cape Town, South Africa; SA-MRC Unit on Child & Adolescent Health, University of Cape Town, South Africa; The Children’s Hospital at Westmead Clinical School, Faculty of Medicine and Health, University of Sydney, Sydney, Australia; University of Melbourne Department of Paediatrics and Murdoch Children’s Research Institute, Royal Children’s Hospital, Melbourne, Australia; Burnet Institute, Melbourne, Australia; Desmond Tutu Tuberculosis Centre, Department of Paediatrics and Child Health, Faculty of Medicine and Health Sciences, Stellenbosch University, Tygerberg, South Africa; Department of Infectious Diseases, Imperial College London, London, UK

**Keywords:** Diagnostic algorithm, diagnostic system, tuberculosis, paediatric, clinical evidence, Diagnosis/* clinical decision-making

## Abstract

**Background:** Many children with pulmonary tuberculosis remain undiagnosed and untreated with related high morbidity and mortality. Diagnostic challenges in children include low bacterial burden, challenges around specimen collection, and limited access to diagnostic expertise. Algorithms that guide decisions to initiate tuberculosis treatment in resource-limited settings could help to close the persistent childhood tuberculosis treatment gap. Recent advances in childhood tuberculosis algorithm development have incorporated prediction modelling, but studies conducted to date have been small and localised, with limited generalizability.

**Methods:** We collated individual participant data including clinical, bacteriological, and radiologic information from prospective diagnostic studies in high-tuberculosis incidence settings enrolling children <10 years with presumptive pulmonary tuberculosis. Using this dataset, we first retrospectively evaluated the performance of several existing treatment-decision algorithms and then developed multivariable prediction models, investigating model generalisability using internal-external cross-validation. A team of experts provided input to adapt the models into a pragmatic treatment-decision algorithm with a pre-determined sensitivity threshold of 85% for use in resource-limited, primary healthcare settings.

**Findings:** Of 4,718 children from 13 studies from 12 countries, 1,811 (38·4%) were classified as having pulmonary tuberculosis; 541 (29·9%) bacteriologically confirmed and 1,270 (70·1%) unconfirmed. Existing treatment-decision algorithms had highly variable diagnostic performance. Our prediction model had a combined sensitivity of 86% [95% confidence interval (CI): 0·68-0·94] and specificity of 37% [95% CI: 0·15-0·66] against a composite reference standard.

**Interpretation:** We adopted an evidence-based approach to develop pragmatic algorithms to guide tuberculosis treatment decisions in children, irrespective of the resources locally available. This approach will empower health workers in resource-limited, primary healthcare settings to initiate tuberculosis treatment in children in order to improve access to care and reduce tuberculosis-related mortality. These algorithms have been included in the operational handbook accompanying the latest WHO guidelines on the management of tuberculosis in children and adolescents.

**Funding:** World Health Organization, US National Institutes of Health

**RESEARCH IN CONTEXT:** *Evidence before the study:* Treatment-decision algorithms relate information gained in the evaluation of children into an assessment of tuberculosis disease risk and empower healthcare workers to make appropriate treatment decisions. Studies in primary healthcare centres have demonstrated that use of treatment-decision algorithms can improve childhood pulmonary tuberculosis case-detection and treatment initiation in settings with high-tuberculosis incidence. To identify primary research studies on treatment-decision algorithm performance evaluation and/or development for childhood pulmonary tuberculosis, we carried out a PubMed search using the terms (‘child*’ OR ‘paediatr*’ OR ‘pediatr*’) AND (‘tuberculosis’ OR ‘TB’) AND (‘treatment-decision’ OR ‘algorithm’ OR ‘diagnos*’) to identify primary research published in any language prior to 29 June 2022. We additionally consulted multiple experts in childhood pulmonary tuberculosis diagnosis and management, and we referred to existing, published reviews of treatment-decision algorithms. With respect to treatment-decision algorithm performance, several studies have retrospectively estimated the performance of treatment-decision algorithms in a single geographic setting; a subset of these studies have also compared the performance of multiple algorithms using data from a single geographic setting. With respect to treatment-decision algorithm development, many existing algorithms have been developed without explicit analysis of data from children with presumptive pulmonary tuberculosis, often developed from expert consensus. Gunasekera et al. used model-based approaches to analyse diagnostic evaluations data (e.g., clinical history, physical examination, chest radiograph, and results from rapid molecular and culture testing for *Mycobacterium tuberculosis*) collected from children with presumptive pulmonary tuberculosis in a single geographic setting to inform the development of a diagnostic algorithm while Marcy et al. and Fourie et al analysed data from multiple geographic settings. However, these studies were relatively small with limited assessment of generalisability.

*Added value of this study:* We collated individual participant data from 13 prospective diagnostic studies from 12 countries including 4,718 children with presumptive pulmonary tuberculosis from geographically diverse settings with a high incidence of tuberculosis in order to 1) evaluate the performance of existing treatment-decision algorithms and 2) develop multivariable logistic regression models to quantify the contribution of individual features to discriminate tuberculosis from non-tuberculosis. A panel of child tuberculosis experts provided input into performance targets and advised on how to incorporate scores derived from these models into pragmatic treatment-decision algorithms to assist in the evaluation of children presenting with presumptive pulmonary tuberculosis in primary healthcare centres.

*Implications of all the available evidence:* Our findings suggest that evidence-based, pragmatic treatment-decision algorithms can be developed to make sensitive and clinically appropriate decisions to treat a child with pulmonary tuberculosis. Although the specificity does not reach optimal targets for childhood tuberculosis diagnosis, pragmatic treatment-decision algorithms provide clinically relevant guidance that can empower health workers to start children on tuberculosis treatment at the primary healthcare setting and will likely contribute to reducing the case-detection gap in childhood tuberculosis. External, prospective evaluation of these novel algorithms in diverse settings is required, including assessment of their accuracy, feasibility, acceptability, impact, and cost-effectiveness. This work led to a new interim WHO recommendation to support the use of treatment-decision algorithms in the evaluation of children with presumptive tuberculosis in the 2022 updated consolidated guidelines on the management of tuberculosis in children. Two algorithms developed from this work have been included in the WHO operational handbook accompanying these guidelines.

## INTRODUCTION

Tuberculosis is a leading cause of mortality among children worldwide,^1^ accounting for ∼2·5% of the 6 million deaths in children <5 years each year.^2^ Modelling suggests that more than 96% of tuberculosis deaths in children and adolescents (<15 years) occurred in those not receiving tuberculosis treatment.^3^ The World Health Organization (WHO) estimates that fewer than 50% of the 1·1 million children <15 years who develop tuberculosis are diagnosed; the proportion is even lower among children <5 years, at about 27%.^1^ Thus, efforts to improve diagnosis, and thereby improve access to tuberculosis treatment, are an important opportunity to reduce tuberculosis morbidity and deaths among children.

Diagnosing pulmonary tuberculosis among children is challenging as respiratory specimens tend to be paucibacillary, resulting in a low yield of bacteriologic confirmation.^4^ Furthermore, collecting respiratory specimens from young children is invasive and requires resources that are generally concentrated in higher-level healthcare centres. Thus, careful symptom review, clinical examination, chest radiography, and history of *Mycobacterium tuberculosis* (*Mtb*) exposure can inform treatment decisions in clinical care. However, paediatric clinical expertise and resources to make a diagnosis are often limited at primary healthcare centres. This limits treatment access and leads to either delays in treatment initiation or no treatment initiation which are associated with worse outcomes, including mortality.^5,6^ Facilitating appropriate diagnostic assessment with rapid treatment initiation at healthcare settings where children initially present could contribute to reductions in tuberculosis-related morbidity and mortality.

Treatment-decision algorithms aim to standardise clinical assessment and decision-making. Algorithms relate information gained in the evaluation of children into an assessment of tuberculosis disease risk and empower healthcare workers to make appropriate treatment decisions. Adopting an algorithmic approach to treatment decision-making has been shown to improve childhood tuberculosis case detection and treatment access at primary healthcare settings.^7,8^ However, these algorithms were developed using consensus expert opinion rather than analysis of data.

Recent approaches for algorithm generation have used data from cross-sectional childhood tuberculosis diagnostic studies to quantify the contribution of clinical characteristics to the risk of tuberculosis disease.^9-11^ Evidence-based approaches are objective and offer the potential for validation; however, existing studies have been small and not generalisable. In this study, we assembled individual participant data (IPD) from children investigated for presumptive pulmonary tuberculosis. We then sought to first evaluate the performance of currently used diagnostic algorithms and then develop evidence-based treatment-decision algorithms. This work was conducted to inform the 2022 WHO guidelines for the management of tuberculosis in children and adolescents and the accompanying WHO operational handbook.^12,13^

## METHODS

### Establishment of individual participant data

We identified potential sources of IPD through responses to a public WHO Global Tuberculosis Programme Public Call for Data on the Management of Children with Tuberculosis in July 2020 and through referral from paediatric tuberculosis experts. Studies were eligible for inclusion if they 1) prospectively recruited consecutive participants <10 years (the definition of a child in the 2022 WHO guideline) attending healthcare centres in high-tuberculosis incidence countries for clinical evaluation of pulmonary tuberculosis and 2) provided a final research classification of pulmonary tuberculosis for each child. Tuberculosis was classified using the revised US National Institutes of Health (NIH) clinical case definitions of intrathoracic tuberculosis in children.^14^ Broadly, this defines a confirmed tuberculosis case as culture- or Xpert MTB/RIF-confirmed *Mtb* from respiratory specimen(s); an unconfirmed tuberculosis case as having symptoms, chest radiography findings, and/or immune tests of *Mtb* sensitivity suggestive of tuberculosis (including follow-up to verify or rule out tuberculosis); and an unlikely tuberculosis case as meeting criteria for neither confirmed nor unconfirmed tuberculosis. If the study did not use the NIH classification, we used the study-specific definition of unconfirmed tuberculosis and unlikely tuberculosis. Quality assessment was performed using a modified version of the Newcastle-Ottawa Scale for cohort studies.^15^

After identification of eligible studies, we requested IPD including details from the clinical history, physical examination, chest radiograph, and results from rapid molecular and culture testing for *Mtb* performed on respiratory specimens collected at study entry (<u>Supplementary Appendix A)</u>. All data assembly and analysis were carried out using R software version 4.1.1. To account for the uncertainty associated with incomplete data, we used multilevel multiple imputation by chained equations (MICE) implemented in the *MICE* package to generate 100 imputed datasets (<u>Supplementary Appendix B)</u>.^16^

### *Evaluation* of *existing treatment-decision algorithms*

We identified existing treatment-decision algorithms and scores (henceforth referred to as algorithms) to guide the evaluation of children with presumptive pulmonary tuberculosis through consultation with members of the WHO Guideline Development Group (GDG) on the management of tuberculosis in children and adolescents. We defined a composite reference standard using the NIH definitions of confirmed and unconfirmed pulmonary tuberculosis to evaluate the performance of these algorithms. We carried out a sensitivity analysis of performance using a reference standard of confirmed pulmonary tuberculosis only (excluding children with unconfirmed tuberculosis). We used the “reitsma” function from the R package *mada* to pool study-level sensitivity and specificity estimates using a bivariate random effects meta-analysis (<u>Supplementary Appendix C)</u>.^17,18^

### Prediction model development and validation

We developed a multivariable logistic regression model to predict pulmonary tuberculosis using the composite reference standard in accordance with the Transparent Reporting of a Multivariable Prediction Model for Individual Prognosis or Diagnosis standards using the internal-external cross-validation framework.^19,20^ Predictors included clinical features commonly considered in the evaluation of presumptive childhood pulmonary tuberculosis in primary and secondary healthcare centres that were available in the data with <50% missingness. We also built a model without chest radiograph data to inform predictions in healthcare centres without access to radiology services.

To account for possible heterogeneity in the distribution of predictor and outcome variables, we fit the prediction model separately in each study and subsequently pooled their regression coefficients and respective standard errors. This approach was implemented in the “metapred” function of package *metamisc*.^20,21^ To account for missing data, we generated a prediction model as described above from each of the 100 imputed datasets and then used Rubin’s rules to pool the regression coefficients and standard errors to generate a final, single prediction model and compute odds ratios with 95% confidence interval (95% CI).^22^ We examined the c-statistic (also known as the area under the receiver operating characteristic curve) to assess the model’s ability to distinguish between children with tuberculosis and unlikely tuberculosis, and we examined the observed:expected (O:E) ratio to assess whether there were studies in which the model over- or under-predicted tuberculosis.

### Algorithm development

To generate clinically and programmatically implementable algorithms, we scaled the coefficient estimates for the parameters of the final prediction models (developed from all *n* studies) to estimate scores for each parameter such that a combined score of >10 corresponded to classification of tuberculosis at fixed sensitivities of 90%, 85%, 80%, 75%, and 70% (<u>Supplementary Appendix D)</u>. To estimate the sensitivity and specificity of the score in classifying tuberculosis using the composite reference standard, study-level sensitivities and specificities were pooled using the bivariate model of Reitsma et al. (implemented in the *mada* package) accounting for uncertainty introduced by imputation of missing data.^17,18^ As a sensitivity analysis, we evaluated the performance of the score against a reference standard of confirmed pulmonary tuberculosis only.

We worked with staff from the WHO Global TB Programme to identify a group of experts in childhood tuberculosis (henceforth referred to as the expert group; <u>Supplementary Appendix E</u>) to advise on the development of two treatment-decision algorithms from these scores.

Specifically, we sought advice on how to use this score within an algorithm intended to be used at primary healthcare centres and on selection of a performance target for the development of the score to be included within the algorithm.

### Ethics

This study was approved by the Stellenbosch University Health Research Ethics Committee (Ref No. X21/02/003) and the Yale Institutional Review Board (Ref No. 2000028046). All collaborating investigators confirmed institutional ethical approval for their original data collection.

## RESULTS

### Data assembly

Eighteen studies were identified as having potentially appropriate data, largely sourced from diagnostic evaluation studies. The study investigators for two studies were unable to provide data in the necessary timeline and an additional three studies did not meet the inclusion criteria. From the 13 included studies carried out in 12 countries, 4,718 IPD records from children <10 years with presumptive pulmonary tuberculosis were available, of which, 541 (11·5%) were classified as having confirmed tuberculosis, 1,270 (26·9%) as having unconfirmed tuberculosis and 2,818 (59·7%) unlikely tuberculosis (**Tables 1 and 2;** <u>Supplementary Appendices F and G</u>). The data were predominantly collected at secondary or tertiary/referral healthcare centres. Although each study was required to include children with presumptive pulmonary tuberculosis, studies differed slightly with respect to inclusion criteria, variable definitions, and reference classification of tuberculosis (<u>Supplementary Appendices H-K</u>). All contributing studies achieved quality assessment scores of 4/5 or 5/5. (<u>Supplementary Appendix L</u>).

**Table 1.**
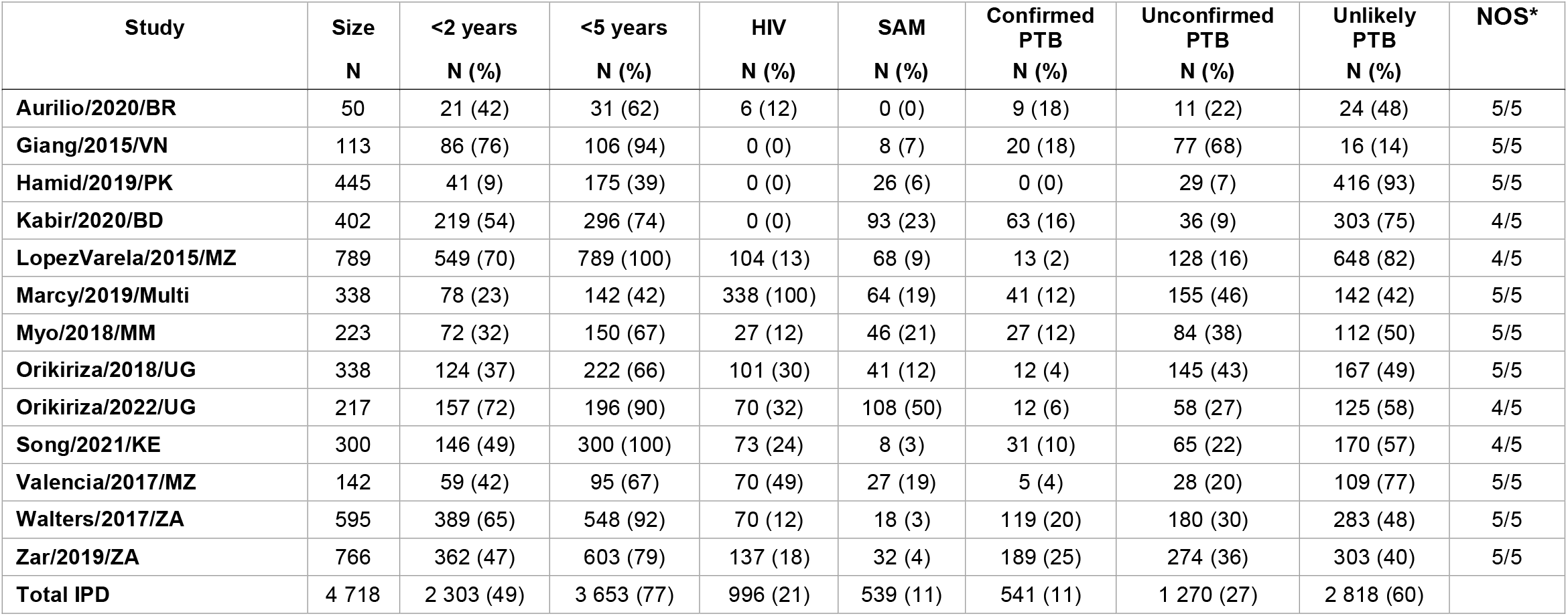
Study-level descriptions of data included in the individual participant dataset. HIV – human immunodeficiency virus, SAM – severely acutely malnourished, PTB – pulmonary tuberculosis, Refs – references, BD – Bangladesh, BR – Brazil, KE – Kenya, MM – Myanmar, Multi – Multi-country study (includes Burkina Faso, Cameroon, Vietnam, and Cambodia), MZ – Mozambique, PK – Pakistan, UG – Uganda, VN – Vietnam, ZA – South Africa. *Note: Modified version of the Newcastle-Ottawa Scale for cohort studies.

**Table 2.** Characteristics of individual participant dataset. All clinical, bacteriology, and imaging data collected from the initial evaluation. IPD – Individual participant data, IQR – interquartile range, BCG – bacille Calmette-Guerin, HIV – human immunodeficiency virus, Mtb – *Mycobacterium tuberculosis*, CXR – chest x-ray, TST – tuberculin skin test.

| Variable | Value | Confirmed tuberculosis (N=541) | Unconfirmed tuberculosis (N=1 270) | Unlikely tuberculosis (N=2 818) |
| --- | --- | --- | --- | --- |
|  |  | n (%) OR median (IQR) | n (%) OR median (IQR) | n (%) OR median (IQR) |
| Age | *Months* | 25.6 (12,58.7) | 24.1 (12,53.6) | 24.4 (13.4,54) |
|  | *Unknown* | 0 (0) | 0 (0) | 0 (0) |
| Sex | *Male* | 285 (53) | 715 (56) | 1512 (54) |
|  | *Female* | 256 (47) | 554 (44) | 1306 (46) |
|  | *Unknown* | 0 (0) | 1 (0) | 0 (0) |
| Weight-for-age | *z-score* | -1.7 (-2.9,-0.6) | -1.7 (-2.9,-0.6) | -1.8 (-3,-0.7) |
|  | *Unknown* | 7 (1) | 16 (1) | 119 (4) |
| BCG vaccination | *Evidence of BCG vaccination* | 462 (85) | 1 084 (85) | 2 105 (75) |
|  | *No evidence of BCG vaccination* | 45 (8) | 89 (7) | 152 (5) |
|  | *Unknown* | 34 (6) | 97 (8) | 561 (20) |
| HIV status | *HIV-positive* | 111 (21) | 396 (31) | 464 (16) |
|  | *HIV-negative* | 425 (79) | 841 (66) | 2 254 (80) |
|  | *Unknown* | 5 (1) | 33 (3) | 100 (4) |
| Cough duration | *No cough* | 75 (14) | 205 (16) | 823 (29) |
|  | *Cough 0-13 days* | 148 (27) | 279 (22) | 598 (21) |
|  | *Cough 14-20 days* | 90 (17) | 140 (11) | 321 (11) |
|  | *Cough 21-27 days* | 52 (10) | 96 (8) | 224 (8) |
|  | *Cough >27 days* | 125 (23) | 260 (20) | 512 (18) |
|  | *Unknown* | 51 (9) | 290 (23) | 340 (12) |
| Cough ≥ 2 weeks | *Cough ≥2 weeks present* | 295 (55) | 697 (55) | 1 247 (44) |
|  | *Cough ≥2 weeks not present* | 238 (44) | 562 (44) | 1531 (54) |
|  | *Unknown* | 8 (1) | 11 (1) | 40 (1) |
| Fever duration | *No fever* | 146 (27) | 352 (28) | 1 157 (41) |
|  | *Fever 0-13 days* | 136 (25) | 281 (22) | 712 (25) |
|  | *Fever 14-20 days* | 67 (12) | 122 (10) | 262 (9) |
|  | *Fever 21-27 days* | 20 (4) | 40 (3) | 88 (3) |
|  | *Fever >27 days* | 77 (14) | 119 (9) | 184 (7) |
|  | *Unknown* | 95 (18) | 356 (28) | 415 (15) |
| Fever ≥1 week | *Fever ≥1 week present* | 209 (39) | 388 (31) | 779 (28) |
|  | *Fever ≥1 week not present* | 242 (45) | 650 (51) | 1 776 (63) |
|  | *Unknown* | 90 (17) | 232 (18) | 263 (9) |
| Lethargy | *Lethargy* | 268 (50) | 479 (38) | 838 (30) |
|  | *No lethargy* | 226 (42) | 565 (44) | 1 204 (43) |
|  | *Unknown* | 47 (9) | 226 (18) | 776 (28) |
| Weight loss | *Weight loss* | 358 (66) | 763 (60) | 1 631 (58) |
|  | *No weight loss* | 172 (32) | 492 (39) | 1 155 (41) |
|  | *Unknown* | 11 (2) | 15 (1) | 32 (1) |
| Documented tuberculosis exposure | *Documented tuberculosis exposure in previous 12 months* | 257 (48) | 491 (39) | 497 (18) |
|  | *No documented tuberculosis exposure in previous 12 months* | 284 (52) | 775 (61) | 2315 (82) |
|  | *Unknown* | 0 (0) | 4 (0) | 6 (0) |
| Night sweats | *Night sweats* | 206 (38) | 422 (33) | 568 (20) |
|  | *No night sweats* | 187 (35) | 577 (45) | 1 845 (65) |
|  | *Unknown* | 148 (27) | 271 (21) | 405 (14) |
| Haemoptysis | *Haemoptysis* | 2 (0) | 13 (1) | 12 (0) |
|  | *No haemoptysis* | 88 (16) | 410 (32) | 851 (30) |
|  | *Unknown* | 451 (83) | 847 (67) | 1 955 (69) |
| Objective fever (≥38 degrees Celsius) | *Objective fever* | 71 (13) | 101 (8) | 156 (6) |
|  | *No objective fever* | 228 (42) | 738 (58) | 1 476 (52) |
|  | *Unknown* | 242 (45) | 431 (34) | 1 186 (42) |
| Tachycardia | *Tachycardia* | 80 (15) | 150 (12) | 175 (6) |
|  | *No tachycardia* | 179 (33) | 611 (48) | 1 187 (42) |
|  | *Unknown* | 282 (52) | 509 (40) | 1 456 (52) |
| **Tachypnoea** | *Tachypnoea* | 128 (24) | 264 (21) | 320 (11) |
|  | *No tachypnoea* | 276 (51) | 742 (58) | 1 494 (53) |
|  | *Unknown* | 137 (25) | 264 (21) | 1 004 (36) |
| **Peripheral lymphadenopathy** | *Peripheral lymphadenopathy* | 134 (25) | 242 (19) | 280 (10) |
|  | *No peripheral lymphadenopathy* | 295 (55) | 759 (60) | 1 444 (51) |
|  | *Unknown* | 112 (21) | 269 (21) | 1 094 (39) |
| **First Xpert MTB/RIF** | *Xpert positive for Mtb* | 247 (46) | 0 (0) | 0 (0) |
|  | *Xpert negative for Mtb* | 273 (50) | 1 148 (90) | 2 306 (82) |
|  | *Unknown/not performed* | 21 (4) | 122 (10) | 512 (18) |
| **Overall CXR read** | *CXR consistent with tuberculosis* | 344 (64) | 644 (51) | 653 (23) |
|  | *CXR not consistent with tuberculosis* | 101 (19) | 474 (37) | 1 744 (62) |
|  | *Unknown/not assessed* | 96 (18) | 152 (12) | 421 (15) |
| **Opacities on CXR** | *Opacities present on CXR* | 237 (44) | 487 (38) | 594 (21) |
|  | *Opacities not present on CXR* | 154 (28) | 459 (36) | 1 042 (37) |
|  | *Unknown/not assessed* | 150 (28) | 324 (26) | 1 182 (42) |
| **Cavities on CXR** | *Cavities present on CXR* | 32 (6) | 25 (2) | 20 (1) |
|  | *Cavities not present on CXR* | 348 (64) | 793 (62) | 973 (35) |
|  | *Unknown/not assessed* | 161 (30) | 452 (36) | 1 825 (65) |
| **Miliary infiltrate on CXR** | *Miliary infiltrate present on CXR* | 33 (6) | 19 (1) | 7 (0) |
|  | *Miliary infiltrate not present on CXR* | 358 (66) | 927 (73) | 1 614 (57) |
|  | *Unknown/not assessed* | 150 (28) | 324 (26) | 1 197 (42) |
| **Intrathoracic lymphadenopathy on CXR** | *Intrathoracic lymphadenopathy present on CXR* | 154 (28) | 248 (20) | 104 (4) |
|  | *Intrathoracic lymphadenopathy not present on CXR* | 237 (44) | 684 (54) | 1 485 (53) |
|  | *Unknown/not assessed* | 150 (28) | 338 (27) | 1 229 (44) |
| **Pleural effusion on CXR** | *Pleural effusion present on CXR* | 56 (10) | 63 (5) | 49 (2) |
|  | *Pleural effusion not present on CXR* | 335 (62) | 883 (70) | 1 570 (56) |
|  | *Unknown/not assessed* | 150 (28) | 324 (26) | 1 199 (43) |
| **Tuberculin skin test** | *TST positive* | 298 (55) | 464 (37) | 262 (9) |
|  | *TST negative* | 174 (32) | 674 (53) | 1 972 (70) |
|  | *Unknown/not performed* | 69 (13) | 132 (10) | 584 (21) |

### Existing treatment-decision algorithm performance

We evaluated the performance of eight existing treatment-decision algorithms. One of these algorithms was evaluated only on data from children living with HIV, and another evaluated only on data from children without HIV. Because some algorithms considered evidence that were not available in the IPD, we modified all algorithms slightly (<u>Supplementary Appendix M</u>). The sensitivities varied from 17% to 93% with specificities varying from 88% to 16% when evaluated against the composite reference standard (**Figure 1**; <u>Supplementary Appendix N)</u>. A sensitivity analysis evaluating performance to discriminate confirmed tuberculosis from unlikely tuberculosis demonstrated marginally higher sensitivities and comparable specificities to the performance in the entire dataset (<u>Supplementary Appendix O</u>).

**Figure 1.**
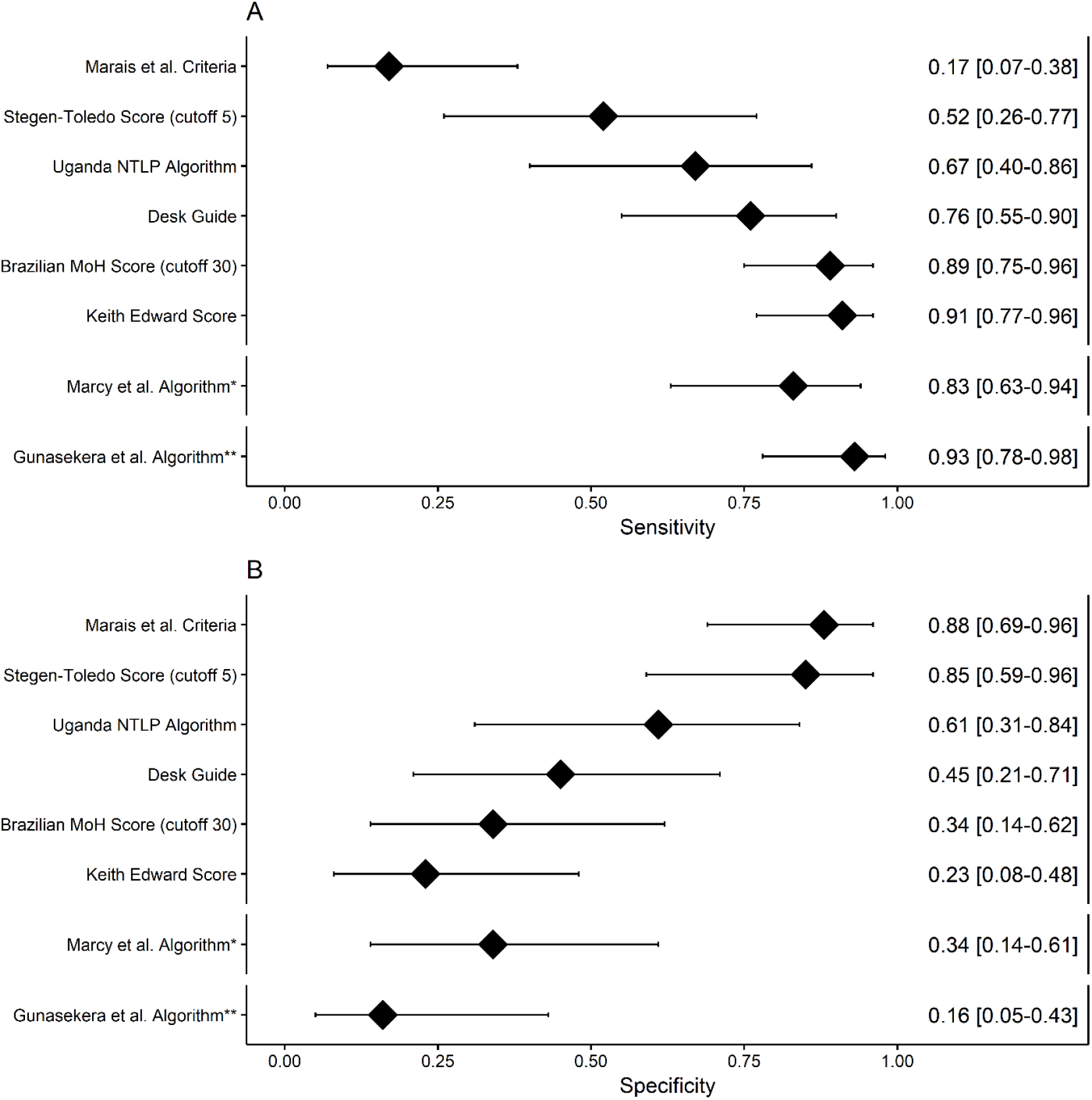
Performance of existing treatment-decision algorithms at classifying tuberculosis. Retrospective estimates of the pooled **(A)** sensitivity and **(B)** specificity of eight algorithms to guide decisions to treat children with presumptive pulmonary tuberculosis, had they been used to evaluate the children for whom we have IPD records. The reference classification of pulmonary tuberculosis included bacteriologically-confirmed pulmonary tuberculosis as well as unconfirmed pulmonary tuberculosis. Modifications were made to the algorithms to maximise the use of the available IPD. IPD – individual participant data, HIV – human immunodeficiency virus, BD – Bangladesh, BR – Brazil, KE – Kenya, MM – Myanmar, Multi – (PAANTHER) Multi-country study (includes Burkina Faso, Cameroon, Vietnam, and Cambodia), MZ – Mozambique, PK – Pakistan, UG – Uganda, VN – Vietnam, ZA – South Africa, MoH – (Brazil) Ministry of Health, NTLP – (Uganda) National TB and Leprosy Program. *Performance estimates of the Marcy et al. Algorithm were derived from only HIV-positive children in the IPD that excludes data form the Marcy/2016/Multi cohort (from which the algorithm was developed), **Performance estimates of the Gunasekera et al. Algorithm were derived from only HIV-negative children in the IPD that excludes data from the Walter/2017/ZA population (from which the algorithm was developed).

### Prediction model development and validation

Odds ratios and 95% CI of the predictors included in the model are displayed in **Table 3**. The pooled c-statistic for the prediction model including chest x-ray features was 0·71 (95% CI: 0·66-0·76), with a summary O:E ratio of 0·90 (95% CI: 0·28-2·98). Additional internal-external cross-validation c-statistic and O:E ratio estimates are included in <u>Supplementary Appendix P</u>. Estimates for the model without chest x-ray features are included in <u>Supplementary Appendix Q</u>.

**Table 3.** Estimates from logistic regression prediction model to classify pulmonary tuberculosis using variables from initial evaluation. Parameter estimate with 95% confidence intervals, odds ratio estimate with 95% confidence interval, and p-values for each parameter included in the logistic regression prediction model. The estimate provided for each predictor is computed against a reference that reflects the absence of that feature. The model parameter estimates account for potential clustering at the study-level as well as uncertainty introduced by missing data. *P-value calculated using Rubin’s rules for multiple imputed data. OR – odds ratio, CI – confidence interval, CXR – chest x-ray.

| Predictors | Coefficient | Coefficient 95% CI | OR | OR 95% CI | P-value* |
| --- | --- | --- | --- | --- | --- |
| Intercept | -1.92 | -2.58, -1.25 | -- | -- | -- |
| Cough duration ≥2 weeks<br>(Absence is no cough or cough <2 weeks) | 0.17 | -0.09, 0.43 | 1.19 | 0.91, 1.54 | 0.86 |
| Fever duration ≥2 weeks<br>(Absence is no fever or fever <2 weeks) | 0.45 | 0.16, 0.74 | 1.57 | 1.18, 2.09 | 0.25 |
| Lethargy | 0.25 | 0.02, 0.48 | 1.28 | 1.02, 1.62 | 0.66 |
| Weight loss | 0.22 | -0.03, 0.48 | 1.25 | 0.97, 1.62 | 0.75 |
| History of documented tuberculosis exposure | 1.43 | 0.87, 2.00 | 4.20 | 2.39, 7.38 | <0.001 |
| Haemoptysis | 0.34 | -0.37, 1.05 | 1.40 | 0.69, 2.86 | 0.78 |
| Night sweats | 0.2 | 0.02, 0.38 | 1.22 | 1.02, 1.47 | 0.71 |
| Peripheral lymphadenopathy | 0.35 | 0.13, 0.57 | 1.42 | 1.14, 1.77 | 0.35 |
| Temperature ≥38 Celsius | 0.00 | -0.25, 0.26 | 1.00 | 0.78, 1.30 | >0.999 |
| Tachycardia | 0.15 | -0.13, 0.42 | 1.16 | 0.88, 1.53 | 0.90 |
| Tachypnoea | -0.05 | -0.27, 0.16 | 0.95 | 0.77, 1.18 | 0.98 |
| Cavities on CXR | 0.47 | -0.11, 1.05 | 1.60 | 0.90, 2.85 | 0.53 |
| Intrathoracic lymphadenopathy on CXR | 1.46 | 1.00, 1.92 | 4.32 | 2.73, 6.85 | <0.001 |
| Opacities on CXR | 0.43 | 0.02, 0.84 | 1.54 | 1.02, 2.32 | 0.45 |
| Miliary infiltrate on CXR | 1.27 | 0.57, 1.97 | 3.56 | 1.76, 7.19 | <0.001 |
| Pleural effusion on CXR | 0.64 | 0.2, 1.09 | 1.90 | 1.22, 2.96 | 0.13 |

### Algorithm development

The scores derived from the model prediction coefficients that correspond to classification of all tuberculosis with respective sensitivities of 90%, 85%, 80%, 75%, and 70% can be found in <u>Supplementary Appendix R</u>. The study-level and summary performance of these scores in classifying tuberculosis can be found in <u>Supplementary Appendix S</u>.

To balance the consequences of untreated tuberculosis versus the consequences of overtreatment, the expert group recommended a sensitivity threshold of 85% in classifying tuberculosis using the composite reference standard, resulting in the development of a score with a sensitivity of 0·86 (95% CI: 0·68-0·94) and a specificity of 0·37 (95% CI: 0·15-0·66) (**Figure 2**). An analysis of the performance in classifying confirmed tuberculosis vs. unlikely tuberculosis demonstrated a sensitivity of 0·88 (95% CI: 0·71-0·95) and specificity of 0·37 (95% CI: 0·15-0·67) (<u>Supplementary Appendix T</u>). Under a sensitivity threshold of 85%, the model that included only features from the baseline clinical evaluation (without chest x-ray findings) had a sensitivity of 0·84 (95% CI: 0·66-0·93) and specificity of 0·30 (95% CI: 0·13-0·56) in classifying tuberculosis (<u>Supplementary Appendix U</u>).

**Figure 2.**
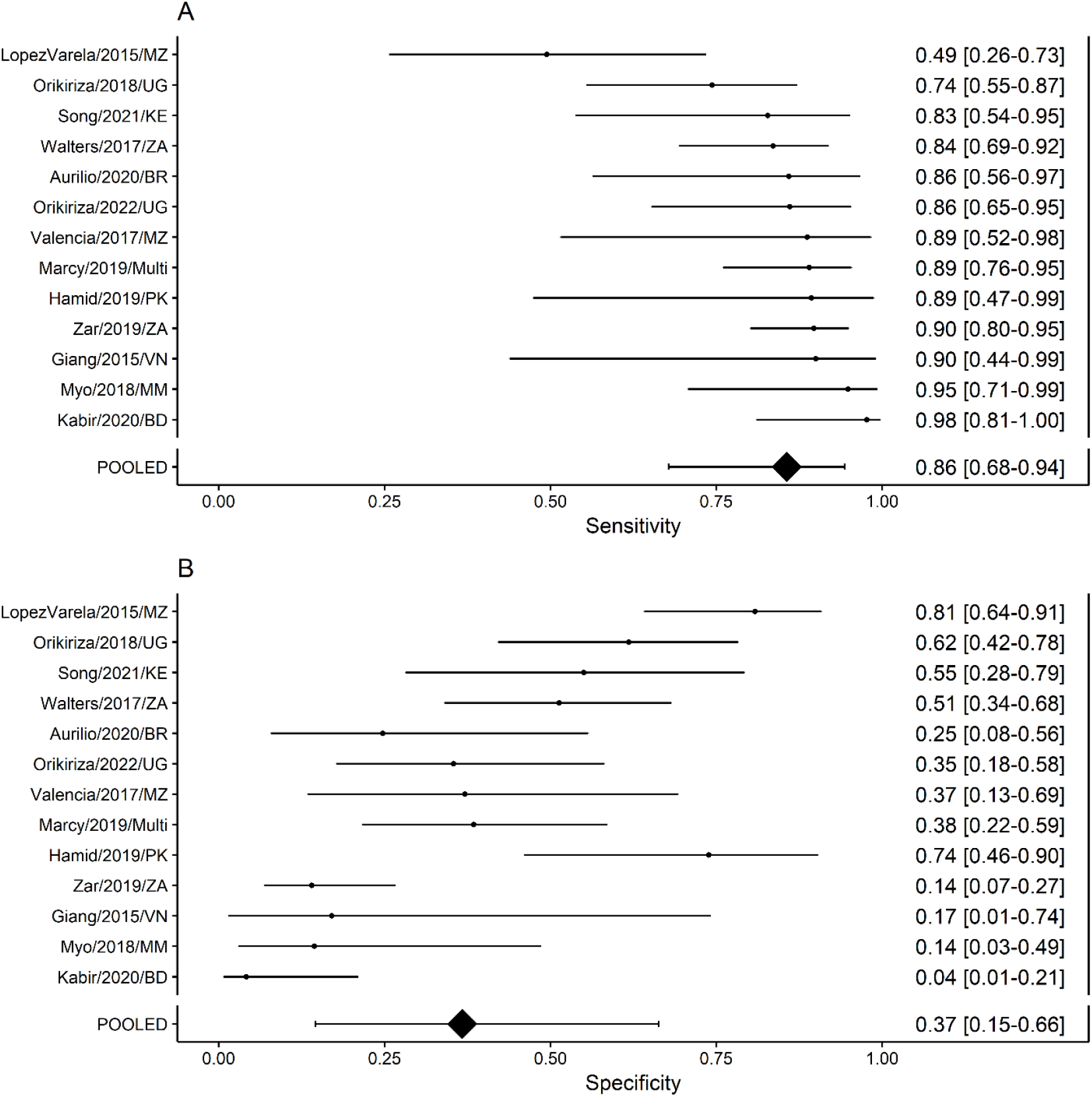
Forest plot depicting performance of scaled scores from prediction model to classify tuberculosis with 85% sensitivity. Study-level and pooled estimates of the **(A)** sensitivity and **(B)** specificity of classifying tuberculosis (composite reference standard: bacteriologically-confirmed pulmonary tuberculosis and unconfirmed pulmonary tuberculosis) of the scores derived from the prediction model developed from the IPD to classify TB with 85% sensitivity. IPD – individual participant data, BD – Bangladesh, BR – Brazil, KE – Kenya, MM – Myanmar, Multi – Multi-country study (includes Burkina Faso, Cameroon, Vietnam, and Cambodia), MZ – Mozambique, PK – Pakistan, UG – Uganda, VN – Vietnam, ZA – South Africa.

To adapt the scores into treatment-decision algorithms to be used at primary healthcare centres, the expert group recommended the following triage steps prior to classification using the score: 1) identifying children with clinical symptoms and signs requiring urgent referral to higher levels of healthcare, and 2) stratifying children by risk of mortality and progression of tuberculosis. Higher-risk children were defined by the expert group as those <2 years, severely malnourished, and/or living with HIV. These children would be evaluated using the score at the time of the initial evaluation. Children not meeting this definition would be treated for the most likely non-tuberculosis condition and complete re-evaluation in 1-2 weeks: those with persistent/worsening symptoms at follow-up would be evaluated using the score. The expert group additionally recommended to pursue, wherever available, bacteriological testing on respiratory and/or stool specimens with rapid molecular diagnostics for all children and urine lateral flow assays for HIV-positive children to align with existing WHO recommendations^23^.

The expert group recommendations resulted in the development of a treatment-decision algorithm (**Figure 3**), in which children <10 years with presumptive pulmonary tuberculosis are triaged by risk of tuberculosis-related morbidity and mortality prior to being evaluated for the presence of clinical and chest x-ray features to assign a score corresponding to tuberculosis risk. A total score of >10 results in classification of tuberculosis with a sensitivity of 85%. Known exposure to tuberculosis alone has a score >10, so this was placed above the other elements. The same parameters were used to construct the treatment-decision algorithm from the model without chest x-ray features (<u>Supplementary Appendix V</u>).

**Figure 3.**
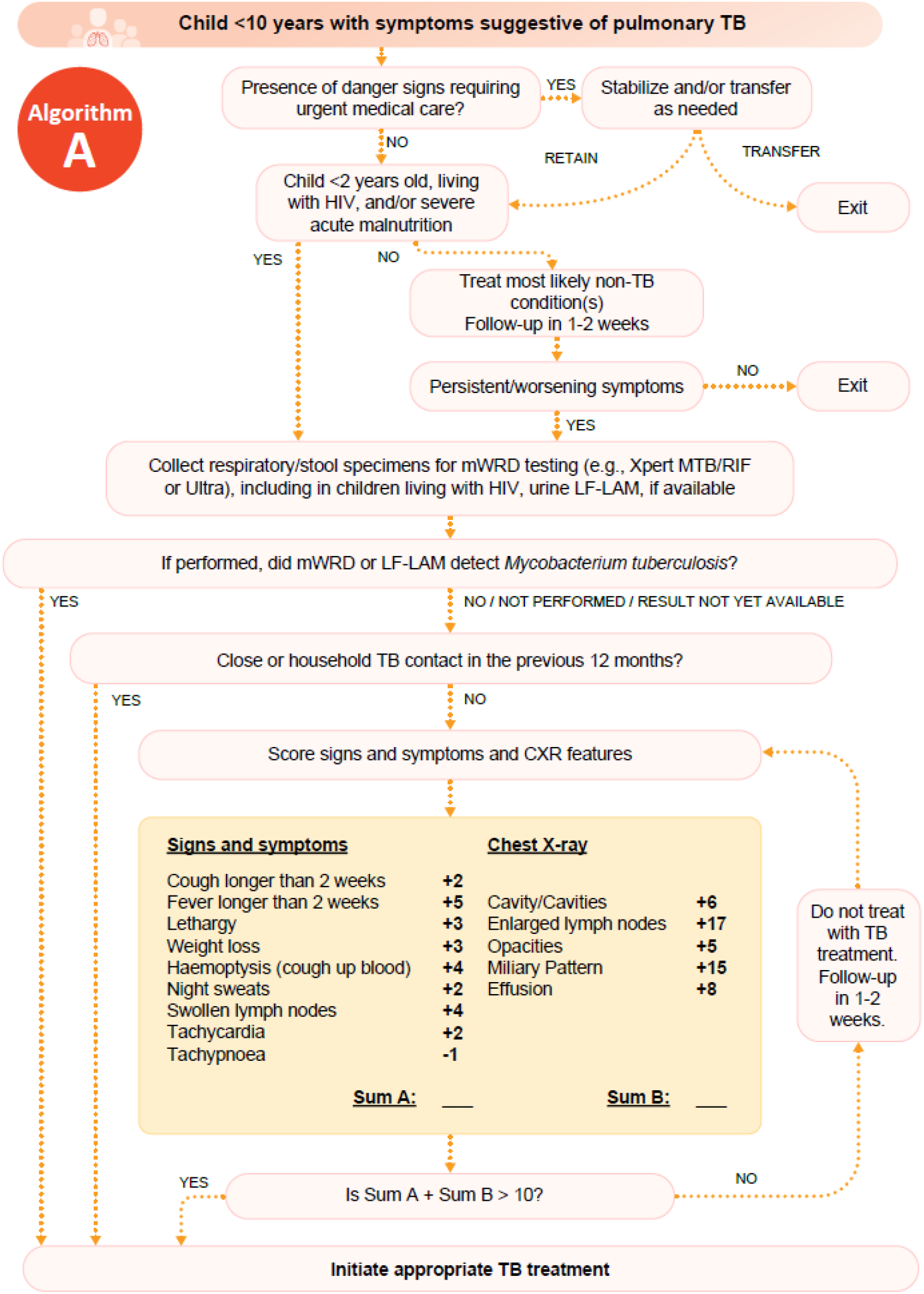
Treatment-decision algorithm including chest x-ray features derived from the prediction model. Tuberculosis treatment-decision algorithm for use among children <10 years with symptoms suggestive of pulmonary tuberculosis, reproduced from the operational handbook accompanying the WHO consolidated guidelines on the management of tuberculosis in children and adolescents.^12,13^ Selection steps prior to entering scoring system reflect recommendations from the WHO expert panel to enrich the probability of tuberculosis among the population of children proceeding through the algorithm to the model such that the probability would more closely reflect the preselected population producing the data from which the prediction model was built while balancing the consequences of untreated tuberculosis among high-risk children. Scores associated with features from clinical history and physical exam and chest X-ray translate to risk of tuberculosis and are scaled from the prediction model developed from the IPD. Guidance on the practical use of this algorithm is outlined in the WHO operational handbook. WHO – World Health Organization, TB – tuberculosis, IPD – individual participant data, HIV – human immunodeficiency virus, mWRD – molecular WHO-recommended rapid diagnostic test, CLHIV – children living with HIV, LF-LAM – lateral flow urine lipoarabinomannan assay, CXR – chest X-ray.

## DISCUSSION

We assembled a large IPD dataset from nearly five thousand children from geographically diverse, high-tuberculosis incidence settings to evaluate existing treatment-decision algorithms and develop new evidence-based treatment-decision algorithms to guide evaluation of children with presumptive pulmonary tuberculosis. This algorithm-building approach uses the best currently available data to provide practical guidance to healthcare workers in primary healthcare settings to identify which clinical features, with or without chest x-ray assessment, indicate whether initiation of tuberculosis treatment is warranted. The newly developed algorithms were incorporated into the WHO operational handbook to support implementation of the new consolidated guidelines.^12,13^

Modelling diagnostic evaluations IPD provides a quantitative description of the strength of evidence for childhood pulmonary tuberculosis treatment-initiation decisions. Of the clinical features, only reported exposure to tuberculosis was independently sufficient to meet the threshold for treatment initiation. This was true even in the model-based score without chest x-ray features, suggesting that none of the common clinical features could independently inform highly sensitive/specific treatment decisions. While results from tuberculin skin testing were used by studies to inform classification of tuberculosis and to improve imputation of missing data, we did not include these in the algorithms given operational limitations in using tuberculin skin testing at scale in high-burden settings. Of the chest X-ray features included in the algorithm, the presence of intrathoracic lymphadenopathy and a miliary pattern, respectively, were independently sufficient to start treatment. It is worth noting that inclusion of chest x-ray features only increased the specificity of the score slightly as compared to the score developed from the model with clinical features only. Chest x-ray has additional utility in guiding childhood pulmonary tuberculosis treatment duration for severe vs. non-severe disease,^24^ in monitoring TB treatment response (including associated complications and sequelae) and in the diagnosis of other non-tuberculosis intrathoracic pathology.

The decision to prioritise sensitivity in our algorithm development is critical to initiate more children with tuberculosis on appropriate therapy; however, many children may be falsely treated for tuberculosis given the resulting specificity. No test or algorithm meets the WHO-target sensitivity and specificity for a confirmatory diagnostic test for childhood pulmonary tuberculosis.^25^ Thus, the expert group advised to develop an algorithm with a minimum sensitivity target of 85% as an acceptable balance between sensitivity and the resulting specificity. Tuberculosis treatment for drug-susceptible disease is relatively safe in children;^26^ especially considering the potentially severe consequences of a missed tuberculosis diagnosis. However, falsely treating for tuberculosis carries risk of delayed diagnosis of other disease, drug adverse events, and unnecessary burden on families and healthcare services. It should be noted that while these performance estimates relate to the score component of the algorithm, the overall sensitivity and specificity of the whole algorithm, including the triage steps, remains unknown and should be evaluated prospectively. As low-risk children are made to wait prior to being evaluated with the scored part of the algorithm, symptoms in some with diagnoses other than tuberculosis will resolve, likely improving specificity.

We note that the model-based scoring component of the algorithm demonstrates considerable study-level heterogeneity in sensitivity and specificity. Although this IPD is the largest of its size compiled to date, there were not enough studies to quantitatively describe the features that drive the observed heterogeneity. Given that we used data made available to WHO following a public call rather than conducting a systematic review, it is possible that some diagnostic studies may have been excluded. The inclusion of more data from existing, ongoing, and future studies, may allow meta-regression to describe study-level sources of heterogeneity. Heterogeneity may have been driven in part by varied tuberculosis prevalence in the cohorts included as well as heterogeneities in disease presentation. Given that the pre-existing treatment-decision algorithms demonstrated similar heterogeneities in performance compared to the evidence-based algorithm developed, we suggest that our approach is robust as it offers the flexibility to further interrogate the sources of heterogeneity as additional data are available. A distinct advantage of the modelling approach we used for algorithm development is the ability to revise and calibrate the model to specific settings as additional data become available.

We considered it important to evaluate existing treatment-decision algorithms and develop new algorithms using a composite standard rather than solely a microbiological standard, given the high percentage of children treated for tuberculosis without bacteriological confirmation, even in the best resourced settings, which reflects the paucibacillary nature of disease in most young children. However, this reference standard remains imperfect, and misclassification may occur.^27^ The underlying composition of the unconfirmed tuberculosis group may represent a heterogeneous group in which some children have tuberculosis, and some have other causes for their observed symptoms and signs. Additionally, it is possible that inclusion of unconfirmed pulmonary tuberculosis biased the estimation of the prediction model parameters, especially those used to classify the unconfirmed group. Although this is a limitation of our study, the similar performance estimates of the score developed in the primary analysis using both the composite and confirmed tuberculosis reference standards suggest that this may not be a major issue.

Given that our algorithms are intended to guide decisions to treat children in primary healthcare centres, it is a limitation that IPD was derived from primarily tertiary and referral health centres. We are not aware of studies that provide this quality of diagnostic evaluations data from presumptive childhood tuberculosis in primary healthcare centres. However, in several studies, children presenting at primary healthcare settings were directly referred for study evaluation, providing some degree of reassurance as to the generalisability of results. The pre-test probability of tuberculosis (i.e., the prevalence) is likely substantially lower among children attending primary healthcare centres and the clinical presentation may be different as compared to tertiary and referral centres from which the data were obtained. These are important given that many children with tuberculosis first present to primary healthcare centres.^28^ We believe that the risk-stratification and delayed entry of lower risk children with presumptive tuberculosis (who should tolerate the delay) is a practical attempt to safely raise the pre-test probability when implementing the algorithm in primary health centres. Prospective external validation of the entire algorithms will be critical to determining their accuracy, acceptability and feasibility of use at different levels of the healthcare system.

There are inherent limitations to developing a prediction model developed on data from multiple cohorts for a disease with an imperfect diagnostic gold standard. Study inclusion criteria varied, which impacts the baseline tuberculosis prevalence and applicability of the score prediction estimates. Additionally, prediction variable definitions varied among the included studies—for example, history of weight loss was variably defined as caregiver reported history of weight loss or objective weight loss and/or deviation from previous growth trajectory. This heterogeneity is also true for the study-level reference classifications, especially for unconfirmed tuberculosis. Some studies used a previous version of the NIH reference classification, which included probable and possible tuberculosis categories that we reclassified as unconfirmed tuberculosis, despite limitations using this approach.^29^ These may contribute to heterogeneities in estimating the association between the predictors and the outcome of tuberculosis. Finally, we note that using a prespecified prediction model, as we did, may lead to overfitting.^30^ Despite a reasonable summary O:E ratio for our model, the heterogeneity in study-level O:E ratio demonstrated in our internal-external cross-validation suggests that overfitting may be an issue. As more data become available, future investigation into the causes driving heterogeneity may inform more nuanced use of this algorithm within specific contexts and populations.

Pragmatic treatment-decision algorithms can lead to better detection of tuberculosis in children, with improved access to early treatment and reduced tuberculosis morbidity and mortality. Although we developed these algorithms using a thorough modelling analysis of a large high-quality IPD dataset, the disappointing specificity of the scoring component suggests that improved diagnostic tools will be necessary to meet sensitivity and specificity targets. As these diagnostic tools are identified, their data may be incorporated into treatment-decision algorithms to improve the specificity of the algorithms while maintaining high sensitivity. Additionally, decision-analytic modelling of the relative “cost” of false positive versus false negative classification of tuberculosis and prevalence of tuberculosis may provide insight to select an appropriate sensitivity threshold in future algorithm development.

Treatment-decision algorithms are now conditionally recommended by the WHO in the evaluation of children with presumptive tuberculosis, which should lead to improved diagnostic capacity at and treatment initiation at primary healthcare centres where paediatric tuberculosis expertise may be lacking. This work represents a paradigm shift in pragmatic and evidence-based approaches using advanced analytic methods to develop algorithms that draw on the best globally available data. This approach can be further improved and interrogated as additional data and diagnostic tools become available.

## Data Availability

Data is available upon written request to the authors.

## AUTHOR CONTRIBUTIONS

Conceptualization: KSG, OM, JAS

Data curation: KSG, OM, EL-V, MFF, MB, SA, FA, AA, OA, RBA, SB, IB, KPC, LC, MC, ESC, ALG-B, ACH, JH, SK, LL, AAM, KM, DN, MPN, PO, MP, CCS, SAS, JPS, RS, NTTT, VU, MMvdZ, EGW, HJZ

Formal analysis: KSG, OM, JM, JLW, TPAD, TC, JAS

Funding acquisition: KSG, AB, SV, KV, TC, JAS

Investigation: KSG, OM, JM, EL-V, MPS, MFF, MB, SA, FA, AA, OA, RBA, SB, IB, AB, KPC, LC, MC, ESC, ALG-B, ACH, JH, SK, AM, FM, AAM, KM, DN, MPN, PO, MP, CCS, SAS, JPS, RS, NTTT, VU, MMvdZ, SV, KV, EGW, JLW, HJZ, BJM, SMG, TPAD, TC, JAS

Methodology: KSG, OM, JM, AB, SV, KV, TPAD, TC, JAS

Software: JM, TPAD

Validation: KSG, JAS

Visualization: KSG

Writing - original draft: KSG, TC, JAS

Writing - review & editing: all authors

## ROLE OF THE FUNDING SOURCE

This work was supported by the WHO Global Tuberculosis Programme (GTB) as well as the Eunice Kennedy Shriver National Institute of Child Health & Human Development of the United States National Institutes of Health (NIH). It was used to inform WHO consolidated guidelines on tuberculosis management of TB in children and adolescents and the accompanying operational handbook, both published by WHO in March 2022. This research was also supported in part by the President’s Emergency Plan for AIDS Relief (PEPFAR) through the Centers for Disease Control and Prevention (CDC).

### Disclaimer

Staff from the WHO GTB as well as staff from the CDC are co-authors of this paper, offered feedback on the methodology and investigation, reviewed the manuscript, and made the decision to submit. The authors alone are responsible for the views expressed in this article and they do not necessarily represent the views, decisions or policies of the NIH, the WHO, the CDC, and PEPFAR.

## ACKNOWLEDGEMENTS

KSG was supported by the NIH the Eunice Kennedy Shriver National Institute of Child Health and Human Development [F30HD105440] as well as the Yale Medical Scientist Training Program [T32GM007205]. SA, SB and SK received funding support from USAID through Research for Decision Makers’ Activity, and the paper was prepared through the Structured Operational Research and Training Initiative (SORT IT), a global partnership led by the Special Program for Research and Training in Tropical Diseases at the World Health Organization. LMC was supported by the NIH/NIAID (K23 AI143479). RS was supported by the NIH through the Eunice Kennedy Shriver National Institute of Child Health and Human Development (K23HD072802). MMVDZ is supported by a career development grant from the EDCTP2 program supported by the European Union (TMA2019SFP-2836 TB lung-FACT2), the Fogarty International Center of the National Institutes of Health (Award Number K43TW011028) and by funding from the South African Medical Research Council. EW was supported by a scholarship for doctoral studies from the Medical Research Council of South Africa under MRC Clinician Researcher Programme, the Faculty of Medicine and Health Sciences at Stellenbosch University (Early Career Grant and Temporary Research Assistantship grant), the Harry Crossley Foundation and the South African National Research Foundation (Thuthuka programme funding for doctoral students). HJZ and MPN received funding from the NIH (R01HD058971) and the Regional Prospective Observational Research in Tuberculosis Consortium, co-funded by MRC of South Africa and NIH. TC and JLW were supported by the NIH through the Institute of Allergy and Infectious Disease [R01AI147854 and R01AI137093, respectively]. JAS was supported by a Clinician Scientist Fellowship jointly funded by the UK Medical Research Council (MRC) and the UK Department for International Development (DFID) under the MRC/DFID Concordat agreement (MR/R007942/1).

We acknowledge the patients and their caregivers for their willingness to participate in each individual study. We hope that their contribution will benefit other families affected by tuberculosis. We additionally would like to thank the following individuals who contributed to execution of this work: Albert Okumu, Andrea T. Cruz, Carlos M. Perez-Velez, Chad Heilig, Colleen Wright, Elisha Okeyo, Ha Thi Minh Ðang, Hendrik Simon Schaaf, James Orwa, Lazarus Odeny, Lesley Workman, Maria R. Jaswal, Mariaem Andres, Mark Fajans, Moe Zaw, Parisa Hariri, Patrice Ahenda, Prisca Rabuogi, Rose Abwunza, Rumana Nasrin, Sabrina Choudhury, Scott Lee, Shaikh Shumail, Shoaib Ahmed, Susan Musau, Syed Mohammad Mazidur Rahman, Walter Mchembere, Yi Kyaw, Leonardo Martinez, Vivian Cox, Bryan Vonasek, and Alexander Kay. Finally, we would like to acknowledge the members of the WHO Guideline Development Group who considered this evidence to inform the 2022 WHO consolidated guidelines on the management of tuberculosis in children and adolescents.

## DATA AVAILABILITY

Data is available upon written request to the authors.

