## Supplement for "Development and validation of treatment-decision algorithms for children evaluated for pulmonary tuberculosis: an individual participant data meta-analysis"

### Abbreviations and shorthand

AUC – area under the receiver-operator curve

BCG - Bacille Calmette-Guérin vaccine

BD – Bangladesh

BMI – body mass index

BR – Brazil

CI – confidence interval

c-Statistic – concordance statistic

CXR – chest X-ray

EPTB – extrapulmonary tuberculosis

ES – expectorated sputum

GA – gastric aspirate

ART –Antiretroviral therapy

HIV – human immunodeficiency virus

IPD – individual participant data

IS – induced sputum

KE – Kenya

LF-LAM – lateral flow urine lipoarabinomannan assay

MICE – multiple imputation by chained equations

MM – Myanmar

MoH – Ministry of Health

*Mtb –* Mycobacterium tuberculosis

Multi – (PAANTHER) Multi-country study (includes Burkina Faso, Cameroon, Vietnam, and Cambodia)

mWRD – molecular WHO-recommended rapid diagnostic test

MZ – Mozambique

NTLP – National TB and Leprosy Program

O:E – observed: expected ratio

OR – odds ratio

PAANTHER - Pediatric Asian African Network for Tuberculosis and HIV Research

PK – Pakistan

PPD – purified protein derivative

PTB – pulmonary tuberculosis

SAM – severely acutely malnourished

TB – tuberculosis

TRIPOD - transparent reporting of a multivariable prediction model for individual prognosis or diagnosis

TST – tuberculin skin test

UG – Uganda

VN – Vietnam

WFAZ – weight-for-age Z-score

WFL – weight-for-length

WHO – World Health Organization

Xpert – Xpert MTB/RIF

Xpert Ultra – Xpert MTB/RIF Ultra

ZA – South Africa

### Appendix A: Data requested

**Table S1. Data requested from studies and suggested format**

| **FIELD** | **VARIABLE** | **DESCRIPTION** | **FORMAT** | **CODE** | **LABEL** |
| --- | --- | --- | --- | --- | --- |
| studyID | Study ID | Data source cohort | int | 1 | Aurilio/2020/BR |
|  |  |  |  | 2 | Song/2021/KE |
|  |  |  |  | 3 | LopezVarela/2015/MZ |
|  |  |  |  | 4 | Valencia/2017/MZ |
|  |  |  |  | 5 | Myo/2018/MM |
|  |  |  |  | 6 | Marcy/2016/Multi |
|  |  |  |  | 7 | Hamid/2019/PK |
|  |  |  |  | 8 | Walters/2017/ZA |
|  |  |  |  | 9 | Orikiriza/2018/UG |
|  |  |  |  | 10 | Orikiriza/2021/UG |
|  |  |  |  | 11 | Giang/2015/VN |
|  |  |  |  | 12 | Kabir/2020/BD |
|  |  |  |  | 13 | Zar/2019/ZA |
| age | Age (months) | Age (months) at enrolment | num | ### | NA = unknown |
| sex | Sex | Participant sex | int | 0 | Female |
|  |  |  |  | 1 | Male |
|  |  |  |  | NA | Unknown |
| weight | Weight (kg) | Weight (kg) at initial evaluation | num | ### | NA = unknown |
| height | Height (cm) | Height/length (cm) at initial evaluation | num | ### | NA = unknown |
| bcg_evidence | BCG vaccination | Evidence of BGC vaccination (BCG scar or BCG recorded in immunization record) at initial evaluation | int | 0 | No evidence of BCG vaccination |
|  |  |  |  | 1 | Evidence of BCG vaccination |
|  |  |  |  | NA | Unknown |
| HIV_status | HIV status | Participant HIV status | int | 0 | HIV-negative |
|  |  |  |  | 1 | HIV-positive |
|  |  |  |  | NA | Unknown |
| cough_less2wk_gr2wk_gr3wk_gr4wk | Cough duration | Duration of cough at initial evaluation | int | 0 | No cough |
|  |  |  |  | 1 | Cough 0-13 days |
|  |  |  |  | 2 | Cough 14-20 days |
|  |  |  |  | 3 | Cough 21-27 days |
|  |  |  |  | 4 | Cough ≥28 days |
|  |  |  |  | NA | Unknown |
| cough_greater_2wk | Cough duration | Presence of cough ≥2 weeks at initial evaluation | int | 0 | Cough ≥2 weeks not present |
|  |  |  |  | 1 | Cough ≥2 weeks present |
|  |  |  |  | NA | Unknown |
| fever_less2wk_gr2wk_gr3wk_gr4wk | Fever duration | Duration of fever at initial evaluation | int | 0 | No fever |
|  |  |  |  | 1 | Fever 0-13 days |
|  |  |  |  | 2 | Fever 14-20 days |
|  |  |  |  | 3 | Fever 21-27 days |
|  |  |  |  | 4 | Fever ≥28 days |
|  |  |  |  | NA | Unknown |
| fever_greater_1wk | Fever duration | Presence of fever ≥1 week at initial evaluation | int | 0 | Fever ≥1 week not present |
|  |  |  |  | 1 | Fever ≥1 week present |
|  |  |  |  | NA | Unknown |
| lethargy | Lethargy | Presenting history of unusual lethargy or lack of playfulness at initial evaluation | int | 0 | No lethargy |
|  |  |  |  | 1 | Lethargy |
|  |  |  |  | NA | Unknown |
| weight_loss | Weight loss | Presenting history of poor growth over the preceding 3 months AND not responding to nutritional rehabilitation (or antiretroviral therapy if HIV infected) | int | 0 | No weight loss |
|  |  |  |  | 1 | Weight loss |
|  |  |  |  | NA | Unknown |
| significant_tbc | Documented TB exposure | Documented exposure to MTB at initial evaluation in previous 12 months | int | 0 | No documented TB exposure in previous 12 months |
|  |  |  |  | 1 | Documented TB exposure in previous 12 months |
|  |  |  |  | NA | Unknown |
| night_sweats | Night sweats | Presenting history of night sweats at initial evaluation | int | 0 | No night sweats |
|  |  |  |  | 1 | Night sweats |
|  |  |  |  | NA | Unknown |
| hemoptysis | Haemoptysis | Presenting history of haemoptysis at initial evaluation | int | 0 | No haemoptysis |
|  |  |  |  | 1 | Haemoptysis |
|  |  |  |  | NA | Unknown |
| temp | Temperature (C) | Recorded temperature at initial evaluation | num | ### | NA=unknown |
| heart_rate | Heart rate (per min) | Heart rate (per minute) at initial evaluation | num | ### | NA=unknown |
| respiratory_rate | Respiratory rate (per min) | Respiratory rate (per minute) at initial evaluation | num | ### | NA=unknown |
| peripheral_lad | Peripheral lymphadenopathy | Peripheral lymphadenopathy (at cervical, submandibular, and/or axillary nodes) at initial evaluation | int | 0 | No peripheral lymphadenopathy |
|  |  |  |  | 1 | Peripheral lymphadenopathy |
|  |  |  |  | NA | Unknown |
| first_xpert_result | First Xpert MTB/RIF | Result from first Xpert MTB/RIF (not Ultra) performed on ES/IS (or GA for young children) collected at initial evaluation | int | 0 | Xpert negative for Mtb |
|  |  |  |  | 1 | Xpert positive for Mtb |
|  |  |  |  | NA | Unknown/not performed |
| CXRfinal_result | CXR consistent with TB | Result of CXR performed at initial evaluation as assessed by reader performing clinical evaluation/making TB-treatment decision or by reader to inform research classification of TB if former not available | int | 0 | CXR not consistent with TB |
|  |  |  |  | 1 | CXR consistent with TB |
|  |  |  |  | NA | Unknown/not assessed |
| CXRopacity_result | Opacities on CXR | Opacities (e.g., alveolar consolidation and/or bronchopneumonia) on CXR performed at initial evaluation as assessed by reader performing clinical evaluation/making TB-treatment decision or by reader to inform research classification of TB if former not available | int | 0 | Opacities not present on CXR |
|  |  |  |  | 1 | Opacities present on CXR |
|  |  |  |  | NA | Unknown/not assessed |
| CXRcavity_result | Cavities on CXR | Cavities on CXR performed at initial evaluation as assessed by reader performing clinical evaluation/making TB-treatment decision or by reader to inform research classification of TB if former not available | int | 0 | Cavities not present on CXR |
|  |  |  |  | 1 | Cavities present on CXR |
|  |  |  |  | NA | Unknown/not assessed |
| CXRmili_result | Miliary infiltrate on CXR | Miliary infiltrate on CXR performed at initial evaluation as assessed by reader performing clinical evaluation/making TB-treatment decision or by reader to inform research classification of TB if former not available | int | 0 | Miliary infiltrate not present on CXR |
|  |  |  |  | 1 | Miliary infiltrate present on CXR |
|  |  |  |  | NA | Unknown/not assessed |
| CXRnodes_result | Intrathoracic lymphadenopathy on CXR | Intrathoracic lymphadenopathy (e.g., perihilar nodes, paratracheal nodes, mediastinal nodes) on CXR performed at initial evaluation as assessed by reader performing clinical evaluation/making TB-treatment decision or by reader to inform research classification of TB if former not available | int | 0 | Intrathoracic lymphadenopathy not present on CXR |
|  |  |  |  | 1 | Intrathoracic lymphadenopathy present on CXR |
|  |  |  |  | NA | Unknown/not assessed |
| CXReffusion_result | Pleural effusion on CXR | Pleural effusion on CXR performed at initial evaluation as assessed by reader performing clinical evaluation/making TB-treatment decision or by reader to inform research classification of TB if former not available | int | 0 | Pleural effusion not present on CXR |
|  |  |  |  | 1 | Pleural effusion present on CXR |
|  |  |  |  | NA | Unknown/not assessed |
| TST_result | Tuberculin skin test | Tuberculin skin test positive at initial evaluation | num | 0 | TST negative |
|  |  |  |  | 1 | TST positive |
|  |  |  |  | NA | Unknown/not performed |
| TB_classification | TB classification | Final classification of TB | int | 0 | Unlikely TB |
|  |  |  |  | 1 | Bacteriologically-confirmed TB |
|  |  |  |  | 2 | Unconfirmed TB |
|  |  |  |  | NA | Unknown |

### Appendix B: Information about multiple imputation by chained equations

Imputation of missing data was carried out using the multiple imputation by chained equations (MICE) methods implemented in the R package, *mice*.^1^ MICE is a fully conditionally specified modeling approach that first imputes the mean for missing data in each variable, then uses regression modeling to re-impute missing data in each variable by conditioning on the remaining variables, and finally iteratively updates imputations using the newly imputed data.

All IPD specified in Table S1 were included in the imputation models. We included a cluster-specific random effects term in the imputation model for each variable whenever possible to allow for study-level heterogeneities in the baseline distribution for each imputed variable. For continuous variables and categorical variables, we used a two-level predictive mean matching model implemented using the “2l.pmm” method in the R package, *miceadds*.^2^ For binary variables, we used a two-level logistic model implemented using the “2l.bin” method. Few binary variables gave a singular fit warnings using two-level methods; for these variables, we reflexed to using a one-level logistic model implemented using the “logreg” method. MICE was run using the “mice” function with 20 iterations to generate 100 imputed datasets. The methods used to impute each variable are specified as follows:

*2l.pmm*: cough_less2wk_gr2wk_gr3wk_gr4wk, fever_less2wk_gr2wk_gr3wk_gr4wk, weight, temp, heart_rate, respiratory_rate, height, TB_classification

*2l.bin*: cough_greater_2wk, fever_greater_1wk, lethargy, weight_loss, night_sweats, peripheral_lad, significant_tbc, bcg_evidence, HIV_status, CXRfinal_result, CXRnodes_result, CXRopacity_result, CXReffusion_result, TST_result, CXRcavity_result, first_xpert_result

*logreg*: sex, haemoptysis, CXRmili_result

Additional imputation specifications are as follows:

*Cluster variable*: studyID

*No imputation method specified (fully complete data)*: studyID, age

Convergence of imputation was visually assessed by trace plots of the mean and standard deviation of each variable over the imputation iterations.

### Appendix C: Estimation of algorithm performance accounting for multiply imputed data

For each algorithm, sensitivity and specificity estimates were computed at the study-level and pooled using a bivariate model as implemented in the “reitsma” function in the *mada* package in R.^3^ To account for the uncertainty associated with missing data, we imputed 100 datasets as specified in Appendix B. Study-level and pooled estimates of sensitivity and specificity were computed for each of the 100 datasets. We pooled the sensitivity and specificity estimates computed at the level of each imputed dataset using Rubin’s rules to yield a point estimate and 95% confidence intervals for the study-level and for the overall pooled estimates computed by the “reitsma” function.^4^

### Appendix D: Develop a score from models produced on multiply imputed data

A general form of a multivariate logistic regression equation is given as follows:

$$logit\left( p \right)=\beta_{0}+\beta_{1}*x_{1}+ \beta_{2}*x_{2}+\ldots+ \beta_{n}*x_{n}$$

Where p is the probability of tuberculosis, $x_{1\ldots n}$ refers to the predictors and $\beta_{1\ldots n}$ refers to the coefficients describing the relationship between the predictor and the logit-transformed probability. We fit the prediction model to the data, and we identified the probability corresponding to classification of tuberculosis with a given sensitivity compared to the reference standard. For example, let us specify an interest in classifying tuberculosis with a sensitivity of at least 85%. We obtained a threshold probability by subtracting the intercept from the logit-transformed probability corresponding to diagnosis with at least an 85% sensitivity. We scaled the threshold probability to 10 by multiplying by a scaling factor, and we multiplied the coefficients for each predictor by that scaling factor to obtain the score for that predictor. Thus, the score for each individual meeting entry criteria was obtained by summing the scaled coefficients for each factor present in the patient, and a total score of >10 constituted a diagnosis of tuberculosis with a sensitivity of 85% using this treatment-decision algorithm.

Given that we generated multiple imputed datasets, we had to take additional steps to determine the probability threshold and pooled coefficient estimates. We used the “metapred” function in package *metamisc* to fit a logistic regression model on each imputed dataset, resulting in 100 logistic regression models.^5^ Note that the model generated by “metapred” is a pooled model of models with the same specifications fit at the study-level. A pooled estimate of each parameter coefficient was obtained by taking the mean of each of the 100 coefficient estimates for each parameter. A pooled probability threshold was determined by taking the mean of the probability threshold corresponding to classification of tuberculosis with a given sensitivity compared to the reference standard of each model. The pooled probability threshold and pooled coefficient estimates were used to produce the scores as described above.

### Appendix E: Composition of group of experts advising algorithm development

- *Farhana Amanullah*: Indus Hospital & Health Network, Karachi, Pakistan
- *Annemieke Brands*: Global Tuberculosis Programme, World Health Organization, Geneva, Switzerland
- *Stephen Graham*: Centre for International Child Health, University of Melbourne Department of Paediatrics and Murdoch Children’s Research Institute, Royal Children’s Hospital, Melbourne, Australia and International Union Against Tuberculosis and Lung Disease (The Union), Paris, France
- *Kenneth Gunasekera*: Department of Epidemiology of Microbial Diseases, Yale School of Public Health, New Haven, United States of America
- *Alexander Kay*: Global TB Program, Baylor College of Medicine and Texas Children's Hospital, Houston, United States of America
- *Anna Mandalakas*: Global TB Program, Baylor College of Medicine and Texas Children's Hospital, Houston, United States of America
- *Ben Marais*: The Children’s Hospital at Westmead Clinical School, Faculty of Medicine and Health, University of Sydney, Sydney Australia
- *Olivier Marcy*: University of Bordeaux, Inserm, Institut de Recherche pour le Développement, Bordeaux, France
- *James Seddon*: Desmond Tutu Tuberculosis Centre, Department of Paediatrics and Child Health, Faculty of Medicine and Health Sciences, Stellenbosch University, Tygerberg, South Africa and Department of Infectious Diseases, Imperial College London, London, United Kingdon
- *Moorine Sekkade*: National Tuberculosis and Leprosy Program, Kampala, Uganda
- *Sabine Verkuijl*: Global Tuberculosis Programme, World Health Organization, Geneva, Switzerland
- Kerri Viney Global Tuberculosis Programme, World Health Organization, Geneva, Switzerland and School of Public Health, University of Sydney, Sydney, Australia
- *Bryan Vonasek*: Department of Pediatrics, University of Wisconsin School of Medicine and Public Health, Madison, United States of America

### Appendix F: Studies involved and data contributed to IPD.

**Figure S1.** Flow-diagram demonstrating how the eighteen studies that were identified as having potentially appropriate data for this analysis led to inclusion of 4,718 IPD records from children <10 years with presumptive pulmonary TB (1,811 [38·3%] were found to have either bacteriologically-confirmed TB or unconfirmed TB). *Data from three studies were not considered in this analysis for reflecting a population of children on treatment for TB (n=1) and active case finding populations (n=2). TB – tuberculosis, IPD – individual participant data, PTB – pulmonary tuberculosis.

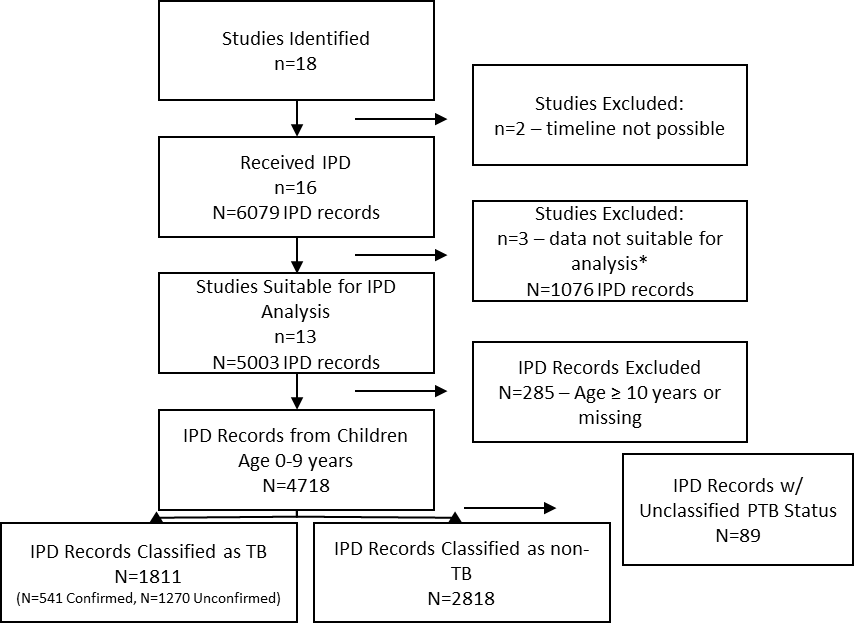

### Appendix G: Study information

**Table S2. Study information for Kabir/2020/BD**

| **Geographic setting** | Dhaka, Bangladesh | | |
| --- | --- | --- | --- |
| **Representing co-authors (in no particular order)** | Sayera Banu, Senjuti Kabir, Shakil Ahmed | | |
| **Healthcare setting** | **Name** | **Healthcare level** | **Recruitment setting** |
|  | Dhaka Medical College Hospital | Tertiary | Inpatient |
|  | Sir Salimullah Medical College and Mitford Hospital | Tertiary | Inpatient |
|  | Shaheed Suhrawardy Medical College and Hospital | Tertiary | Inpatient |
|  | icddr,b Dhaka Hospital | Tertiary | Inpatient |
| **Enrolment duration** | 22 January 2018 – 04 April 2019 | | |
| **Purpose for data collection** | Evaluate the performance of Xpert MTB/RIF Ultra assay on stool specimen for the diagnosis of childhood pulmonary TB | | |
| **Study design** | Cross-sectional study with follow-up of children diagnosed both bacteriologically and clinically at every month over phone started on antituberculosis therapy of 6 months | | |
| **Inclusion criteria** | Children aged 0-<15 years with symptoms suggestive of pulmonary TB based on any of the following: persistent non-remitting cough for >14 days weeks not responding to antibiotics, persistent documented fever for >14 days, document weight loss or failure to gain weight over the preceding 3 months, or fatigue/reduced playfulness/decreased activity | | |
| **Exclusion criteria** | Children with serious co-morbid condition (e.g., in intensive care unit, co-morbid heart condition, etc.); physician unable to collect respiratory specimen; children started on anti-tuberculosis treatment; children suspected clinically to have intestinal TB | | |
| **Standardized form to guide evaluation?** | YES | | |
| **Standardized form to guide CXR evaluation?** | NO: local treating physicians evaluated CXR | | |
| **Standardized follow-up for all children?** | NO: only children diagnosed w/ TB and initiated on anti-tuberculosis treatment were followed-up over phone to assess symptom resolution. Moreover, five children with “trace call” results on Xpert Ultra using stool were not advised for anti-TB treatment by treating physicians and were followed-up over phone at six months to assess if they had developed TB. | | |
| **Reference classification of TB** | Confirmed PTB – bacteriologically positive on culture/Xpert/Xpert Ultra on either induced sputum or stool specimens;  Unconfirmed PTB – bacteriologically negative but diagnosed by managing clinical team based on symptoms, CXR, TST, and contact history;  Unlikely PTB – not meeting criteria for confirmed TB or unconfirmed TB;  Classifications made by study team (separate from managing clinical team) based on data from the initial evaluation | | |
| **No. screened/No. enrolled** | 454/447 | | |
| **Ethics review** | Protocol no.PR#17072, Approved from Institutional Review Board, icddr,b constitutes of two Committee Research Review Committee on RRC on 18 July 2017 and Ethical Review Committee on 28 August 2017 | | |
| **Reference** | ^6^ | | |

**Table S3. Study information for Aurilio/2020/BR**

| **Geographic setting** | Rio de Janeiro, Brazil |
| --- | --- |

| **Representing co-authors (in no particular order)** | Clemax Couto Sant’Anna, Rafaela Baroni Aurilio |
| --- | --- |

| **Healthcare setting** | **Name** | **Healthcare level** | **Recruitment setting** |
| --- | --- | --- | --- |
|  | Instituto de Puericultura e Pediatria Martagao Gesteira | Tertiary | Inpatient/Outpatient |
|  | Hospital Raphael de Paula Souza | Tertiary | Inpatient/Outpatient |
|  | Hospital Universitário Antonio Pedro | Tertiary | Inpatient/Outpatient |
| **Enrolment duration** | 17 April 2014 – 27 July 2020 | | |
| **Purpose for data collection** | Evaluate Xpert MTB/RIF as diagnostic test for PTB in children | | |
| **Study design** | Prospective cohort study with baseline assessment and follow-up of all children at 60 days. Assessment of treatment outcome at 6 months or upon completion of treatment for those started on anti-TB treatment. | | |
| **Inclusion criteria** | Children aged 0-19 years with symptoms of respiratory infection for ≥14 days and abnormal CR | | |
| **Exclusion criteria** | Inappropriate samples for Xpert | | |
| **Standardized form to guide evaluation?** | YES | | |
| **Standardized form to guide CXR evaluation?** | YES | | |
| **Standardized follow-up for all children?** | YES | | |
| **Reference classification of TB** | Graham 2015: Confirmed PTB, Unconfirmed PTB, Unlikely PTB;^7^  Retrospective classifications made by study team (separate from managing clinical team) at the 2-month follow-up visit | | |
| **No. screened/No. enrolled** | 50/50 (among those children <10 years old) | | |
| **Ethics review** | Instituto de Puericultura e Pediatria Martagão Gesteira (24/02/2015, number 961·452 and 07/11/2017 number 2·369·814) and Hospital Universitário Antônio Pedro (28/07/2015 number 1·160·695) | | |
| **Reference** | ^8^ | | |

**Table S4. Study information for Song/2021/KE**

| **Geographic setting** | Kisumu County, Kenya |
| --- | --- |

| **Representing co-authors (in no particular order)** | Jonathan P. Smith, Eleanor S. Click, Rinn Song, Kevin P. Cain |
| --- | --- |

| **Healthcare setting** | **Name** | **Healthcare level** | **Recruitment setting** |
| --- | --- | --- | --- |
|  | Jaramogi Oginga Odinga Teaching and Referral Hospital | Tertiary | Inpatient/Outpatient |
|  | Additional patients from unspecified secondary inpatient/outpatient, and contact tracing | | |
| **Enrolment duration** | October 2013 – August 2015 | | |
| **Purpose for data collection** | Determine the performance of a wide panel of specimen types and microbiological tests in children evaluated for TB | | |
| **Study design** | Prospective cohort study with baseline assessment and follow-up of all children at 2 weeks, 2 months, and 6 months. | | |
| **Inclusion criteria** | Children aged <5 years, who weighed >2·5 kg with 1) cervical lymphadenopathy despite antibiotics or 2) parenchymal abnormality on chest radiograph in addition to at least one of the following symptoms: 1) cough despite antibiotics, 2) malnutrition despite treatment for malnutrition, and 3) fever despite antibiotics or antimalarials. | | |
| **Exclusion criteria** | Currently on anti-tuberculosis treatment or isoniazid preventive therapy or history of anti-tuberculosis treatment or isoniazid preventive therapy in the 6 months prior to enrolment. | | |
| **Standardized form to guide evaluation?** | YES | | |
| **Standardized form to guide CXR evaluation?** | YES | | |
| **Standardized follow-up for all children?** | YES | | |
| **Reference classification of TB** | Graham 2015: Confirmed PTB, Unconfirmed PTB, Unlikely PTB;^7^  Retrospective classifications made by study team (separate from managing clinical team) using information from all visits up to the 2-month follow-up | | |
| **No. screened/No. enrolled** | 2564/300 | | |
| **Ethics review** | US Centers for Disease Control and Prevention (#6334)  Kenya Medical Research Institute (#2343)  Jaramogi Oginga Odinga Teaching and Referral Hospital  Children’s Hospital Boston/Harvard Medical School (relied on the review and oversight of the Centers for Disease Control and Prevention institutional review board) | | |
| **Reference** | ^9^ | | |

**Table S5. Study information for LopezVarela/2015/MZ**

| **Geographic setting** | Manhiça District, Mozambique |
| --- | --- |

| **Representing co-authors (in no particular order)** | Elisa Lopez-Varela, Orvalho Augusto |
| --- | --- |

| **Healthcare setting** | **Name** | **Healthcare level** | **Recruitment setting** |
| --- | --- | --- | --- |
|  | Manhiça District Hospital (and attached health centre) | Secondary | Inpatient/Outpatient |
|  | *1483 patients from Manhiça District Hospital and Manhiça Health Research Centre Health and Demographic Surveillance System peripheral health centres (Palmeira, Maragra, Ilha, Josina, Taninga) and 180 contacts through contact tracing | | |
| **Enrolment duration** | 2011 - 2012 | | |
| **Purpose for data collection** | Estimate the annual minimum incidence of TB in children <3 in the Manhiça District | | |
| **Study design** | Prospective cohort study with baseline assessment and follow-up of all children within 6 months of enrolment. Persistently symptomatic children had additional evaluation and testing. | | |
| **Inclusion criteria** | Children aged 0-3 years with symptoms suggestive of PTB or EPTB or who are close contacts of notified TB cases. Symptoms suggestive of PTB included one or more of the following: cough ≥14 days not responding to appropriate antibiotics, fever ≥14 days after excluding malaria/pneumonia, chronic or acute malnutrition or failure to gain weight for more than 2 months, unexplained wheeze ≥14 months not responding to treatment, lower respiratory tract infection ≥14 days not responding to antibiotics after 72 hours, contact with TB case in previous 12 months) | | |
| **Exclusion criteria** | Children aged >3 who reside outside the study area or with diagnosis of TB at pre-enrolment | | |
| **Standardized form to guide evaluation?** | YES | | |
| **Standardized form to guide CXR evaluation?** | YES | | |
| **Standardized follow-up for all children?** | NO | | |
| **Reference classification of TB** | Graham 2012: Confirmed PTB, Probable PTB, Possible PTB, PTB Unlikely, MTB infection;^10^  Classification made by the managing clinical team using information from the baseline visit and all available follow-up visits | | |
| **No. screened/No. enrolled** | 1663/789 | | |
| **Ethics review** | The study protocol was approved by the Mozambican National Bioethics Committee and the Hospital Clinic of Barcelona Ethics Review Committee. | | |
| **Reference** | ^11^ | | |

**Table S6. Study information for Valencia/2017/MZ**

| **Geographic setting** | Manhiça District, Mozambique |
| --- | --- |

| **Representing co-authors (in no particular order)** | Alberto L. García-Basteiro, Lucía Carratalá-Castro |
| --- | --- |

| **Healthcare setting** | **Name** | **Healthcare level** | **Recruitment setting** |
| --- | --- | --- | --- |
|  | Manhiça District Hospital (and attached health centre) | Secondary | Inpatient/Outpatient |
|  | *Manhiça Health Care Centre and Manhiça District Hospital and 9 peripheral health care centre (Maluana, Munguine, Taninga, Maragra, Malavela, Palmeiras, Chibututuine, Calanga, Chibucutzo) | | |
| **Enrolment duration** | 20 August 2013 – 20 August 2014 | | |
| **Purpose for data collection** | Improve the quality of TB surveillance indicators using newly introduced Xpert MTB/RIF | | |
| **Study design** | Cross-sectional study with baseline assessment. Digital CXR only if clinician ordered. Two-week follow-up for children not initially started on TB treatment. Follow-up of all children started on TB treatment at months 2 and 6. | | |
| **Inclusion criteria** | Children and adults with symptoms suggestive of PTB or EPTB or who are close contacts of notified TB cases. Symptoms suggestive of PTB include cough ≥2 weeks, night sweats, weight loss, fever, and/or haemoptysis. | | |
| **Exclusion criteria** | Children diagnosed with TB prior to enrolment. | | |
| **Standardized form to guide evaluation?** | YES | | |
| **Standardized form to guide CXR evaluation?** | YES | | |
| **Standardized follow-up for all children?** | NO | | |
| **Reference classification of TB** | Confirmed PTB – Bacteriologically positive on culture/Xpert on any respiratory specimen;  Unconfirmed PTB - bacteriologically negative but diagnosed by managing clinical team based on symptoms, CXR, TST, and contact history;  Unlikely PTB - not meeting criteria for confirmed TB or unconfirmed TB;  Classification made by the managing clinical team using information from the initial visit and information gathered on follow-up at 2-weeks for those not started on TB treatment and after first few weeks of follow-up for those started on TB treatment | | |
| **No. screened/No. enrolled** | NA/142 | | |
| **Ethics review** | The study was approved by CISM local bioethics committee (CIBS) and the National Bioethics Committee (CNBS). Ref. 199/CNBS13 | | |
| **Reference** | ^12^ | | |

**Table S7. Study information for Myo/2018/MM**

| **Geographic setting** | Mandalay, Myanmar |
| --- | --- |

| **Representing co-authors (in no particular order)** | Aye Aye Myint, Kyaw Myo |
| --- | --- |

| **Healthcare setting** | **Name** | **Healthcare level** | **Recruitment setting** |
| --- | --- | --- | --- |
|  | Children Hospital, Mandalay | Tertiary | Inpatient and Casualty/Emergency |
| **Enrolment duration** | 01 January 2015 – 21 March 2017 | | |
| **Purpose for data collection** | Evaluate Xpert MTB/RIF as diagnostic test for PTB in children | | |
| **Study design** | Prospective cohort study with baseline assessment and 8-week follow-up. | | |
| **Inclusion criteria** | Children aged 0-12 years with cough ≥14 days and one of the following: fever >7 days, weight loss or failure to thrive, unexplained loss of appetite, or lethargy. | | |
| **Exclusion criteria** | Receipt of anti-tuberculosis treatment for >72 hours before specimen collection. | | |
| **Standardized form to guide evaluation?** | YES | | |
| **Standardized form to guide CXR evaluation?** | YES | | |
| **Standardized follow-up for all children?** | YES | | |
| **Reference classification of TB** | Graham 2015: Confirmed PTB, Unconfirmed PTB, Unlikely PTB;  Retrospective classifications made by study team (separate from managing clinical team) at the 2-month follow-up visit | | |
| **No. screened/No. enrolled** | 259/255 | | |
| **Ethics review** | Research Ethics Committee, University of Medicine, Mandalay | | |
| **Reference** | ^13^ | | |

**Table S8. Study information for Marcy/2019/Multi**

| **Geographic setting** | Bobo Dioulasso, Burkina Faso  Phnom Penh, Cambodia  Siem Reap, Cambodia  Yaounde, Cameroon  Ho Chi Minh City, Vietnam |
| --- | --- |

| **Representing co-authors (in no particular order)** | Olivier Marcy, Vibol Ung |
| --- | --- |

| **Healthcare setting** | **Name** | **Healthcare level** | **Recruitment setting** |
| --- | --- | --- | --- |
|  | Pediatric Department, Centre Hospitalier Universitaire Souro Sanou, (Bobo Dioulasso, Burkina Faso) | Tertiary | Inpatient/Outpatient |
|  | National Pediatric Hospital (Phnom Penh, Cambodia) | Tertiary | Inpatient/Outpatient |
|  | Angkor Hospital for Children (Siem Reap, Cambodia) | Tertiary | Inpatient/Outpatient |
|  | Centre Hospitalier de la Caisse d’Essos (Yaounde, Cameroon) | Secondary | Inpatient/Outpatient |
|  | Centre Mère et Enfant de la Fondation Chantal Biya (Yaounde, Cameroon) | Secondary | Inpatient/Outpatient |
|  | Pediatric Department, Pham Ngoc Thach Hospital (Ho Chi Minh City, Vietnam) | Tertiary | Inpatient/Outpatient |
|  | Infectious Diseases Department, Pediatric Hospital No. 1 (Nhi Dong 1) (Ho Chi Minh City, Vietnam) | Tertiary | Inpatient/Outpatient |
|  | Infectious Diseases Department, Pediatric Hospital No. 2 (Nhi Dong 2) (Ho Chi Minh City, Vietnam) | Tertiary | Inpatient/Outpatient |
| **Enrolment duration** | April 2011 – December 2014 | | |
| **Purpose for data collection** | Evaluate Xpert MTB/RIF performed on stool for Mtb and to assess response to antituberculosis treatment for children living with HIV | | |
| **Study design** | Prospective cohort study with baseline assessment and follow-up of all children at months 1,2,3, and 6. | | |
| **Inclusion criteria** | Children aged 0-12 years with HIV-1 infection (irrespective of HAART) and one or more of the following: cough >14 days, fever > 14 days, failure to thrive (deviation from previous growth trajectory in previous 3 months or weight-for-age Z-score <-2), failure to improve on broad spectrum antibiotics for pulmonary infection, or CXR suggestive of PTB | | |
| **Exclusion criteria** | History of any anti-tuberculosis treatment in the 2 years prior to enrolment. | | |
| **Standardized form to guide evaluation?** | YES | | |
| **Standardized form to guide CXR evaluation?** | YES | | |
| **Standardized follow-up for all children?** | YES | | |
| **Reference classification of TB** | Graham 2015: Confirmed PTB, Unconfirmed PTB, Unlikely PTB;  Retrospective classifications made by an algorithm following the Graham 2015 classification using information from all visits up to the 6-month follow-up visit | | |
| **No. screened/No. enrolled** | NA/438 | | |
| **Ethics review** | Ethics Committee for Research in Health (Burkina Faso);  National Ethics Comity for Health and Research (Phnom Penh,  Cambodia);  National Ethics Committee (Cameroon);  Division of Health Operations Research Ministry of Public Health (Cameroon);  Pham Ngoc Thach Hospital Institutional Review Board (Vietnam);  Ho Chi Minh City Department of Health (Vietnam);  Ho Chi Minh City People’s Committee (Vietnam). | | |
| **Reference** | ^14^ | | |

**Table S9. Study information for Hamid/2019/PK**

| **Geographic setting** | Karachi, Pakistan |
| --- | --- |

| **Representing co-authors (in no particular order)** | Iraj Batool, Aliya Anwar, Sara Ahmed Siddiqui, Farhana Amanullah |
| --- | --- |

| **Healthcare setting** | **Name** | **Healthcare level** | **Recruitment setting** |
| --- | --- | --- | --- |
|  | **The** Indus Hospital Ghauri Clinic | Tertiary/Referral | Outpatient |
|  | Participants were referred from the TB screening/ contact tracing program as well as from community general physicians, and family physicians/pediatricians/ surgeons of the Indus hospital. | | |
| **Enrolment duration** | 01 January 2019 – 06 April 2020 | | |
| **Purpose for data collection** | **To** identify gaps in child TB care delivery and improve Pediatric TB Program implementation | | |
| **Study design** | Cross-sectional study with baseline assessment and 1-month follow-up | | |
| **Inclusion criteria** | Children aged 0-10 years with any of the following: 2 symptoms of TB (cough ≥14 days, fever, weight loss, lethargy, loss of appetite, night sweats), a known TB exposure within the past 2 years with ≥1 symptom suggestive of TB, swollen lymph node for >14 days, previous history of TB and ≥1 symptom suggestive of TB | | |
| **Exclusion criteria** | N/A | | |
| **Standardized form to guide evaluation?** | YES | | |
| **Standardized form to guide CXR evaluation?** | YES | | |
| **Standardized follow-up for all children?** | YES | | |
| **Reference classification of TB** | Confirmed PTB – bacteriologically positive on culture/Xpert/ on any respiratory specimen (including stool);  Unconfirmed PTB – bacteriologically negative or not tested, but clinically diagnosed based on symptoms, CXR, +/-TST, and contact history;  Unlikely PTB – not meeting criteria for confirmed TB or unconfirmed TB;  Retrospective classification made by managing clinical teams at the 1-month follow-up visit | | |
| **No. screened/No. enrolled** | NA/447 | | |
| **Ethics review** | Data collection was approved by the Indus Hospital Research Center IRB. | | |
| **Reference** | ^15^ | | |

**Table S10. Study information for Zar/2019/ZA**

| **Geographic setting** | Cape Town, South Africa  Gqeberha South Africa |
| --- | --- |

| **Representing co-authors (in no particular order)** | Heather J. Zar, Mark P. Nicol |
| --- | --- |

| **Healthcare setting** | **Name** | **Healthcare level** | **Recruitment setting** |
| --- | --- | --- | --- |
|  | Red Cross War Memorial Children’s Hospital | Tertiary/Referral | Inpatient/Outpatient and Emergency/Casualty |
|  | Dora Nginza Provincial Hospital | Tertiary/Referral | Inpatient/Outpatient and Emergency/Casualty |
| **Enrolment duration** | 01 February 2010 – 31 January 2017 | | |
| **Purpose for data collection** | Novel tuberculosis diagnostics in HIV-infected and HIV-uninfected children. | | |
| **Study design** | Prospective cohort study with baseline assessment and follow-up of all children at months 1, 2, and 6. | | |
| **Inclusion criteria** | Children aged 0-15 years with clinical suspicion of PTB based on cough and one of (household TB contact within preceding 3 months, weight loss of failure to gain weight for preceding 3 months, positive TST, or chest radiograph suggestive of PTB) or clinical suspicion of EPTB | | |
| **Exclusion criteria** | Children who had received treatment for tuberculosis or TB prophylaxis for >72 hours prior to enrolment; patients living outside the catchment area; patients for whom adequate clinical samples could not be obtained; or patients for whom informed consent or permission for HIV testing could not be obtained | | |
| **Standardized form to guide evaluation?** | YES | | |
| **Standardized form to guide CXR evaluation?** | YES | | |
| **Standardized follow-up for all children?** | YES | | |
| **Reference classification of TB** | Graham 2015: Confirmed PTB, Unconfirmed PTB, Unlikely PTB;  Retrospective classifications made by study team (separate from managing clinical team) using data from all visits up to the 3-month follow-up visit | | |
| **No. screened/No. enrolled** | 4548/1346 | | |
| **Ethics review** | University of Cape Town Human Research Ethics Committee (HREC), 2008, Ref no. 045/2008. | | |
| **Reference** | ^16^ | | |

**Table S11. Study information for Walters/2017/ZA**

| **Geographic setting** | Cape Town, South Africa |
| --- | --- |

| **Representing co-authors (in no particular order)** | Anneke C. Hesseling, Marieke M. van der Zalm, Megan Palmer, Elisabetta G. Walters |
| --- | --- |

| **Healthcare setting** | **Name** | **Healthcare level** | **Recruitment setting** |
| --- | --- | --- | --- |
|  | Tygerberg Hospital | Tertiary/Referral | Inpatient/Outpatient |
|  | Karl Bremer Hospital | Secondary | Inpatient/Outpatient |
| **Enrolment duration** | March 2012 – November 2017 | | |
| **Purpose for data collection** | Evaluate feasible strategies to improve and promote microbiological testing of children with PTB and characterise treatment response. | | |
| **Study design** | Prospective cohort study with baseline assessment and follow-up of all children at months 1, 2, and 6. | | |
| **Inclusion criteria** | Children 0-12 with any of the following: cough ≥2 weeks, unexplained fever ≥1 week, poor growth/weight loss over the preceding 3 months, or cough <1 week with a known TB exposure in the previous 12 months, a positive TST, or a CXR suggestive of PTB | | |
| **Exclusion criteria** | Children who had received treatment for tuberculosis for >1 day or were being evaluated for EPTB without being evaluated for PTB. | | |
| **Standardized form to guide evaluation?** | YES | | |
| **Standardized form to guide CXR evaluation?** | YES | | |
| **Standardized follow-up for all children?** | YES | | |
| **Reference classification of TB** | Graham 2015: Confirmed PTB, Unconfirmed PTB, Unlikely PTB;  Retrospective classifications made by study team (separate from managing clinical team) using information from all visits up to the 2-month follow-up visit | | |
| **No. screened/No. enrolled** | NA/620 | | |
| **Ethics review** | Health Research Ethics Committee of Stellenbosch University Faculty of Health Sciences No. N11/09/282 | | |
| **Reference** | ^17^ | | |

**Table S12. Study information for Orikiriza/2018/UG**

| **Geographic setting** | Mbarara, Uganda |
| --- | --- |

| **Representing co-authors (in no particular order)** | Maryline Bonnet, Patrick Orikiriza |
| --- | --- |

| **Healthcare setting** | **Name** | **Healthcare level** | **Recruitment setting** |
| --- | --- | --- | --- |
|  | Mbarara Regional Referral Hospital | Tertiary/Referral | Inpatient/Outpatient (includes children referred from TB contact screening) |
| **Enrolment duration** | 12 April 2012 – 14 January 2014 | | |
| **Purpose for data collection** | Evaluate the performance of Xpert MTB/RIF on induced sputum and to assess treatment outcome and safety of pediatric TB drug dosages. | | |
| **Study design** | Prospective cohort study with baseline assessment and follow-up at 3 months for children not started on TB treatment and follow-up at 12 months for children started on TB treatment. | | |
| **Inclusion criteria** | Children aged 0-14 with any of: weight loss/failure to thrive/growth faltering over preceding 3 months, non-remittent cough or wheeze >14 days, night sweats in preceding 14 days, unexplained fever for ≥7 days, chest pain within the preceding 2 weeks, unexplained fatigue/weakness/apathy/lethargy in previous 2 weeks, or abnormal CXR suggestive of TB | | |
| **Exclusion criteria** | Children who had received >3 days of treatment for tuberculosis or had completed treatment within the past 6 months or with poor access to follow-up evaluation. | | |
| **Standardized form to guide evaluation?** | YES | | |
| **Standardized form to guide CXR evaluation?** | YES | | |
| **Standardized follow-up for all children?** | YES | | |
| **No. screened/No. enrolled** | 467/392 | | |
| **Reference classification of TB** | Graham 2012: Confirmed PTB, Probable PTB, Possible PTB, PTB Unlikely;  Retrospective classification made by blinded, independent endpoint review committee at 3-month visit for children not started on TB treatment and 6-month visit for children started on TB treatment | | |
| **Ethics review** | MUST Research Ethics Committee (MUST-REC), Uganda National Council for Science and Technology (UNCST), Comité de Protection des Personnes (CPP), Iles de France XI France. | | |
| **Reference** | ^18^ | | |

**Table S13. Study information for Orikiriza/2022/UG**

| **Geographic setting** | Mbarara, Uganda |
| --- | --- |

| **Representing co-authors (in no particular order)** | Maryline Bonnet, Dorah Nampijja |
| --- | --- |

| **Healthcare setting** | **Name** | **Healthcare level** | **Recruitment setting** |
| --- | --- | --- | --- |
|  | Mbarara Regional Referral Hospital | Tertiary/Referral | Inpatient |
| **Enrolment duration** | September 2015 – March 2018 | | |
| **Purpose for data collection** | Evaluate the performance of Xpert MTB/RIF on stool and urine AlereLAM among children with increased risk of disseminated or severe TB. | | |
| **Study design** | Prospective cohort study with baseline assessment and follow-up at weeks 1, 2, 8, and 24 for all children. | | |
| **Inclusion criteria** | Children aged 0-1 year or HIV-infected or with severe malnutrition and either 1) at least two of the following: cough >2 weeks, fever >1 week, severe malnutrition, >2 lethargy >2 weeks, known exposure to TB within preceding 2 years, or 2) any sign suggestive of TB meningitis or disseminated/miliary TB | | |
| **Exclusion criteria** | Children who received anti-tuberculosis treatment | | |
| **Standardized form to guide evaluation?** | YES | | |
| **Standardized form to guide CXR evaluation?** | YES | | |
| **Standardized follow-up for all children?** | YES | | |
| **Reference classification of TB** | Graham 2015: Confirmed PTB, Unconfirmed PTB, Unlikely PTB;  Automated diagnostic algorithm for retrospective classification using information from all visits up to the 6-month follow-up visit, review by independent endpoint committee for cases not classified by the algorithm | | |
| **No. screened/No. enrolled** | 238/219 | | |
| **Ethics review** | MUST Research Ethics Committee (MUST-REC), Uganda National Council for Science and Technology (UNCST), Comité de Protection des Personnes (CPP), Iles de France XI France. | | |
| **Reference** | ^19^ | | |

**Table S14. Study information for Giang/2015/VN**

| **Geographic setting** | Ho Chi Minh City, Vietnam |
| --- | --- |

| **Representing co-authors (in no particular order)** | Julie Huynh, Maxine Caws, Nguyen Thuy Thuong Thuong |
| --- | --- |

| **Healthcare setting** | **Name** | **Healthcare level** | **Recruitment setting** |
| --- | --- | --- | --- |
|  | Pham Ngoc Thach Hospital | Tertiary/Referral | Inpatient |
| **Enrolment duration** | 01 April 2013 – 01 October 2013 | | |
| **Purpose for data collection** | Evaluate the performance of Xpert MTB/RIF for the diagnosis of TB in HIV-uninfected children. | | |
| **Study design** | Prospective cohort study with baseline assessment and unspecified minimum follow-up (consistent with routine clinical practice). | | |
| **Inclusion criteria** | HIV-uninfected children aged 0-15 years with 1 or more of: persistent unexplained fever, cough >2 weeks, night sweats, weight loss, failure to thrive, reduced playfulness/lethargy, and/or any of the following for infants <60 days: neonatal pneumonia, unexplained hepatomegaly, or sepsis-like illness | | |
| **Exclusion criteria** | Children who received anti-tuberculosis treatment prior to specimen collection for Mtb confirmation or children living with HIV. | | |
| **Standardized form to guide evaluation?** | YES | | |
| **Standardized form to guide CXR evaluation?** | NO | | |
| **Standardized follow-up for all children?** | YES | | |
| **Reference classification of TB** | Graham 2012: Confirmed PTB, Probable PTB, Possible PTB, PTB Unlikely;  Retrospective classification made by the managing clinical team at 2-month visit | | |
| **No. screened/No. enrolled** | 154/150 | | |
| **Ethics review** | Pham Ngoc Thach Hospital Institutional review Board (IRB), the Oxford Tropical Ethics Committee (OxTREC) and the Health services of Ho Chi Minh City. | | |
| **Reference** | ^20^ | | |

### Appendix H: Modifications to IPD received

**Table S15. Modifications to IPD from Kabir/2020/BD**

| VARIABLE | DESCRIPTION | CODE | LABEL | MODIFICATION |
| --- | --- | --- | --- | --- |
| HIV status | Participant HIV status | 0 | HIV-negative | HIV status was not collected as a part of this study. In consultation with study authors, we assumed that all children in this study were HIV-negative given low HIV prevalence |
|  |  | 1 | HIV-positive |  |
|  |  | NA | Unknown |  |
| Documented TB exposure | Known exposure to MTB at initial evaluation in previous 12 months | 0 | No documented TB exposure in previous 12 months | An exposure was defined as having a family member living with the child who was diagnosed with and received treatment for TB in the previous 12 months. |
|  |  | 1 | Documented TB exposure in previous 12 months |  |
|  |  | NA | Unknown |  |
| First Xpert MTB/RIF | Result from first Xpert MTB/RIF (not Ultra) performed on ES/IS (or GA for young children) collected at initial evaluation | 0 | Xpert negative for Mtb | Result from first Xpert performed on induced sputum specimens. |
|  |  | 1 | Xpert positive for Mtb |  |
|  |  | NA | Unknown/not performed |  |
| CXR consistent with TB | Result of CXR performed at initial evaluation as assessed by reader performing clinical evaluation/making TB-treatment decision or by reader to inform research classification of TB if former not available | 0 | CXR not consistent with TB | CXR assessment made by the managing clinical team. |
|  |  | 1 | CXR consistent with TB |  |
|  |  | NA | Unknown/not assessed |  |
| TB classification | Final classification of TB | 0 | Unlikely TB | See note on reference classification in study description table above. |
|  |  | 1 | Bacteriologically-confirmed TB |  |
|  |  | 2 | Unconfirmed TB |  |
|  |  | NA | Unknown |  |

**Table S16. Modifications to IPD from Aurilio/2020/BR**

| VARIABLE | DESCRIPTION | CODE | LABEL | MODIFICATION |
| --- | --- | --- | --- | --- |
| Documented TB exposure | Known exposure to MTB at initial evaluation in previous 12 months | 0 | No documented TB exposure in previous 12 months | An exposure was defined as a mother, household member, or someone spending ~4 hours a day with the child having documented or reported positive Xpert or TB culture [or receiving treatment for TB] in the previous 12 months). |
|  |  | 1 | Documented TB exposure in previous 12 months |  |
|  |  | NA | Unknown |  |
| First Xpert MTB/RIF | Result from first Xpert MTB/RIF (not Ultra) performed on ES/IS (or GA for young children) collected at initial evaluation | 0 | Xpert negative for Mtb | Result from first Xpert: mostly GA/ES/IS, some pleural effusion, bronchoalveolar lavage, and tracheal aspirate. |
|  |  | 1 | Xpert positive for Mtb |  |
|  |  | NA | Unknown/not performed |  |
| CXR consistent with TB | Result of CXR performed at initial evaluation as assessed by reader performing clinical evaluation/making TB-treatment decision or by reader to inform research classification of TB if former not available | 0 | CXR not consistent with TB | All CXR assessments made by the study team. |
|  |  | 1 | CXR consistent with TB |  |
|  |  | NA | Unknown/not assessed |  |
| Opacities on CXR | Opacities (e.g., alveolar consolidation and/or bronchopneumonia) on CXR performed at initial evaluation as assessed by reader performing clinical evaluation/making TB-treatment decision or by reader to inform research classification of TB if former not available | 0 | Opacities not present on CXR | Received data corresponding to presence of alveolar opacification and bronchopneumonia; if either of these were positive, then the CXR was said to demonstrate opacities. |
|  |  | 1 | Opacities present on CXR |  |
|  |  | NA | Unknown/not assessed |  |
| Intrathoracic lymphadenopathy on CXR | Intrathoracic lymphadenopathy (e.g., perihilar nodes, paratracheal nodes, mediastinal nodes) on CXR performed at initial evaluation as assessed by reader performing clinical evaluation/making TB-treatment decision or by reader to inform research classification of TB if former not available | 0 | Intrathoracic lymphadenopathy not present on CXR | Received data corresponding to presence of perihilar lymphadenopathy, paratracheal lymphadenopathy, and calcified nodes; if any of these were positive, then the CXR was said to demonstrate nodes. |
|  |  | 1 | Intrathoracic lymphadenopathy present on CXR |  |
|  |  | NA | Unknown/not assessed |  |
| TB classification | Final classification of TB | 0 | Unlikely TB | See note on reference classification in study description table above. |
|  |  | 1 | Bacteriologically-confirmed TB |  |
|  |  | 2 | Unconfirmed TB |  |
|  |  | NA | Unknown |  |

**Table S17. Modifications to IPD from Song/2021/KE**

| VARIABLE | DESCRIPTION | CODE | LABEL | MODIFICATION |
| --- | --- | --- | --- | --- |
| Documented TB exposure | Known exposure to MTB at initial evaluation in previous 12 months | 0 | No documented TB exposure in previous 12 months | Tuberculosis exposure was defined as caregiver-reported household contact with someone with TB within 24 months prior to enrollment. |
|  |  | 1 | Documented TB exposure in previous 12 months |  |
|  |  | NA | Unknown |  |
| First Xpert MTB/RIF | Result from first Xpert MTB/RIF (not Ultra) performed on ES/IS (or GA for young children) collected at initial evaluation | 0 | Xpert negative for Mtb | Result from first Xpert: all GA specimens. |
|  |  | 1 | Xpert positive for Mtb |  |
|  |  | NA | Unknown/not performed |  |
| CXR consistent with TB | Result of CXR performed at initial evaluation as assessed by reader performing clinical evaluation/making TB-treatment decision or by reader to inform research classification of TB if former not available | 0 | CXR not consistent with TB | CXR assessment made by expert readers |
|  |  | 1 | CXR consistent with TB |  |
|  |  | NA | Unknown/not assessed |  |
| TB classification | Final classification of TB | 0 | Unlikely TB | See note on reference classification in study description table above. |
|  |  | 1 | Bacteriologically-confirmed TB |  |
|  |  | 2 | Unconfirmed TB |  |
|  |  | NA | Unknown |  |

**Table S18. Modifications to IPD from LopezVarela/2015/MZ**

| VARIABLE | DESCRIPTION | CODE | LABEL | MODIFICATION |
| --- | --- | --- | --- | --- |
| Weight loss | Presenting history of poor growth over the preceding 3 months AND not responding to nutritional rehabilitation (or antiretroviral therapy if HIV infected) | 0 | No weight loss | Used WAZ/HAZ for this definition. |
|  |  | 1 | Weight loss |  |
|  |  | NA | Unknown |  |
| Documented TB exposure | Known exposure to MTB at initial evaluation in previous 12 months | 0 | No documented TB exposure in previous 12 months | Exposure to TB was defined as contact with someone diagnosed as or being treated for TB. No time-limit, but given that all kids were under the age of 3, this would have included any exposure during lifetime. For those children identified through active case finding, the definition was contact with a smear-positive adult with PTB registered at the district National TB Program (NTP) in the previous 24 months. |
|  |  | 1 | Documented TB exposure in previous 12 months |  |
|  |  | NA | Unknown |  |
| Peripheral lymphadenopathy | Peripheral lymphadenopathy (at cervical, submandibular, and/or axillary nodes) at initial evaluation | 0 | No peripheral lymphadenopathy | Received data corresponding to presence of cervical lymphadenopathy and axillary lymphadenopathy; if either of these were positive, then the child was said to have peripheral lymphadenopathy. Not all children were assessed for this feature. |
|  |  | 1 | Peripheral lymphadenopathy |  |
|  |  | NA | Unknown |  |
| First Xpert MTB/RIF | Result from first Xpert MTB/RIF (not Ultra) performed on ES/IS (or GA for young children) collected at initial evaluation | 0 | Xpert negative for Mtb | Xpert was not performed as a part of this study. We assumed that a positive culture was equivalent to a positive Xpert result. Preferentially used the result from liquid culture or solid culture of the first GA specimen. If GA was not available, then we took the result of either liquid culture or solid culture from the first ES specimen. |
|  |  | 1 | Xpert positive for Mtb |  |
|  |  | NA | Unknown/not performed |  |
| CXR consistent with TB | Result of CXR performed at initial evaluation as assessed by reader performing clinical evaluation/making TB-treatment decision or by reader to inform research classification of TB if former not available | 0 | CXR not consistent with TB | All CXR assessments made by the study team, which was the same as the managing clinical team. |
|  |  | 1 | CXR consistent with TB |  |
|  |  | NA | Unknown/not assessed |  |
| Opacities on CXR | Opacities (e.g., alveolar consolidation and/or bronchopneumonia) on CXR performed at initial evaluation as assessed by reader performing clinical evaluation/making TB-treatment decision or by reader to inform research classification of TB if former not available | 0 | Opacities not present on CXR | Received data corresponding to presence of alveolar opacification; if this was positive, then the CXR was said to demonstrate opacities. |
|  |  | 1 | Opacities present on CXR |  |
|  |  | NA | Unknown/not assessed |  |
| TB classification | Final classification of TB | 0 | Unlikely TB | See note on reference classification in study description table above. 'Probable TB' and 'possible TB' were coded as 'unconfirmed TB,' 'MTB infection' was coded as 'unlikely TB.' |
|  |  | 1 | Bacteriologically-confirmed TB |  |
|  |  | 2 | Unconfirmed TB |  |
|  |  | NA | Unknown |  |

**Table S19. Modifications to IPD from Valencia/2017/MZ**

| VARIABLE | DESCRIPTION | CODE | LABEL | MODIFICATION |
| --- | --- | --- | --- | --- |
| Documented TB exposure | Known exposure to MTB at initial evaluation in previous 12 months | 0 | No documented TB exposure in previous 12 months | An exposure was defined as a mother, household member, or someone spending ~4 hours a day with the child receiving treatment for TB in the previous 12 months. |
|  |  | 1 | Documented TB exposure in previous 12 months |  |
|  |  | NA | Unknown |  |
| Peripheral lymphadenopathy | Peripheral lymphadenopathy (at cervical, submandibular, and/or axillary nodes) at initial evaluation | 0 | No peripheral lymphadenopathy | Received data corresponding to presence of cervical lymphadenopathy; if if this was positive, then the child was said to have peripheral lymphadenopathy. |
|  |  | 1 | Peripheral lymphadenopathy |  |
|  |  | NA | Unknown |  |
| First Xpert MTB/RIF | Result from first Xpert MTB/RIF (not Ultra) performed on ES/IS (or GA for young children) collected at initial evaluation | 0 | Xpert negative for Mtb | Result from first Xpert: all either ES or IS specimens. |
|  |  | 1 | Xpert positive for Mtb |  |
|  |  | NA | Unknown/not performed |  |
| CXR consistent with TB | Result of CXR performed at initial evaluation as assessed by reader performing clinical evaluation/making TB-treatment decision or by reader to inform research classification of TB if former not available | 0 | CXR not consistent with TB | All CXR assessments made by the managing clinical team. CXR was only performed for children for whom the managing clinical team determined that CXR was necessary; thus, not all children had CXR performed. |
|  |  | 1 | CXR consistent with TB |  |
|  |  | NA | Unknown/not assessed |  |
| Opacities on CXR | Opacities (e.g., alveolar consolidation and/or bronchopneumonia) on CXR performed at initial evaluation as assessed by reader performing clinical evaluation/making TB-treatment decision or by reader to inform research classification of TB if former not available | 0 | Opacities not present on CXR | Received data corresponding to presence of alveolar opacification and bronchopneumonia; if either of these were positive, then the CXR was said to demonstrate opacities. |
|  |  | 1 | Opacities present on CXR |  |
|  |  | NA | Unknown/not assessed |  |
| Intrathoracic lymphadenopathy on CXR | Intrathoracic lymphadenopathy (e.g., perihilar nodes, paratracheal nodes, mediastinal nodes) on CXR performed at initial evaluation as assessed by reader performing clinical evaluation/making TB-treatment decision or by reader to inform research classification of TB if former not available | 0 | Intrathoracic lymphadenopathy not present on CXR | Received data corresponding to presence of perihilar lymphadenopathy, paratracheal lymphadenopathy, and calcified nodes; if any of these were positive, then the CXR was said to demonstrate nodes. |
|  |  | 1 | Intrathoracic lymphadenopathy present on CXR |  |
|  |  | NA | Unknown/not assessed |  |
| TB classification | Final classification of TB | 0 | Unlikely TB | See note on reference classification in study description table above. |
|  |  | 1 | Bacteriologically-confirmed TB |  |
|  |  | 2 | Unconfirmed TB |  |
|  |  | NA | Unknown |  |

**Table S20. Modifications to IPD from Myo/2018/MM**

| VARIABLE | DESCRIPTION | CODE | LABEL | MODIFICATION |
| --- | --- | --- | --- | --- |
| Documented TB exposure | Known exposure to MTB at initial evaluation in previous 12 months | 0 | No documented TB exposure in previous 12 months | Defined as a documented or reported exposure to a case of tuberculosis (household or close contact) within the preceding 12 months |
|  |  | 1 | Documented TB exposure in previous 12 months |  |
|  |  | NA | Unknown |  |
| First Xpert MTB/RIF | Result from first Xpert MTB/RIF (not Ultra) performed on ES/IS (or GA for young children) collected at initial evaluation | 0 | Xpert negative for Mtb | Result from first Xpert: all GA specimens. |
|  |  | 1 | Xpert positive for Mtb |  |
|  |  | NA | Unknown/not performed |  |
| CXR consistent with TB | Result of CXR performed at initial evaluation as assessed by reader performing clinical evaluation/making TB-treatment decision or by reader to inform research classification of TB if former not available | 0 | CXR not consistent with TB | CXR assessment made by the study team. |
|  |  | 1 | CXR consistent with TB |  |
|  |  | NA | Unknown/not assessed |  |
| TB classification | Final classification of TB | 0 | Unlikely TB | See note on reference classification in study description table above. |
|  |  | 1 | Bacteriologically-confirmed TB |  |
|  |  | 2 | Unconfirmed TB |  |
|  |  | NA | Unknown |  |

**Table S21. Modifications to IPD from Marcy/2019/Multi**

| VARIABLE | DESCRIPTION | CODE | LABEL | MODIFICATION |
| --- | --- | --- | --- | --- |
| Fever duration | Presence of fever 1 week at initial evaluation | 0 | Fever 1 week not present | Fever duration was not provided in granular enough detail to identify those with fever for greater than or equal to one week. |
|  |  | 1 | Fever 1 week present |  |
|  |  | NA | Unknown |  |
| Lethargy | Presenting history of unusual lethargy or lack of playfulness at initial evaluation | 0 | No lethargy | Positive if the patient experienced lethargy in the previous 4 weeks. |
|  |  | 1 | Lethargy |  |
|  |  | NA | Unknown |  |
| Weight loss | Presenting history of poor growth over the preceding 3 months AND not responding to nutritional rehabilitation (or antiretroviral therapy if HIV infected) | 0 | No weight loss | Positive if the patient experienced weight loss in the previous 4 weeks. |
|  |  | 1 | Weight loss |  |
|  |  | NA | Unknown |  |
| Documented TB exposure | Known exposure to MTB at initial evaluation in previous 12 months | 0 | No documented TB exposure in previous 12 months | Exposure defined as having a household contact with smear + TB in the previous 12 months. |
|  |  | 1 | Documented TB exposure in previous 12 months |  |
|  |  | NA | Unknown |  |
| Night sweats | Presenting history of night sweats at initial evaluation | 0 | No night sweats | Positive if the patient experienced night sweats in the previous 4 weeks. |
|  |  | 1 | Night sweats |  |
|  |  | NA | Unknown |  |
| Haemoptysis | Presenting history of haemoptysis at initial evaluation | 0 | No haemoptysis | Positive if the patient experienced haemoptysis in the previous 4 weeks. |
|  |  | 1 | Haemoptysis |  |
|  |  | NA | Unknown |  |
| Peripheral lymphadenopathy | Peripheral lymphadenopathy (at cervical, submandibular, and/or axillary nodes) at initial evaluation | 0 | No peripheral lymphadenopathy | Received data corresponding to presence of cervical lymphadenopathy, submandibular lymphadenopathy, and axillary lymphadenopathy; if any of these were positive, then the child was said to have peripheral lymphadenopathy. |
|  |  | 1 | Peripheral lymphadenopathy |  |
|  |  | NA | Unknown |  |
| First Xpert MTB/RIF | Result from first Xpert MTB/RIF (not Ultra) performed on ES/IS (or GA for young children) collected at initial evaluation | 0 | Xpert negative for Mtb | Result from first Xpert: mostly ES, with some IS and GA specimens. |
|  |  | 1 | Xpert positive for Mtb |  |
|  |  | NA | Unknown/not performed |  |
| CXR consistent with TB | Result of CXR performed at initial evaluation as assessed by reader performing clinical evaluation/making TB-treatment decision or by reader to inform research classification of TB if former not available | 0 | CXR not consistent with TB | All CXR assessments made by the managing clinical team. |
|  |  | 1 | CXR consistent with TB |  |
|  |  | NA | Unknown/not assessed |  |
| Opacities on CXR | Opacities (e.g., alveolar consolidation and/or bronchopneumonia) on CXR performed at initial evaluation as assessed by reader performing clinical evaluation/making TB-treatment decision or by reader to inform research classification of TB if former not available | 0 | Opacities not present on CXR | Received data corresponding to presence of alveolar opacification; if this was positive, then the CXR was said to demonstrate opacities. |
|  |  | 1 | Opacities present on CXR |  |
|  |  | NA | Unknown/not assessed |  |
| Intrathoracic lymphadenopathy on CXR | Intrathoracic lymphadenopathy (e.g., perihilar nodes, paratracheal nodes, mediastinal nodes) on CXR performed at initial evaluation as assessed by reader performing clinical evaluation/making TB-treatment decision or by reader to inform research classification of TB if former not available | 0 | Intrathoracic lymphadenopathy not present on CXR | Received data corresponding to presence of perihilar lymphadenopathy and paratracheal lymphadenopathy; if either of these were positive, then the CXR was said to demonstrate nodes. |
|  |  | 1 | Intrathoracic lymphadenopathy present on CXR |  |
|  |  | NA | Unknown/not assessed |  |
| TB classification | Final classification of TB | 0 | Unlikely TB | See note on reference classification in study description table above. |
|  |  | 1 | Bacteriologically-confirmed TB |  |
|  |  | 2 | Unconfirmed TB |  |
|  |  | NA | Unknown |  |

**Table S22. Modifications to IPD from Hamid/2019/PK**

| VARIABLE | DESCRIPTION | CODE | LABEL | MODIFICATION |
| --- | --- | --- | --- | --- |
| Age (months) | Age (months) at enrolment | ### | NA = unknown | Age was reported as years old; assumed to be at midpoint of year and converted to months. |
| HIV status | Participant HIV status | 0 | HIV-negative | HIV status was not collected as a part of this study. In consultation with study authors, we assumed that all children in this study were HIV-negative. |
|  |  | 1 | HIV-positive |  |
|  |  | NA | Unknown |  |
| Weight loss | Presenting history of poor growth over the preceding 3 months AND not responding to nutritional rehabilitation (or antiretroviral therapy if HIV infected) | 0 | No weight loss | Defined as subjective weight loss reported by parents/guardians. |
|  |  | 1 | Weight loss |  |
|  |  | NA | Unknown |  |
| Documented TB exposure | Known exposure to MTB at initial evaluation in previous 12 months | 0 | No documented TB exposure in previous 12 months | An exposure was defined as a mother, household member, or someone spending ~4 hours a day with the child having documented or reported positive Xpert or TB culture (or receiving treatment for TB) in the previous 24 months. |
|  |  | 1 | Documented TB exposure in previous 12 months |  |
|  |  | NA | Unknown |  |
| First Xpert MTB/RIF | Result from first Xpert MTB/RIF (not Ultra) performed on ES/IS (or GA for young children) collected at initial evaluation | 0 | Xpert negative for Mtb | Result from first Xpert: performed only on stool specimens. Not all children received Xpert testing. |
|  |  | 1 | Xpert positive for Mtb |  |
|  |  | NA | Unknown/not performed |  |
| CXR consistent with TB | Result of CXR performed at initial evaluation as assessed by reader performing clinical evaluation/making TB-treatment decision or by reader to inform research classification of TB if former not available | 0 | CXR not consistent with TB | All CXR assessments made by the managing clinical team. |
|  |  | 1 | CXR consistent with TB |  |
|  |  | NA | Unknown/not assessed |  |
| TB classification | Final classification of TB | 0 | Unlikely TB | Eighteen children were given a diagnosis of EPTB; these children were removed from the analysis. Otherwise, see note on reference classification in study description table above. |
|  |  | 1 | Bacteriologically-confirmed TB |  |
|  |  | 2 | Unconfirmed TB |  |
|  |  | NA | Unknown |  |

**Table S23. Modifications to IPD from Zar/2019/ZA**

| VARIABLE | DESCRIPTION | CODE | LABEL | MODIFICATION |
| --- | --- | --- | --- | --- |
| Documented TB exposure | Known exposure to MTB at initial evaluation in previous 12 months | 0 | No documented TB exposure in previous 12 months | An exposure was defined as a mother, household member, or someone spending ~4 hours a day with the child having documented or reported positive Xpert or TB culture (or receiving treatment for TB) in the previous 24 months. |
|  |  | 1 | Documented TB exposure in previous 12 months |  |
|  |  | NA | Unknown |  |
| First Xpert MTB/RIF | Result from first Xpert MTB/RIF (not Ultra) performed on ES/IS (or GA for young children) collected at initial evaluation | 0 | Xpert negative for Mtb | Result from first Xpert: performed only on IS specimens. |
|  |  | 1 | Xpert positive for Mtb |  |
|  |  | NA | Unknown/not performed |  |
| CXR consistent with TB | Result of CXR performed at initial evaluation as assessed by reader performing clinical evaluation/making TB-treatment decision or by reader to inform research classification of TB if former not available | 0 | CXR not consistent with TB | All CXR assessments made by the study team; many were determined to be inconclusive for PTB. |
|  |  | 1 | CXR consistent with TB |  |
|  |  | NA | Unknown/not assessed |  |
| Opacities on CXR | Opacities (e.g., alveolar consolidation and/or bronchopneumonia) on CXR performed at initial evaluation as assessed by reader performing clinical evaluation/making TB-treatment decision or by reader to inform research classification of TB if former not available | 0 | Opacities not present on CXR | Received data corresponding to presence of alveolar opacification; if positive, then the CXR was said to demonstrate opacities. |
|  |  | 1 | Opacities present on CXR |  |
|  |  | NA | Unknown/not assessed |  |
| Intrathoracic lymphadenopathy on CXR | Intrathoracic lymphadenopathy (e.g., perihilar nodes, paratracheal nodes, mediastinal nodes) on CXR performed at initial evaluation as assessed by reader performing clinical evaluation/making TB-treatment decision or by reader to inform research classification of TB if former not available | 0 | Intrathoracic lymphadenopathy not present on CXR | Received data corresponding to presence of perihilar lymphadenopathy and paratracheal lymphadenopathy; if either of these were positive, then the CXR was said to demonstrate nodes. |
|  |  | 1 | Intrathoracic lymphadenopathy present on CXR |  |
|  |  | NA | Unknown/not assessed |  |
| TB classification | Final classification of TB | 0 | Unlikely TB | Removed data from 37 individuals with EPTB as not relevant to the analysis population. Otherwise, see note on reference classification in study description table above. |
|  |  | 1 | Bacteriologically-confirmed TB |  |
|  |  | 2 | Unconfirmed TB |  |
|  |  | NA | Unknown |  |

**Table S24. Modifications to IPD from Walters/2017/ZA**

| VARIABLE | DESCRIPTION | CODE | LABEL | MODIFICATION |
| --- | --- | --- | --- | --- |
| Weight loss | Presenting history of poor growth over the preceding 3 months AND not responding to nutritional rehabilitation (or antiretroviral therapy if HIV infected) | 0 | No weight loss | Weight loss was specifically defined as follows: Poor growth documented over the preceding 3 months (clear deviation from the child's previous growth trajectory and/or static growth or weight loss in the preceding 3 months; alternatively, weight-for-age Z-score (WFAZ) ≤2 in children with no previous weight measurements). |
|  |  | 1 | Weight loss |  |
|  |  | NA | Unknown |  |
| Documented TB exposure | Known exposure to MTB at initial evaluation in previous 12 months | 0 | No documented TB exposure in previous 12 months | Exposure to any identified adult TB source case in the preceding 12 months, where exposure was either within the household; or involved the child's primary caregiver; or occurred for >4 hours per day during the period of exposure. |
|  |  | 1 | Documented TB exposure in previous 12 months |  |
|  |  | NA | Unknown |  |
| Peripheral lymphadenopathy | Peripheral lymphadenopathy (at cervical, submandibular, and/or axillary nodes) at initial evaluation | 0 | No peripheral lymphadenopathy | Received data corresponding to presence of cervical lymphadenopathy, submandibular lymphadenopathy, and axillary lymphadenopathy; if any of these were positive, then the child was said to have peripheral lymphadenopathy. |
|  |  | 1 | Peripheral lymphadenopathy |  |
|  |  | NA | Unknown |  |
| First Xpert MTB/RIF | Result from first Xpert MTB/RIF (not Ultra) performed on ES/IS (or GA for young children) collected at initial evaluation | 0 | Xpert negative for Mtb | Result from first Xpert performed. |
|  |  | 1 | Xpert positive for Mtb |  |
|  |  | NA | Unknown/not performed |  |
| CXR consistent with TB | Result of CXR performed at initial evaluation as assessed by reader performing clinical evaluation/making TB-treatment decision or by reader to inform research classification of TB if former not available | 0 | CXR not consistent with TB | All CXR assessments made by the study team. |
|  |  | 1 | CXR consistent with TB |  |
|  |  | NA | Unknown/not assessed |  |
| Opacities on CXR | Opacities (e.g., alveolar consolidation and/or bronchopneumonia) on CXR performed at initial evaluation as assessed by reader performing clinical evaluation/making TB-treatment decision or by reader to inform research classification of TB if former not available | 0 | Opacities not present on CXR | Received data corresponding to presence of alveolar opacification and bronchopneumonia; if either of these were positive, then the CXR was said to demonstrate opacities. |
|  |  | 1 | Opacities present on CXR |  |
|  |  | NA | Unknown/not assessed |  |
| Intrathoracic lymphadenopathy on CXR | Intrathoracic lymphadenopathy (e.g., perihilar nodes, paratracheal nodes, mediastinal nodes) on CXR performed at initial evaluation as assessed by reader performing clinical evaluation/making TB-treatment decision or by reader to inform research classification of TB if former not available | 0 | Intrathoracic lymphadenopathy not present on CXR | Received data corresponding to presence of perihilar lymphadenopathy, paratracheal lymphadenopathy, and calcified nodes; if any of these were positive, then the CXR was said to demonstrate nodes. |
|  |  | 1 | Intrathoracic lymphadenopathy present on CXR |  |
|  |  | NA | Unknown/not assessed |  |
| TB classification | Final classification of TB | 0 | Unlikely TB | See note on reference classification in study description table above. |
|  |  | 1 | Bacteriologically-confirmed TB |  |
|  |  | 2 | Unconfirmed TB |  |
|  |  | NA | Unknown |  |

**Table S25. Modifications to IPD from Orikiriza/2018/UG**

| VARIABLE | DESCRIPTION | CODE | LABEL | MODIFICATION |
| --- | --- | --- | --- | --- |
| Cough duration | Duration of cough at initial evaluation | 0 | No cough | Duration of cough was only provided a greater than or equal to 2 weeks or less than 2 weeks/no cough. |
|  |  | 1 | Cough 0-13 days |  |
|  |  | 2 | Cough 14-20 days |  |
|  |  | 3 | Cough 21-27 days |  |
|  |  | 4 | Cough 28 days |  |
|  |  | NA | Unknown |  |
| Fever duration | Duration of fever at initial evaluation | 0 | No fever | Duration of fever was only provided as greater than or equal to 1 week or less than 1 week/no cough. |
|  |  | 1 | Fever 0-13 days |  |
|  |  | 2 | Fever 14-20 days |  |
|  |  | 3 | Fever 21-27 days |  |
|  |  | 4 | Fever 28 days |  |
|  |  | NA | Unknown |  |
| Lethargy | Presenting history of unusual lethargy or lack of playfulness at initial evaluation | 0 | No lethargy | Lethargy was positive if present for greater than or equal to 2 weeks; negative if no lethargy or for less than 2 weeks. |
|  |  | 1 | Lethargy |  |
|  |  | NA | Unknown |  |
| Documented TB exposure | Known exposure to MTB at initial evaluation in previous 12 months | 0 | No documented TB exposure in previous 12 months | For children referred from another contact study (any child who has lived in the same household with the index case continuously for at least 2 weeks within the 3-month period immediately preceding the diagnosis of smear-positive or culture-positive TB in the index case. For other children, documented as reported contact with a bacteriologically-positive case within the preceding 12 months. |
|  |  | 1 | Documented TB exposure in previous 12 months |  |
|  |  | NA | Unknown |  |
| Night sweats | Presenting history of night sweats at initial evaluation | 0 | No night sweats | Night sweats coded using the following scale: absent, mild, moderate, severe, or life threatening. Recoded absent = 0, and others = 1. |
|  |  | 1 | Night sweats |  |
|  |  | NA | Unknown |  |
| Peripheral lymphadenopathy | Peripheral lymphadenopathy (at cervical, submandibular, and/or axillary nodes) at initial evaluation | 0 | No peripheral lymphadenopathy | Location of peripheral lymphadenopathy not specified. |
|  |  | 1 | Peripheral lymphadenopathy |  |
|  |  | NA | Unknown |  |
| First Xpert MTB/RIF | Result from first Xpert MTB/RIF (not Ultra) performed on ES/IS (or GA for young children) collected at initial evaluation | 0 | Xpert negative for Mtb | Result from first Xpert: performed on two pooled IS specimens. |
|  |  | 1 | Xpert positive for Mtb |  |
|  |  | NA | Unknown/not performed |  |
| CXR consistent with TB | Result of CXR performed at initial evaluation as assessed by reader performing clinical evaluation/making TB-treatment decision or by reader to inform research classification of TB if former not available | 0 | CXR not consistent with TB | All CXR assessments made by the managing clinical team. |
|  |  | 1 | CXR consistent with TB |  |
|  |  | NA | Unknown/not assessed |  |
| Opacities on CXR | Opacities (e.g., alveolar consolidation and/or bronchopneumonia) on CXR performed at initial evaluation as assessed by reader performing clinical evaluation/making TB-treatment decision or by reader to inform research classification of TB if former not available | 0 | Opacities not present on CXR | Received data corresponding to presence of alveolar opacification and bronchopneumonia; if either of these were positive, then the CXR was said to demonstrate opacities. |
|  |  | 1 | Opacities present on CXR |  |
|  |  | NA | Unknown/not assessed |  |
| Intrathoracic lymphadenopathy on CXR | Intrathoracic lymphadenopathy (e.g., perihilar nodes, paratracheal nodes, mediastinal nodes) on CXR performed at initial evaluation as assessed by reader performing clinical evaluation/making TB-treatment decision or by reader to inform research classification of TB if former not available | 0 | Intrathoracic lymphadenopathy not present on CXR | Positive if mediastinal lymphadenopathy was present. |
|  |  | 1 | Intrathoracic lymphadenopathy present on CXR |  |
|  |  | NA | Unknown/not assessed |  |
| TB classification | Final classification of TB | 0 | Unlikely TB | See note on reference classification in study description table above. 'Probable TB' and 'possible TB' were coded as 'unconfirmed TB.' |
|  |  | 1 | Bacteriologically-confirmed TB |  |
|  |  | 2 | Unconfirmed TB |  |
|  |  | NA | Unknown |  |

**Table S26. Modifications to IPD from Orikiriza/2022/UG**

| VARIABLE | DESCRIPTION | CODE | LABEL | MODIFICATION |
| --- | --- | --- | --- | --- |
| BCG vaccination | Evidence of BGC vaccination (BCG scar or BCG recorded in immunization record) at initial evaluation | 0 | No evidence of BCG vaccination | If had a BCG-scar or a positive immunization card/verbal response, then determined to have evidence of BCG vaccination. |
|  |  | 1 | Evidence of BCG vaccination |  |
|  |  | NA | Unknown |  |
| Cough duration | Duration of cough at initial evaluation | 0 | No cough | Duration of cough was only provided a greater than or equal to 2 weeks or less than 2 weeks/no cough. |
|  |  | 1 | Cough 0-13 days |  |
|  |  | 2 | Cough 14-20 days |  |
|  |  | 3 | Cough 21-27 days |  |
|  |  | 4 | Cough 28 days |  |
|  |  | NA | Unknown |  |
| Fever duration | Duration of fever at initial evaluation | 0 | No fever | Duration of fever was only provided as greater than or equal to 1 week or less than 1 week/no cough. |
|  |  | 1 | Fever 0-13 days |  |
|  |  | 2 | Fever 14-20 days |  |
|  |  | 3 | Fever 21-27 days |  |
|  |  | 4 | Fever 28 days |  |
|  |  | NA | Unknown |  |
| Documented TB exposure | Known exposure to MTB at initial evaluation in previous 12 months | 0 | No documented TB exposure in previous 12 months | Contact of a household member with positive Xpert or TB culture in the previous 12 months |
|  |  | 1 | Documented TB exposure in previous 12 months |  |
|  |  | NA | Unknown |  |
| Peripheral lymphadenopathy | Peripheral lymphadenopathy (at cervical, submandibular, and/or axillary nodes) at initial evaluation | 0 | No peripheral lymphadenopathy | Significant peripheral lymphadenopathy on screening (location unspecified). |
|  |  | 1 | Peripheral lymphadenopathy |  |
|  |  | NA | Unknown |  |
| First Xpert MTB/RIF | Result from first Xpert MTB/RIF (not Ultra) performed on ES/IS (or GA for young children) collected at initial evaluation | 0 | Xpert negative for Mtb | Result from first Xpert: performed on GA specimens. |
|  |  | 1 | Xpert positive for Mtb |  |
|  |  | NA | Unknown/not performed |  |
| CXR consistent with TB | Result of CXR performed at initial evaluation as assessed by reader performing clinical evaluation/making TB-treatment decision or by reader to inform research classification of TB if former not available | 0 | CXR not consistent with TB | All CXR assessments made by the managing clinical team. |
|  |  | 1 | CXR consistent with TB |  |
|  |  | NA | Unknown/not assessed |  |
| Opacities on CXR | Opacities (e.g., alveolar consolidation and/or bronchopneumonia) on CXR performed at initial evaluation as assessed by reader performing clinical evaluation/making TB-treatment decision or by reader to inform research classification of TB if former not available | 0 | Opacities not present on CXR | Received data corresponding to presence of alveolar opacification and bronchopneumonia; if either of these were positive, then the CXR was said to demonstrate opacities. |
|  |  | 1 | Opacities present on CXR |  |
|  |  | NA | Unknown/not assessed |  |
| Intrathoracic lymphadenopathy on CXR | Intrathoracic lymphadenopathy (e.g., perihilar nodes, paratracheal nodes, mediastinal nodes) on CXR performed at initial evaluation as assessed by reader performing clinical evaluation/making TB-treatment decision or by reader to inform research classification of TB if former not available | 0 | Intrathoracic lymphadenopathy not present on CXR | Received data corresponding to presence of Gohn focus and hilar lymphadenopathy (grouped together) and mediastinal nodes; if either of these were positive, then the CXR was said to demonstrate nodes. |
|  |  | 1 | Intrathoracic lymphadenopathy present on CXR |  |
|  |  | NA | Unknown/not assessed |  |
| TB classification | Final classification of TB | 0 | Unlikely TB | See note on reference classification in study description table above. |
|  |  | 1 | Bacteriologically-confirmed TB |  |
|  |  | 2 | Unconfirmed TB |  |
|  |  | NA | Unknown |  |

**Table S27. Modifications to IPD from Giang/2015/VN**

| VARIABLE | DESCRIPTION | CODE | LABEL | MODIFICATION |
| --- | --- | --- | --- | --- |
| Cough duration | Duration of cough at initial evaluation | 0 | No cough | Duration of cough was only provided a greater than or equal to 2 weeks or less than 2 weeks/no cough. |
|  |  | 1 | Cough 0-13 days |  |
|  |  | 2 | Cough 14-20 days |  |
|  |  | 3 | Cough 21-27 days |  |
|  |  | 4 | Cough 28 days |  |
|  |  | NA | Unknown |  |
| Fever duration | Duration of fever at initial evaluation | 0 | No fever | Duration of fever was only provided as greater than or equal to 1 week or less than 1 week/no cough. |
|  |  | 1 | Fever 0-13 days |  |
|  |  | 2 | Fever 14-20 days |  |
|  |  | 3 | Fever 21-27 days |  |
|  |  | 4 | Fever 28 days |  |
|  |  | NA | Unknown |  |
| Weight loss | Presenting history of poor growth over the preceding 3 months AND not responding to nutritional rehabilitation (or antiretroviral therapy if HIV infected) | 0 | No weight loss | Subjective weight loss and/or failure to thrive. |
|  |  | 1 | Weight loss |  |
|  |  | NA | Unknown |  |
| Documented TB exposure | Known exposure to MTB at initial evaluation in previous 12 months | 0 | No documented TB exposure in previous 12 months | Exposure was defined as a household or close contact with a TB case (unspecified). |
|  |  | 1 | Documented TB exposure in previous 12 months |  |
|  |  | NA | Unknown |  |
| Peripheral lymphadenopathy | Peripheral lymphadenopathy (at cervical, submandibular, and/or axillary nodes) at initial evaluation | 0 | No peripheral lymphadenopathy | Received data corresponding to presence of cervical lymphadenopathy and submandibular lymphadenopathy; if either of these were positive, then the child was said to have peripheral lymphadenopathy. |
|  |  | 1 | Peripheral lymphadenopathy |  |
|  |  | NA | Unknown |  |
| First Xpert MTB/RIF | Result from first Xpert MTB/RIF (not Ultra) performed on ES/IS (or GA for young children) collected at initial evaluation | 0 | Xpert negative for Mtb | Result from first Xpert: performed on mostly GA specimens. |
|  |  | 1 | Xpert positive for Mtb |  |
|  |  | NA | Unknown/not performed |  |
| CXR consistent with TB | Result of CXR performed at initial evaluation as assessed by reader performing clinical evaluation/making TB-treatment decision or by reader to inform research classification of TB if former not available | 0 | CXR not consistent with TB | Unclear whether this data corresponds to result as assessed by the study team or the managing clinical team. |
|  |  | 1 | CXR consistent with TB |  |
|  |  | NA | Unknown/not assessed |  |
| TB classification | Final classification of TB | 0 | Unlikely TB | See note on reference classification in study description table above. 'Probable TB' and 'possible TB' were coded as 'unconfirmed TB.' One child unable to classify as 'probable TB' or 'possible TB' in original data was coded as 'unconfirmed TB' for the purposes of this analysis |
|  |  | 1 | Bacteriologically-confirmed TB |  |
|  |  | 2 | Unconfirmed TB |  |
|  |  | NA | Unknown |  |

### Appendix I: Descriptions of study-level IPD

All clinical, bacteriology, and imaging data collected from the initial visit. IPD – Individual participant data, TB – tuberculosis, IQR – interquartile range, BCG – bacille Calmette-Guerin, HIV – human immunodeficiency virus, Mtb – *Mycobacterium tuberculosis*, CXR – chest x-ray, TST – tuberculin skin test.

**Table S28. Description of IPD from Kabir/2020/BD**

| **Variable** | **Value** | **Confirmed TB (N=63)** | **Unconfirmed TB (N=36)** | **Unlikely TB (N=303)** |
| --- | --- | --- | --- | --- |
|  |  | n (%) OR median (IQR) | n (%) OR median (IQR) | n (%) OR median (IQR) |
| Age | Months | 25·6 (11,69·6) | 41·5 (7·4,83·6) | 18·5 (8,57·3) |
|  | Unknown | 0 (0) | 0 (0) | 0 (0) |
| Sex | Male | 36 (57) | 24 (67) | 177 (58) |
|  | Female | 27 (43) | 12 (33) | 126 (42) |
|  | Unknown | 0 (0) | 0 (0) | 0 (0) |
| Weight-for-age | z-score | -2.6 (-4.1,-1.5) | -2.7 (-3.7,-1.5) | -2.5 (-3.6,-1.3) |
|  | Unknown | 0 (0) | 0 (0) | 0 (0) |
| BCG vaccination | Evidence of BCG vaccination | 54 (86) | 32 (89) | 227 (75) |
|  | No evidence of BCG vaccination | 9 (14) | 4 (11) | 76 (25) |
|  | Unknown | 0 (0) | 0 (0) | 0 (0) |
| HIV status | HIV-positive | 0 (0) | 0 (0) | 0 (0) |
|  | HIV-negative | 63 (100) | 36 (100) | 303 (100) |
|  | Unknown | 0 (0) | 0 (0) | 0 (0) |
| Cough duration | No cough | 2 (3) | 1 (3) | 10 (3) |
|  | Cough 0-13 days | 18 (29) | 7 (19) | 71 (23) |
|  | Cough 14-20 days | 15 (24) | 5 (14) | 81 (27) |
|  | Cough 21-27 days | 4 (6) | 7 (19) | 31 (10) |
|  | Cough >27 days | 24 (38) | 16 (44) | 110 (36) |
|  | Unknown | 0 (0) | 0 (0) | 0 (0) |
| Cough ≥ 2 weeks | Cough ≥2 weeks present | 43 (68) | 28 (78) | 222 (73) |
|  | Cough ≥2 weeks not present | 20 (32) | 8 (22) | 81 (27) |
|  | Unknown | 0 (0) | 0 (0) | 0 (0) |
| Fever duration | No fever | 1 (2) | 2 (6) | 13 (4) |
|  | Fever 0-13 days | 14 (22) | 7 (19) | 94 (31) |
|  | Fever 14-20 days | 16 (25) | 10 (28) | 77 (25) |
|  | Fever 21-27 days | 7 (11) | 3 (8) | 26 (9) |
|  | Fever >27 days | 25 (40) | 14 (39) | 93 (31) |
|  | Unknown | 0 (0) | 0 (0) | 0 (0) |
| Fever ≥1 week | Fever ≥1 week present | 58 (92) | 32 (89) | 255 (84) |
|  | Fever ≥1 week not present | 5 (8) | 4 (11) | 48 (16) |
|  | Unknown | 0 (0) | 0 (0) | 0 (0) |
| Lethargy | Lethargy | 60 (95) | 33 (92) | 282 (93) |
|  | No lethargy | 3 (5) | 3 (8) | 21 (7) |
|  | Unknown | 0 (0) | 0 (0) | 0 (0) |
| Weight loss | Weight loss | 54 (86) | 33 (92) | 255 (84) |
|  | No weight loss | 9 (14) | 3 (8) | 48 (16) |
|  | Unknown | 0 (0) | 0 (0) | 0 (0) |
| Documented TB exposure | Documented TB exposure in previous 12 months | 17 (27) | 17 (47) | 65 (21) |
|  | No documented TB exposure in previous 12 months | 46 (73) | 19 (53) | 238 (79) |
|  | Unknown | 0 (0) | 0 (0) | 0 (0) |
| Night sweats | Night sweats | 42 (67) | 25 (69) | 179 (59) |
|  | No night sweats | 21 (33) | 11 (31) | 124 (41) |
|  | Unknown | 0 (0) | 0 (0) | 0 (0) |
| Haemoptysis | Haemoptysis | 0 (0) | 1 (3) | 7 (2) |
|  | No haemoptysis | 9 (14) | 6 (17) | 94 (31) |
|  | Unknown | 54 (86) | 29 (81) | 202 (67) |
| Objective fever (≥38 degrees Celsius) | Objective fever | 0 (0) | 0 (0) | 0 (0) |
|  | No objective fever | 0 (0) | 0 (0) | 0 (0) |
|  | Unknown | 63 (100) | 36 (100) | 303 (100) |
| Tachycardia | Tachycardia | 0 (0) | 0 (0) | 0 (0) |
|  | No tachycardia | 0 (0) | 0 (0) | 0 (0) |
|  | Unknown | 63 (100) | 36 (100) | 303 (100) |
| Tachypnoea | Tachypnoea | 0 (0) | 0 (0) | 0 (0) |
|  | No tachypnoea | 0 (0) | 0 (0) | 0 (0) |
|  | Unknown | 63 (100) | 36 (100) | 303 (100) |
| Peripheral lymphadenopathy | Peripheral lymphadenopathy | 0 (0) | 0 (0) | 0 (0) |
|  | No peripheral lymphadenopathy | 0 (0) | 0 (0) | 0 (0) |
|  | Unknown | 63 (100) | 36 (100) | 303 (100) |

| **Variable** | **Value** | **Confirmed TB (N=63)** | **Unconfirmed TB (N=36)** | **Unlikely TB (N=303)** |
| --- | --- | --- | --- | --- |
|  |  | n (%) OR median (IQR) | n (%) OR median (IQR) | n (%) OR median (IQR) |
| First Xpert MTB/RIF | Xpert positive for Mtb | 9 (14) | 0 (0) | 0 (0) |
|  | Xpert negative for Mtb | 54 (86) | 36 (100) | 303 (100) |
|  | Unknown/not performed | 0 (0) | 0 (0) | 0 (0) |
| Overall CXR read | CXR consistent with TB | 57 (90) | 31 (86) | 274 (90) |
|  | CXR not consistent with TB | 6 (10) | 5 (14) | 29 (10) |
|  | Unknown/not assessed | 0 (0) | 0 (0) | 0 (0) |
| Opacities on CXR | Opacities present on CXR | 0 (0) | 0 (0) | 0 (0) |
|  | Opacities not present on CXR | 0 (0) | 0 (0) | 0 (0) |
|  | Unknown/not assessed | 63 (100) | 36 (100) | 303 (100) |
| Cavities on CXR | Cavities present on CXR | 0 (0) | 0 (0) | 0 (0) |
|  | Cavities not present on CXR | 0 (0) | 0 (0) | 0 (0) |
|  | Unknown/not assessed | 63 (100) | 36 (100) | 303 (100) |
| Miliary infiltrate on CXR | Miliary infiltrate present on CXR | 0 (0) | 0 (0) | 0 (0) |
|  | Miliary infiltrate not present on CXR | 0 (0) | 0 (0) | 0 (0) |
|  | Unknown/not assessed | 63 (100) | 36 (100) | 303 (100) |
| Intrathoracic lymphadenopathy on CXR | Intrathoracic lymphadenopathy present on CXR | 0 (0) | 0 (0) | 0 (0) |
|  | Intrathoracic lymphadenopathy not present on CXR | 0 (0) | 0 (0) | 0 (0) |
|  | Unknown/not assessed | 63 (100) | 36 (100) | 303 (100) |
| Pleural effusion on CXR | Pleural effusion present on CXR | 0 (0) | 0 (0) | 0 (0) |
|  | Pleural effusion not present on CXR | 0 (0) | 0 (0) | 0 (0) |
|  | Unknown/not assessed | 63 (100) | 36 (100) | 303 (100) |
| Tuberculin skin test | TST positive | 24 (38) | 25 (69) | 36 (12) |
|  | TST negative | 39 (62) | 11 (31) | 265 (87) |
|  | Unknown/not performed | 0 (0) | 0 (0) | 2 (1) |

**Table S29. Description of IPD from Aurilio/2020/BR**

| **Variable** | **Value** | **Confirmed TB (N=9)** | **Unconfirmed TB (N=11)** | **Unlikely TB (N=24)** | **TB-status unknown (N=6)** |
| --- | --- | --- | --- | --- | --- |
|  |  | n (%) OR median (IQR) | n (%) OR median (IQR) | n (%) OR median (IQR) | n (%) OR median (IQR) |
| Age | Months | 31 (9,65) | 58 (17·5,89) | 32·5 (12,72·2) | 33 (18,79·5) |
|  | Unknown | 0 (0) | 0 (0) | 0 (0) | 0 (0) |
| Sex | Male | 2 (22) | 6 (55) | 10 (42) | 1 (17) |
|  | Female | 7 (78) | 5 (45) | 14 (58) | 5 (83) |
|  | Unknown | 0 (0) | 0 (0) | 0 (0) | 0 (0) |
| Weight-for-age | z-score | 0.3 (-1.7,0.6) | -0.3 (-1.7,-0.1) | 0.1 (-1.1,0.7) | -1.2 (-1.3,-0.6) |
|  | Unknown | 0 (0) | 0 (0) | 0 (0) | 1 (17) |
| BCG vaccination | Evidence of BCG vaccination | 9 (100) | 9 (82) | 23 (96) | 4 (67) |
|  | No evidence of BCG vaccination | 0 (0) | 0 (0) | 0 (0) | 0 (0) |
|  | Unknown | 0 (0) | 2 (18) | 1 (4) | 2 (33) |
| HIV status | HIV-positive | 0 (0) | 1 (9) | 4 (17) | 1 (17) |
|  | HIV-negative | 8 (89) | 9 (82) | 6 (25) | 4 (67) |
|  | Unknown | 1 (11) | 1 (9) | 14 (58) | 1 (17) |
| Cough duration | No cough | 4 (44) | 4 (36) | 12 (50) | 2 (33) |
|  | Cough 0-13 days | 1 (11) | 2 (18) | 4 (17) | 2 (33) |
|  | Cough 14-20 days | 2 (22) | 2 (18) | 3 (12) | 1 (17) |
|  | Cough 21-27 days | 0 (0) | 0 (0) | 2 (8) | 0 (0) |
|  | Cough >27 days | 1 (11) | 3 (27) | 3 (12) | 1 (17) |
|  | Unknown | 1 (11) | 0 (0) | 0 (0) | 0 (0) |
| Cough ≥2 weeks | Cough ≥2 weeks present | 3 (33) | 5 (45) | 8 (33) | 2 (33) |
|  | Cough ≥2 weeks not present | 5 (56) | 6 (55) | 16 (67) | 4 (67) |
|  | Unknown | 1 (11) | 0 (0) | 0 (0) | 0 (0) |
| Fever duration | No fever | 3 (33) | 1 (9) | 5 (21) | 1 (17) |
|  | Fever 0-13 days | 2 (22) | 4 (36) | 8 (33) | 2 (33) |
|  | Fever 14-20 days | 1 (11) | 3 (27) | 4 (17) | 2 (33) |
|  | Fever 21-27 days | 0 (0) | 0 (0) | 1 (4) | 0 (0) |
|  | Fever >27 days | 2 (22) | 3 (27) | 6 (25) | 1 (17) |
|  | Unknown | 1 (11) | 0 (0) | 0 (0) | 0 (0) |
| Fever ≥1 week | Fever ≥1 week present | 3 (33) | 8 (73) | 14 (58) | 3 (50) |
|  | Fever ≥1 week not present | 5 (56) | 3 (27) | 10 (42) | 3 (50) |
|  | Unknown | 1 (11) | 0 (0) | 0 (0) | 0 (0) |
| Lethargy | Lethargy | 0 (0) | 1 (9) | 2 (8) | 4 (67) |
|  | No lethargy | 9 (100) | 10 (91) | 22 (92) | 2 (33) |
|  | Unknown | 0 (0) | 0 (0) | 0 (0) | 0 (0) |
| Weight loss | Weight loss | 3 (33) | 4 (36) | 5 (21) | 4 (67) |
|  | No weight loss | 6 (67) | 7 (64) | 19 (79) | 2 (33) |
|  | Unknown | 0 (0) | 0 (0) | 0 (0) | 0 (0) |
| Documented TB exposure | Documented TB exposure in previous 12 months | 6 (67) | 6 (55) | 11 (46) | 2 (33) |
|  | No documented TB exposure in previous 12 months | 3 (33) | 5 (45) | 11 (46) | 3 (50) |
|  | Unknown | 0 (0) | 0 (0) | 2 (8) | 1 (17) |
| Night sweats | Night sweats | 0 (0) | 1 (9) | 0 (0) | 0 (0) |
|  | No night sweats | 9 (100) | 10 (91) | 24 (100) | 6 (100) |
|  | Unknown | 0 (0) | 0 (0) | 0 (0) | 0 (0) |
| Haemoptysis | Haemoptysis | 0 (0) | 0 (0) | 0 (0) | 0 (0) |
|  | No haemoptysis | 9 (100) | 11 (100) | 24 (100) | 6 (100) |
|  | Unknown | 0 (0) | 0 (0) | 0 (0) | 0 (0) |
| Objective fever ( ≥38 degrees Celsius) | Objective fever | 0 (0) | 0 (0) | 0 (0) | 0 (0) |
|  | No objective fever | 0 (0) | 0 (0) | 0 (0) | 0 (0) |
|  | Unknown | 9 (100) | 11 (100) | 24 (100) | 6 (100) |
| Tachycardia | Tachycardia | 0 (0) | 0 (0) | 0 (0) | 0 (0) |
|  | No tachycardia | 0 (0) | 0 (0) | 0 (0) | 0 (0) |
|  | Unknown | 9 (100) | 11 (100) | 24 (100) | 6 (100) |
| Tachypnoea | Tachypnoea | 0 (0) | 0 (0) | 0 (0) | 0 (0) |
|  | No tachypnoea | 0 (0) | 0 (0) | 0 (0) | 0 (0) |
|  | Unknown | 9 (100) | 11 (100) | 24 (100) | 6 (100) |
| Peripheral lymphadenopathy | Peripheral lymphadenopathy | 0 (0) | 0 (0) | 0 (0) | 0 (0) |
|  | No peripheral lymphadenopathy | 9 (100) | 11 (100) | 24 (100) | 6 (100) |
|  | Unknown | 0 (0) | 0 (0) | 0 (0) | 0 (0) |

| **Variable** | **Value** | **Confirmed TB (N=9)** | **Unconfirmed TB (N=11)** | **Unlikely TB (N=24)** | **TB-status unknown (N=6)** |
| --- | --- | --- | --- | --- | --- |
| First Xpert MTB/RIF | Xpert positive for Mtb | 7 (78) | 0 (0) | 0 (0) | 0 (0) |
|  | Xpert negative for Mtb | 2 (22) | 11 (100) | 23 (96) | 6 (100) |
|  | Unknown/not performed | 0 (0) | 0 (0) | 1 (4) | 0 (0) |
| Overall CXR read | CXR consistent with TB | 4 (44) | 8 (73) | 1 (4) | 1 (17) |
|  | CXR not consistent with TB | 5 (56) | 3 (27) | 23 (96) | 5 (83) |
|  | Unknown/not assessed | 0 (0) | 0 (0) | 0 (0) | 0 (0) |
| Opacities on CXR | Opacities present on CXR | 6 (67) | 4 (36) | 15 (62) | 3 (50) |
|  | Opacities not present on CXR | 3 (33) | 7 (64) | 9 (38) | 3 (50) |
|  | Unknown/not assessed | 0 (0) | 0 (0) | 0 (0) | 0 (0) |
| Cavities on CXR | Cavities present on CXR | 0 (0) | 0 (0) | 1 (4) | 1 (17) |
|  | Cavities not present on CXR | 9 (100) | 11 (100) | 23 (96) | 5 (83) |
|  | Unknown/not assessed | 0 (0) | 0 (0) | 0 (0) | 0 (0) |
| Miliary infiltrate on CXR | Miliary infiltrate present on CXR | 1 (11) | 3 (27) | 0 (0) | 0 (0) |
|  | Miliary infiltrate not present on CXR | 8 (89) | 8 (73) | 24 (100) | 6 (100) |
|  | Unknown/not assessed | 0 (0) | 0 (0) | 0 (0) | 0 (0) |
| Intrathoracic lymphadenopathy on CXR | Intrathoracic lymphadenopathy present on CXR | 1 (11) | 2 (18) | 1 (4) | 0 (0) |
|  | Intrathoracic lymphadenopathy not present on CXR | 8 (89) | 9 (82) | 23 (96) | 6 (100) |
|  | Unknown/not assessed | 0 (0) | 0 (0) | 0 (0) | 0 (0) |
| Pleural effusion on CXR | Pleural effusion present on CXR | 3 (33) | 3 (27) | 8 (33) | 2 (33) |
|  | Pleural effusion not present on CXR | 6 (67) | 8 (73) | 16 (67) | 4 (67) |
|  | Unknown/not assessed | 0 (0) | 0 (0) | 0 (0) | 0 (0) |
| Tuberculin skin test | TST positive | 4 (44) | 4 (36) | 4 (17) | 0 (0) |
|  | TST negative | 2 (22) | 6 (55) | 18 (75) | 2 (33) |
|  | Unknown/not performed | 3 (33) | 1 (9) | 2 (8) | 4 (67) |

**Table S30. Description of IPD from Song/2021/KE**

| **Variable** | **Value** | **Confirmed TB (N=31)** | **Unconfirmed TB (N=65)** | **Unlikely TB (N=170)** | **TB-status unknown (N=34)** |
| --- | --- | --- | --- | --- | --- |
|  |  | n (%) OR median (IQR) | n (%) OR median (IQR) | n (%) OR median (IQR) | n (%) OR median (IQR) |
| Age | Months | 23·2 (12·3,38·1) | 24·3 (10·5,39·5) | 28·1 (15·5,46·7) | 17·6 (9·4,34) |
|  | Unknown | 0 (0) | 0 (0) | 0 (0) | 0 (0) |
| Sex | Male | 14 (45) | 31 (48) | 86 (51) | 18 (53) |
|  | Female | 17 (55) | 34 (52) | 84 (49) | 16 (47) |
|  | Unknown | 0 (0) | 0 (0) | 0 (0) | 0 (0) |
| Weight-for-age | z-score | -0.5 (-1.8,0.2) | -1.1 (-2,-0.2) | -0.6 (-1.6,0.4) | -1.7 (-2,0.2) |
|  | Unknown | 6 (19) | 7 (11) | 18 (11) | 24 (71) |
| BCG vaccination | Evidence of BCG vaccination | 27 (87) | 63 (97) | 162 (95) | 33 (97) |
|  | No evidence of BCG vaccination | 4 (13) | 2 (3) | 8 (5) | 1 (3) |
|  | Unknown | 0 (0) | 0 (0) | 0 (0) | 0 (0) |
| HIV status | HIV-positive | 7 (23) | 24 (37) | 33 (19) | 9 (26) |
|  | HIV-negative | 22 (71) | 40 (62) | 136 (80) | 25 (74) |
|  | Unknown | 2 (6) | 1 (2) | 1 (1) | 0 (0) |
| Cough duration | No cough | 5 (16) | 2 (3) | 16 (9) | 2 (6) |
|  | Cough 0-13 days | 1 (3) | 7 (11) | 17 (10) | 4 (12) |
|  | Cough 14-20 days | 2 (6) | 7 (11) | 10 (6) | 3 (9) |
|  | Cough 21-27 days | 3 (10) | 1 (2) | 4 (2) | 0 (0) |
|  | Cough >27 days | 20 (65) | 48 (74) | 120 (71) | 24 (71) |
|  | Unknown | 0 (0) | 0 (0) | 3 (2) | 1 (3) |
| Cough ≥2 weeks | Cough ≥2 weeks present | 25 (81) | 56 (86) | 134 (79) | 27 (79) |
|  | Cough ≥2 weeks not present | 6 (19) | 9 (14) | 33 (19) | 6 (18) |
|  | Unknown | 0 (0) | 0 (0) | 3 (2) | 1 (3) |
| Fever duration | No fever | 11 (35) | 26 (40) | 74 (44) | 14 (41) |
|  | Fever 0-13 days | 7 (23) | 17 (26) | 49 (29) | 7 (21) |
|  | Fever 14-20 days | 7 (23) | 16 (25) | 26 (15) | 5 (15) |
|  | Fever 21-27 days | 4 (13) | 2 (3) | 6 (4) | 2 (6) |
|  | Fever >27 days | 1 (3) | 4 (6) | 12 (7) | 5 (15) |
|  | Unknown | 1 (3) | 0 (0) | 3 (2) | 1 (3) |
| Fever ≥1 week | Fever ≥1 week present | 16 (52) | 31 (48) | 75 (44) | 16 (47) |
|  | Fever ≥1 week not present | 14 (45) | 34 (52) | 92 (54) | 17 (50) |
|  | Unknown | 1 (3) | 0 (0) | 3 (2) | 1 (3) |
| Lethargy | Lethargy | 13 (42) | 20 (31) | 45 (26) | 14 (41) |
|  | No lethargy | 18 (58) | 45 (69) | 125 (74) | 20 (59) |
|  | Unknown | 0 (0) | 0 (0) | 0 (0) | 0 (0) |
| Weight loss | Weight loss | 0 (0) | 3 (5) | 3 (2) | 0 (0) |
|  | No weight loss | 23 (74) | 54 (83) | 144 (85) | 9 (26) |
|  | Unknown | 8 (26) | 8 (12) | 23 (14) | 25 (74) |
| Documented TB exposure | Documented TB exposure in previous 12 months | 24 (77) | 31 (48) | 30 (18) | 4 (12) |
|  | No documented TB exposure in previous 12 months | 7 (23) | 34 (52) | 140 (82) | 30 (88) |
|  | Unknown | 0 (0) | 0 (0) | 0 (0) | 0 (0) |
| Night sweats | Night sweats | 22 (71) | 37 (57) | 83 (49) | 13 (38) |
|  | No night sweats | 9 (29) | 27 (42) | 87 (51) | 21 (62) |
|  | Unknown | 0 (0) | 1 (2) | 0 (0) | 0 (0) |
| Haemoptysis | Haemoptysis | 0 (0) | 0 (0) | 0 (0) | 0 (0) |
|  | No haemoptysis | 0 (0) | 0 (0) | 0 (0) | 0 (0) |
|  | Unknown | 31 (100) | 65 (100) | 170 (100) | 34 (100) |
| Objective fever ( ≥38 degrees Celsius) | Objective fever | 4 (13) | 6 (9) | 7 (4) | 5 (15) |
|  | No objective fever | 27 (87) | 59 (91) | 163 (96) | 29 (85) |
|  | Unknown | 0 (0) | 0 (0) | 0 (0) | 0 (0) |
| Tachycardia | Tachycardia | 0 (0) | 0 (0) | 0 (0) | 0 (0) |
|  | No tachycardia | 0 (0) | 0 (0) | 0 (0) | 0 (0) |
|  | Unknown | 31 (100) | 65 (100) | 170 (100) | 34 (100) |
| Tachypnoea | Tachypnoea | 3 (10) | 3 (5) | 3 (2) | 3 (9) |
|  | No tachypnoea | 28 (90) | 62 (95) | 167 (98) | 31 (91) |
|  | Unknown | 0 (0) | 0 (0) | 0 (0) | 0 (0) |
| Peripheral lymphadenopathy | Peripheral lymphadenopathy | 0 (0) | 0 (0) | 1 (1) | 0 (0) |
|  | No peripheral lymphadenopathy | 25 (81) | 58 (89) | 151 (89) | 10 (29) |
|  | Unknown | 6 (19) | 7 (11) | 18 (11) | 24 (71) |

| **Variable** | **Value** | **Confirmed TB (N=31)** | **Unconfirmed TB (N=65)** | **Unlikely TB (N=170)** | **TB-status unknown (N=34)** |
| --- | --- | --- | --- | --- | --- |
|  |  | n (%) OR median (IQR) | n (%) OR median (IQR) | n (%) OR median (IQR) | n (%) OR median (IQR) |
| First Xpert MTB/RIF | Xpert positive for Mtb | 14 (45) | 0 (0) | 0 (0) | 0 (0) |
|  | Xpert negative for Mtb | 15 (48) | 65 (100) | 168 (99) | 32 (94) |
|  | Unknown/not performed | 2 (6) | 0 (0) | 2 (1) | 2 (6) |
| Overall CXR read | CXR consistent with TB | 13 (42) | 16 (25) | 14 (8) | 2 (6) |
|  | CXR not consistent with TB | 17 (55) | 43 (66) | 150 (88) | 15 (44) |
|  | Unknown/not assessed | 1 (3) | 6 (9) | 6 (4) | 17 (50) |
| Opacities on CXR | Opacities present on CXR | 0 (0) | 0 (0) | 0 (0) | 0 (0) |
|  | Opacities not present on CXR | 0 (0) | 0 (0) | 0 (0) | 0 (0) |
|  | Unknown/not assessed | 31 (100) | 65 (100) | 170 (100) | 34 (100) |
| Cavities on CXR | Cavities present on CXR | 0 (0) | 0 (0) | 0 (0) | 0 (0) |
|  | Cavities not present on CXR | 0 (0) | 0 (0) | 0 (0) | 0 (0) |
|  | Unknown/not assessed | 31 (100) | 65 (100) | 170 (100) | 34 (100) |
| Miliary infiltrate on CXR | Miliary infiltrate present on CXR | 0 (0) | 0 (0) | 0 (0) | 0 (0) |
|  | Miliary infiltrate not present on CXR | 0 (0) | 0 (0) | 0 (0) | 0 (0) |
|  | Unknown/not assessed | 31 (100) | 65 (100) | 170 (100) | 34 (100) |
| Intrathoracic lymphadenopathy on CXR | Intrathoracic lymphadenopathy present on CXR | 0 (0) | 0 (0) | 0 (0) | 0 (0) |
|  | Intrathoracic lymphadenopathy not present on CXR | 0 (0) | 0 (0) | 0 (0) | 0 (0) |
|  | Unknown/not assessed | 31 (100) | 65 (100) | 170 (100) | 34 (100) |
| Pleural effusion on CXR | Pleural effusion present on CXR | 0 (0) | 0 (0) | 0 (0) | 0 (0) |
|  | Pleural effusion not present on CXR | 0 (0) | 0 (0) | 0 (0) | 0 (0) |
|  | Unknown/not assessed | 31 (100) | 65 (100) | 170 (100) | 34 (100) |
| Tuberculin skin test | TST positive | 15 (48) | 15 (23) | 17 (10) | 1 (3) |
|  | TST negative | 11 (35) | 44 (68) | 146 (86) | 23 (68) |
|  | Unknown/not performed | 5 (16) | 6 (9) | 7 (4) | 10 (29) |

**Table S31. Description of IPD from LopezVarela/2015/MZ**

| **Variable** | **Value** | **Confirmed TB (N=13)** | **Unconfirmed TB (N=128)** | **Unlikely TB (N=648)** |
| --- | --- | --- | --- | --- |
|  |  | n (%) OR median (IQR) | n (%) OR median (IQR) | n (%) OR median (IQR) |
| Age | Months | 20·3 (10·6,24·1) | 19·5 (12·9,26) | 19·8 (14,25·7) |
|  | Unknown | 0 (0) | 0 (0) | 0 (0) |
| Sex | Male | 5 (38) | 72 (56) | 353 (54) |
|  | Female | 8 (62) | 56 (44) | 295 (46) |
|  | Unknown | 0 (0) | 0 (0) | 0 (0) |
| Weight-for-age | z-score | -2.3 (-3.8,-1.5) | -2.4 (-3.5,-1.4) | -1.9 (-2.9,-1.1) |
|  | Unknown | 0 (0) | 2 (2) | 10 (2) |
| BCG vaccination | Evidence of BCG vaccination | 13 (100) | 127 (99) | 648 (100) |
|  | No evidence of BCG vaccination | 0 (0) | 1 (1) | 0 (0) |
|  | Unknown | 0 (0) | 0 (0) | 0 (0) |
| HIV status | HIV-positive | 2 (15) | 42 (33) | 60 (9) |
|  | HIV-negative | 11 (85) | 86 (67) | 588 (91) |
|  | Unknown | 0 (0) | 0 (0) | 0 (0) |
| Cough duration | No cough | 8 (62) | 86 (67) | 479 (74) |
|  | Cough 0-13 days | 0 (0) | 24 (19) | 70 (11) |
|  | Cough 14-20 days | 2 (15) | 5 (4) | 34 (5) |
|  | Cough 21-27 days | 1 (8) | 6 (5) | 29 (4) |
|  | Cough >27 days | 2 (15) | 7 (5) | 35 (5) |
|  | Unknown | 0 (0) | 0 (0) | 1 (0) |
| Cough ≥2 weeks | Cough ≥2 weeks present | 5 (38) | 18 (14) | 98 (15) |
|  | Cough ≥2 weeks not present | 8 (62) | 110 (86) | 549 (85) |
|  | Unknown | 0 (0) | 0 (0) | 1 (0) |
| Fever duration | No fever | 9 (69) | 103 (80) | 569 (88) |
|  | Fever 0-13 days | 2 (15) | 22 (17) | 59 (9) |
|  | Fever 14-20 days | 1 (8) | 1 (1) | 10 (2) |
|  | Fever 21-27 days | 0 (0) | 0 (0) | 2 (0) |
|  | Fever >27 days | 0 (0) | 0 (0) | 1 (0) |
|  | Unknown | 1 (8) | 2 (2) | 7 (1) |
| Fever ≥1 week | Fever ≥1 week present | 1 (8) | 4 (3) | 16 (2) |
|  | Fever ≥1 week not present | 11 (85) | 122 (95) | 625 (96) |
|  | Unknown | 1 (8) | 2 (2) | 7 (1) |
| Lethargy | Lethargy | 0 (0) | 0 (0) | 0 (0) |
|  | No lethargy | 0 (0) | 0 (0) | 0 (0) |
|  | Unknown | 13 (100) | 128 (100) | 648 (100) |
| Weight loss | Weight loss | 8 (62) | 106 (83) | 554 (85) |
|  | No weight loss | 5 (38) | 22 (17) | 94 (15) |
|  | Unknown | 0 (0) | 0 (0) | 0 (0) |
| Documented TB exposure | Documented TB exposure in previous 12 months | 2 (15) | 26 (20) | 59 (9) |
|  | No documented TB exposure in previous 12 months | 11 (85) | 102 (80) | 589 (91) |
|  | Unknown | 0 (0) | 0 (0) | 0 (0) |
| Night sweats | Night sweats | 1 (8) | 0 (0) | 0 (0) |
|  | No night sweats | 12 (92) | 128 (100) | 648 (100) |
|  | Unknown | 0 (0) | 0 (0) | 0 (0) |
| Haemoptysis | Haemoptysis | 0 (0) | 0 (0) | 0 (0) |
|  | No haemoptysis | 0 (0) | 0 (0) | 0 (0) |
|  | Unknown | 13 (100) | 128 (100) | 648 (100) |
| Objective fever ( ≥38 degrees Celsius) | Objective fever | 1 (8) | 5 (4) | 21 (3) |
|  | No objective fever | 11 (85) | 119 (93) | 618 (95) |
|  | Unknown | 1 (8) | 4 (3) | 9 (1) |
| Tachycardia | Tachycardia | 1 (8) | 3 (2) | 9 (1) |
|  | No tachycardia | 11 (85) | 123 (96) | 637 (98) |
|  | Unknown | 1 (8) | 2 (2) | 2 (0) |
| Tachypnoea | Tachypnoea | 2 (15) | 10 (8) | 24 (4) |
|  | No tachypnoea | 10 (77) | 116 (91) | 623 (96) |
|  | Unknown | 1 (8) | 2 (2) | 1 (0) |
| Peripheral lymphadenopathy | Peripheral lymphadenopathy | 1 (8) | 2 (2) | 1 (0) |
|  | No peripheral lymphadenopathy | 0 (0) | 0 (0) | 0 (0) |
|  | Unknown | 12 (92) | 126 (98) | 647 (100) |

| **Variable** | **Value** | **Confirmed TB (N=13)** | **Unconfirmed TB (N=128)** | **Unlikely TB (N=648)** |
| --- | --- | --- | --- | --- |
|  |  | n (%) OR median (IQR) | n (%) OR median (IQR) | n (%) OR median (IQR) |
| First Xpert MTB/RIF | Xpert positive for Mtb | 6 (46) | 0 (0) | 0 (0) |
|  | Xpert negative for Mtb | 4 (31) | 99 (77) | 498 (77) |
|  | Unknown/not performed | 3 (23) | 29 (23) | 150 (23) |
| Overall CXR read | CXR consistent with TB | 8 (62) | 62 (48) | 90 (14) |
|  | CXR not consistent with TB | 3 (23) | 66 (52) | 537 (83) |
|  | Unknown/not assessed | 2 (15) | 0 (0) | 21 (3) |
| Opacities on CXR | Opacities present on CXR | 8 (62) | 58 (45) | 89 (14) |
|  | Opacities not present on CXR | 3 (23) | 70 (55) | 538 (83) |
|  | Unknown/not assessed | 2 (15) | 0 (0) | 21 (3) |
| Cavities on CXR | Cavities present on CXR | 0 (0) | 0 (0) | 0 (0) |
|  | Cavities not present on CXR | 0 (0) | 0 (0) | 0 (0) |
|  | Unknown/not assessed | 13 (100) | 128 (100) | 648 (100) |
| Miliary infiltrate on CXR | Miliary infiltrate present on CXR | 0 (0) | 1 (1) | 0 (0) |
|  | Miliary infiltrate not present on CXR | 11 (85) | 127 (99) | 627 (97) |
|  | Unknown/not assessed | 2 (15) | 0 (0) | 21 (3) |
| Intrathoracic lymphadenopathy on CXR | Intrathoracic lymphadenopathy present on CXR | 1 (8) | 8 (6) | 6 (1) |
|  | Intrathoracic lymphadenopathy not present on CXR | 10 (77) | 102 (80) | 589 (91) |
|  | Unknown/not assessed | 2 (15) | 18 (14) | 53 (8) |
| Pleural effusion on CXR | Pleural effusion present on CXR | 0 (0) | 5 (4) | 3 (0) |
|  | Pleural effusion not present on CXR | 11 (85) | 123 (96) | 624 (96) |
|  | Unknown/not assessed | 2 (15) | 0 (0) | 21 (3) |
| Tuberculin skin test | TST positive | 3 (23) | 26 (20) | 45 (7) |
|  | TST negative | 10 (77) | 100 (78) | 603 (93) |
|  | Unknown/not performed | 0 (0) | 2 (2) | 0 (0) |

**Table S32. Description of IPD from Valencia/2017/MZ**

| **Variable** | **Value** | **Confirmed TB (N=5)** | **Unconfirmed TB (N=28)** | **Unlikely TB (N=109)** |
| --- | --- | --- | --- | --- |
|  |  | n (%) OR median (IQR) | n (%) OR median (IQR) | n (%) OR median (IQR) |
| Age | Months | 13·7 (12·1,40·1) | 22·2 (12·3,54·3) | 38·9 (13·8,83·8) |
|  | Unknown | 0 (0) | 0 (0) | 0 (0) |
| Sex | Male | 4 (80) | 16 (57) | 61 (56) |
|  | Female | 1 (20) | 11 (39) | 48 (44) |
|  | Unknown | 0 (0) | 1 (4) | 0 (0) |
| Weight-for-age | z-score | -4 (-4.5,-1.4) | -2.3 (-3.8,-1.2) | -2.1 (-3.5,-0.9) |
|  | Unknown | 0 (0) | 5 (18) | 7 (6) |
| BCG vaccination | Evidence of BCG vaccination | 0 (0) | 0 (0) | 0 (0) |
|  | No evidence of BCG vaccination | 0 (0) | 0 (0) | 0 (0) |
|  | Unknown | 5 (100) | 28 (100) | 109 (100) |
| HIV status | HIV-positive | 3 (60) | 17 (61) | 50 (46) |
|  | HIV-negative | 1 (20) | 9 (32) | 28 (26) |
|  | Unknown | 1 (20) | 2 (7) | 31 (28) |
| Cough duration | No cough | 0 (0) | 3 (11) | 11 (10) |
|  | Cough 0-13 days | 2 (40) | 7 (25) | 40 (37) |
|  | Cough 14-20 days | 1 (20) | 5 (18) | 21 (19) |
|  | Cough 21-27 days | 0 (0) | 1 (4) | 3 (3) |
|  | Cough >27 days | 2 (40) | 10 (36) | 26 (24) |
|  | Unknown | 0 (0) | 2 (7) | 8 (7) |
| Cough ≥2 weeks | Cough ≥2 weeks present | 3 (60) | 16 (57) | 50 (46) |
|  | Cough ≥2 weeks not present | 2 (40) | 10 (36) | 51 (47) |
|  | Unknown | 0 (0) | 2 (7) | 8 (7) |
| Fever duration | No fever | 1 (20) | 6 (21) | 41 (38) |
|  | Fever 0-13 days | 2 (40) | 10 (36) | 46 (42) |
|  | Fever 14-20 days | 0 (0) | 3 (11) | 5 (5) |
|  | Fever 21-27 days | 0 (0) | 1 (4) | 1 (1) |
|  | Fever >27 days | 2 (40) | 6 (21) | 5 (5) |
|  | Unknown | 0 (0) | 2 (7) | 11 (10) |
| Fever ≥1 week | Fever ≥1 week present | 4 (80) | 15 (54) | 32 (29) |
|  | Fever ≥1 week not present | 1 (20) | 11 (39) | 66 (61) |
|  | Unknown | 0 (0) | 2 (7) | 11 (10) |
| Lethargy | Lethargy | 4 (80) | 7 (25) | 45 (41) |
|  | No lethargy | 1 (20) | 21 (75) | 64 (59) |
|  | Unknown | 0 (0) | 0 (0) | 0 (0) |
| Weight loss | Weight loss | 5 (100) | 21 (75) | 66 (61) |
|  | No weight loss | 0 (0) | 7 (25) | 43 (39) |
|  | Unknown | 0 (0) | 0 (0) | 0 (0) |
| Documented TB exposure | Documented TB exposure in previous 12 months | 2 (40) | 9 (32) | 21 (19) |
|  | No documented TB exposure in previous 12 months | 3 (60) | 19 (68) | 88 (81) |
|  | Unknown | 0 (0) | 0 (0) | 0 (0) |
| Night sweats | Night sweats | 1 (20) | 9 (32) | 33 (30) |
|  | No night sweats | 4 (80) | 19 (68) | 76 (70) |
|  | Unknown | 0 (0) | 0 (0) | 0 (0) |
| Haemoptysis | Haemoptysis | 0 (0) | 0 (0) | 0 (0) |
|  | No haemoptysis | 0 (0) | 0 (0) | 0 (0) |
|  | Unknown | 5 (100) | 28 (100) | 109 (100) |
| Objective fever ( ≥38 degrees Celsius) | Objective fever | 0 (0) | 5 (18) | 16 (15) |
|  | No objective fever | 5 (100) | 19 (68) | 82 (75) |
|  | Unknown | 0 (0) | 4 (14) | 11 (10) |
| Tachycardia | Tachycardia | 0 (0) | 0 (0) | 0 (0) |
|  | No tachycardia | 3 (60) | 24 (86) | 0 (0) |
|  | Unknown | 2 (40) | 4 (14) | 109 (100) |
| Tachypnoea | Tachypnoea | 0 (0) | 3 (11) | 0 (0) |
|  | No tachypnoea | 3 (60) | 22 (79) | 0 (0) |
|  | Unknown | 2 (40) | 3 (11) | 109 (100) |
| Peripheral lymphadenopathy | Peripheral lymphadenopathy | 0 (0) | 8 (29) | 20 (18) |
|  | No peripheral lymphadenopathy | 5 (100) | 20 (71) | 89 (82) |
|  | Unknown | 0 (0) | 0 (0) | 0 (0) |

| **Variable** | **Value** | **Confirmed TB (N=5)** | **Unconfirmed TB (N=28)** | **Unlikely TB (N=109)** |
| --- | --- | --- | --- | --- |
|  |  | n (%) OR median (IQR) | n (%) OR median (IQR) | n (%) OR median (IQR) |
| First Xpert MTB/RIF | Xpert positive for Mtb | 5 (100) | 0 (0) | 0 (0) |
|  | Xpert negative for Mtb | 0 (0) | 22 (79) | 91 (83) |
|  | Unknown/not performed | 0 (0) | 6 (21) | 18 (17) |
| Overall CXR read | CXR consistent with TB | 1 (20) | 0 (0) | 0 (0) |
|  | CXR not consistent with TB | 1 (20) | 7 (25) | 28 (26) |
|  | Unknown/not assessed | 3 (60) | 21 (75) | 81 (74) |
| Opacities on CXR | Opacities present on CXR | 2 (40) | 5 (18) | 17 (16) |
|  | Opacities not present on CXR | 0 (0) | 7 (25) | 0 (0) |
|  | Unknown/not assessed | 3 (60) | 16 (57) | 92 (84) |
| Cavities on CXR | Cavities present on CXR | 0 (0) | 3 (11) | 0 (0) |
|  | Cavities not present on CXR | 2 (40) | 9 (32) | 1 (1) |
|  | Unknown/not assessed | 3 (60) | 16 (57) | 108 (99) |
| Miliary infiltrate on CXR | Miliary infiltrate present on CXR | 0 (0) | 0 (0) | 0 (0) |
|  | Miliary infiltrate not present on CXR | 2 (40) | 12 (43) | 1 (1) |
|  | Unknown/not assessed | 3 (60) | 16 (57) | 108 (99) |
| Intrathoracic lymphadenopathy on CXR | Intrathoracic lymphadenopathy present on CXR | 1 (20) | 7 (25) | 0 (0) |
|  | Intrathoracic lymphadenopathy not present on CXR | 1 (20) | 5 (18) | 1 (1) |
|  | Unknown/not assessed | 3 (60) | 16 (57) | 108 (99) |
| Pleural effusion on CXR | Pleural effusion present on CXR | 0 (0) | 2 (7) | 0 (0) |
|  | Pleural effusion not present on CXR | 2 (40) | 10 (36) | 1 (1) |
|  | Unknown/not assessed | 3 (60) | 16 (57) | 108 (99) |
| Tuberculin skin test | TST positive | 0 (0) | 0 (0) | 0 (0) |
|  | TST negative | 1 (20) | 3 (11) | 0 (0) |
|  | Unknown/not performed | 4 (80) | 25 (89) | 109 (100) |

**Table S33. Description of IPD from Myo/2018/MM**

| **Variable** | **Value** | **Confirmed TB (N=27)** | **Unconfirmed TB (N=84)** | **Unlikely TB (N=112)** |
| --- | --- | --- | --- | --- |
|  |  | n (%) OR median (IQR) | n (%) OR median (IQR) | n (%) OR median (IQR) |
| Age | Months | 22 (7,60) | 36 (12,72) | 36 (17·2,72) |
|  | Unknown | 0 (0) | 0 (0) | 0 (0) |
| Sex | Male | 9 (33) | 47 (56) | 65 (58) |
|  | Female | 18 (67) | 37 (44) | 47 (42) |
|  | Unknown | 0 (0) | 0 (0) | 0 (0) |
| Weight-for-age | z-score | -2.2 (-3.4,-1) | -1.7 (-2.9,-0.9) | -1.8 (-3,-1) |
|  | Unknown | 0 (0) | 0 (0) | 0 (0) |
| BCG vaccination | Evidence of BCG vaccination | 13 (48) | 61 (73) | 82 (73) |
|  | No evidence of BCG vaccination | 14 (52) | 23 (27) | 30 (27) |
|  | Unknown | 0 (0) | 0 (0) | 0 (0) |
| HIV status | HIV-positive | 4 (15) | 8 (10) | 15 (13) |
|  | HIV-negative | 23 (85) | 49 (58) | 50 (45) |
|  | Unknown | 0 (0) | 27 (32) | 47 (42) |
| Cough duration | No cough | 0 (0) | 0 (0) | 0 (0) |
|  | Cough 0-13 days | 9 (33) | 45 (54) | 62 (55) |
|  | Cough 14-20 days | 0 (0) | 6 (7) | 10 (9) |
|  | Cough 21-27 days | 18 (67) | 33 (39) | 40 (36) |
|  | Cough >27 days | 0 (0) | 0 (0) | 0 (0) |
|  | Unknown | 0 (0) | 0 (0) | 0 (0) |
| Cough ≥2 weeks | Cough ≥2 weeks present | 18 (67) | 39 (46) | 50 (45) |
|  | Cough ≥2 weeks not present | 9 (33) | 45 (54) | 62 (55) |
|  | Unknown | 0 (0) | 0 (0) | 0 (0) |
| Fever duration | No fever | 0 (0) | 3 (4) | 3 (3) |
|  | Fever 0-13 days | 5 (19) | 35 (42) | 46 (41) |
|  | Fever 14-20 days | 6 (22) | 19 (23) | 34 (30) |
|  | Fever 21-27 days | 1 (4) | 4 (5) | 6 (5) |
|  | Fever >27 days | 15 (56) | 23 (27) | 23 (21) |
|  | Unknown | 0 (0) | 0 (0) | 0 (0) |
| Fever ≥1 week | Fever ≥1 week present | 25 (93) | 73 (87) | 91 (81) |
|  | Fever ≥1 week not present | 2 (7) | 11 (13) | 21 (19) |
|  | Unknown | 0 (0) | 0 (0) | 0 (0) |
| Lethargy | Lethargy | 0 (0) | 0 (0) | 0 (0) |
|  | No lethargy | 0 (0) | 0 (0) | 0 (0) |
|  | Unknown | 27 (100) | 84 (100) | 112 (100) |
| Weight loss | Weight loss | 24 (89) | 59 (70) | 91 (81) |
|  | No weight loss | 3 (11) | 25 (30) | 21 (19) |
|  | Unknown | 0 (0) | 0 (0) | 0 (0) |
| Documented TB exposure | Documented TB exposure in previous 12 months | 11 (41) | 40 (48) | 37 (33) |
|  | No documented TB exposure in previous 12 months | 16 (59) | 44 (52) | 75 (67) |
|  | Unknown | 0 (0) | 0 (0) | 0 (0) |
| Night sweats | Night sweats | 0 (0) | 0 (0) | 0 (0) |
|  | No night sweats | 0 (0) | 0 (0) | 0 (0) |
|  | Unknown | 27 (100) | 84 (100) | 112 (100) |
| Hemoptysis | Haemoptysis | 0 (0) | 0 (0) | 0 (0) |
|  | No haemoptysis | 0 (0) | 0 (0) | 0 (0) |
|  | Unknown | 27 (100) | 84 (100) | 112 (100) |
| Objective fever ( ≥38 degrees Celsius) | Objective fever | 0 (0) | 0 (0) | 0 (0) |
|  | No objective fever | 0 (0) | 0 (0) | 0 (0) |
|  | Unknown | 27 (100) | 84 (100) | 112 (100) |
| Tachycardia | Tachycardia | 0 (0) | 0 (0) | 0 (0) |
|  | No tachycardia | 0 (0) | 0 (0) | 0 (0) |
|  | Unknown | 27 (100) | 84 (100) | 112 (100) |
| Tachypnoea | Tachypnoea | 0 (0) | 0 (0) | 0 (0) |
|  | No tachypnoea | 0 (0) | 0 (0) | 0 (0) |
|  | Unknown | 27 (100) | 84 (100) | 112 (100) |
| Peripheral lymphadenopathy | Peripheral lymphadenopathy | 0 (0) | 0 (0) | 0 (0) |
|  | No peripheral lymphadenopathy | 0 (0) | 0 (0) | 0 (0) |
|  | Unknown | 27 (100) | 84 (100) | 112 (100) |

| **Variable** | **Value** | **Confirmed TB (N=27)** | **Unconfirmed TB (N=84)** | **Unlikely TB (N=112)** |
| --- | --- | --- | --- | --- |
|  |  | n (%) OR median (IQR) | n (%) OR median (IQR) | n (%) OR median (IQR) |
| First Xpert MTB/RIF | Xpert positive for Mtb | 26 (96) | 0 (0) | 0 (0) |
|  | Xpert negative for Mtb | 1 (4) | 84 (100) | 108 (96) |
|  | Unknown/not performed | 0 (0) | 0 (0) | 4 (4) |
| Overall CXR read | CXR consistent with TB | 26 (96) | 61 (73) | 44 (39) |
|  | CXR not consistent with TB | 0 (0) | 20 (24) | 43 (38) |
|  | Unknown/not assessed | 1 (4) | 3 (4) | 25 (22) |
| Opacities on CXR | Opacities present on CXR | 0 (0) | 0 (0) | 0 (0) |
|  | Opacities not present on CXR | 0 (0) | 0 (0) | 0 (0) |
|  | Unknown/not assessed | 27 (100) | 84 (100) | 112 (100) |
| Cavities on CXR | Cavities present on CXR | 0 (0) | 0 (0) | 0 (0) |
|  | Cavities not present on CXR | 0 (0) | 0 (0) | 0 (0) |
|  | Unknown/not assessed | 27 (100) | 84 (100) | 112 (100) |
| Miliary infiltrate on CXR | Miliary infiltrate present on CXR | 0 (0) | 0 (0) | 0 (0) |
|  | Miliary infiltrate not present on CXR | 0 (0) | 0 (0) | 0 (0) |
|  | Unknown/not assessed | 27 (100) | 84 (100) | 112 (100) |
| Intrathoracic lymphadenopathy on CXR | Intrathoracic lymphadenopathy present on CXR | 0 (0) | 0 (0) | 0 (0) |
|  | Intrathoracic lymphadenopathy not present on CXR | 0 (0) | 0 (0) | 0 (0) |
|  | Unknown/not assessed | 27 (100) | 84 (100) | 112 (100) |
| Pleural effusion on CXR | Pleural effusion present on CXR | 0 (0) | 0 (0) | 0 (0) |
|  | Pleural effusion not present on CXR | 0 (0) | 0 (0) | 0 (0) |
|  | Unknown/not assessed | 27 (100) | 84 (100) | 112 (100) |
| Tuberculin skin test | TST positive | 15 (56) | 18 (21) | 4 (4) |
|  | TST negative | 12 (44) | 66 (79) | 108 (96) |
|  | Unknown/not performed | 0 (0) | 0 (0) | 0 (0) |

**Table S34. Description of IPD from Marcy/2019/Multi**

| **Variable** | **Value** | **Confirmed TB (N=41)** | **Unconfirmed TB (N=155)** | **Unlikely TB (N=142)** |
| --- | --- | --- | --- | --- |
|  |  | n (%) OR median (IQR) | n (%) OR median (IQR) | n (%) OR median (IQR) |
| Age | Months | 85·6 (37·7,97·7) | 70·8 (35·5,94·3) | 60·5 (19·4,98·6) |
|  | Unknown | 0 (0) | 0 (0) | 0 (0) |
| Sex | Male | 22 (54) | 85 (55) | 61 (43) |
|  | Female | 19 (46) | 70 (45) | 81 (57) |
|  | Unknown | 0 (0) | 0 (0) | 0 (0) |
| Weight-for-age | z-score | -2.5 (-3.2,-1.3) | -2.8 (-3.9,-2) | -2.5 (-3.6,-1.3) |
|  | Unknown | 1 (2) | 0 (0) | 0 (0) |
| BCG vaccination | Evidence of BCG vaccination | 37 (90) | 114 (74) | 119 (84) |
|  | No evidence of BCG vaccination | 3 (7) | 41 (26) | 22 (15) |
|  | Unknown | 1 (2) | 0 (0) | 1 (1) |
| HIV status | HIV-positive | 41 (100) | 155 (100) | 142 (100) |
|  | HIV-negative | 0 (0) | 0 (0) | 0 (0) |
|  | Unknown | 0 (0) | 0 (0) | 0 (0) |
| Cough duration | No cough | 5 (12) | 7 (5) | 9 (6) |
|  | Cough 0-13 days | 3 (7) | 12 (8) | 20 (14) |
|  | Cough 14-20 days | 7 (17) | 38 (25) | 34 (24) |
|  | Cough 21-27 days | 2 (5) | 17 (11) | 14 (10) |
|  | Cough >27 days | 23 (56) | 81 (52) | 65 (46) |
|  | Unknown | 1 (2) | 0 (0) | 0 (0) |
| Cough ≥2 weeks | Cough ≥2 weeks present | 32 (78) | 136 (88) | 113 (80) |
|  | Cough ≥2 weeks not present | 8 (20) | 19 (12) | 29 (20) |
|  | Unknown | 1 (2) | 0 (0) | 0 (0) |
| Fever duration | No fever | 4 (10) | 23 (15) | 46 (32) |
|  | Fever 0-13 days | 8 (20) | 25 (16) | 47 (33) |
|  | Fever 14-20 days | 11 (27) | 38 (25) | 19 (13) |
|  | Fever 21-27 days | 1 (2) | 16 (10) | 8 (6) |
|  | Fever >27 days | 14 (34) | 50 (32) | 20 (14) |
|  | Unknown | 3 (7) | 3 (2) | 2 (1) |
| Fever ≥1 week | Fever ≥1 week present | 0 (0) | 0 (0) | 0 (0) |
|  | Fever ≥1 week not present | 0 (0) | 0 (0) | 0 (0) |
|  | Unknown | 41 (100) | 155 (100) | 142 (100) |
| Lethargy | Lethargy | 23 (56) | 77 (50) | 63 (44) |
|  | No lethargy | 16 (39) | 76 (49) | 79 (56) |
|  | Unknown | 2 (5) | 2 (1) | 0 (0) |
| Weight loss | Weight loss | 35 (85) | 104 (67) | 83 (58) |
|  | No weight loss | 5 (12) | 51 (33) | 59 (42) |
|  | Unknown | 1 (2) | 0 (0) | 0 (0) |
| Documented TB exposure | Documented TB exposure in previous 12 months | 8 (20) | 14 (9) | 9 (6) |
|  | No documented TB exposure in previous 12 months | 33 (80) | 141 (91) | 133 (94) |
|  | Unknown | 0 (0) | 0 (0) | 0 (0) |
| Night sweats | Night sweats | 10 (24) | 71 (46) | 42 (30) |
|  | No night sweats | 30 (73) | 83 (54) | 100 (70) |
|  | Unknown | 1 (2) | 1 (1) | 0 (0) |
| Hemoptysis | Haemoptysis | 1 (2) | 10 (6) | 1 (1) |
|  | No haemoptysis | 39 (95) | 145 (94) | 140 (99) |
|  | Unknown | 1 (2) | 0 (0) | 1 (1) |
| Objective fever ( ≥38 degrees Celsius) | Objective fever | 20 (49) | 22 (14) | 25 (18) |
|  | No objective fever | 20 (49) | 133 (86) | 113 (80) |
|  | Unknown | 1 (2) | 0 (0) | 4 (3) |
| Tachycardia | Tachycardia | 15 (37) | 21 (14) | 12 (8) |
|  | No tachycardia | 25 (61) | 133 (86) | 124 (87) |
|  | Unknown | 1 (2) | 1 (1) | 6 (4) |
| Tachypnoea | Tachypnoea | 14 (34) | 47 (30) | 40 (28) |
|  | No tachypnoea | 22 (54) | 106 (68) | 90 (63) |
|  | Unknown | 5 (12) | 2 (1) | 12 (8) |
| Peripheral lymphadenopathy | Peripheral lymphadenopathy | 14 (34) | 53 (34) | 48 (34) |
|  | No peripheral lymphadenopathy | 26 (63) | 99 (64) | 90 (63) |
|  | Unknown | 1 (2) | 3 (2) | 4 (3) |

| **Variable** | **Value** | **Confirmed TB (N=41)** | **Unconfirmed TB (N=155)** | **Unlikely TB (N=142)** |
| --- | --- | --- | --- | --- |
|  |  | n (%) OR median (IQR) | n (%) OR median (IQR) | n (%) OR median (IQR) |
| First Xpert MTB/RIF | Xpert positive for Mtb | 21 (51) | 0 (0) | 0 (0) |
|  | Xpert negative for Mtb | 19 (46) | 154 (99) | 132 (93) |
|  | Unknown/not performed | 1 (2) | 1 (1) | 10 (7) |
| Overall CXR read | CXR consistent with TB | 27 (66) | 112 (72) | 65 (46) |
|  | CXR not consistent with TB | 12 (29) | 39 (25) | 66 (46) |
|  | Unknown/not assessed | 2 (5) | 4 (3) | 11 (8) |
| Opacities on CXR | Opacities present on CXR | 15 (37) | 62 (40) | 35 (25) |
|  | Opacities not present on CXR | 24 (59) | 89 (57) | 95 (67) |
|  | Unknown/not assessed | 2 (5) | 4 (3) | 12 (8) |
| Cavities on CXR | Cavities present on CXR | 1 (2) | 6 (4) | 0 (0) |
|  | Cavities not present on CXR | 38 (93) | 145 (94) | 129 (91) |
|  | Unknown/not assessed | 2 (5) | 4 (3) | 13 (9) |
| Miliary infiltrate on CXR | Miliary infiltrate present on CXR | 3 (7) | 8 (5) | 5 (4) |
|  | Miliary infiltrate not present on CXR | 36 (88) | 143 (92) | 125 (88) |
|  | Unknown/not assessed | 2 (5) | 4 (3) | 12 (8) |
| Intrathoracic lymphadenopathy on CXR | Intrathoracic lymphadenopathy present on CXR | 5 (12) | 67 (43) | 17 (12) |
|  | Intrathoracic lymphadenopathy not present on CXR | 34 (83) | 84 (54) | 113 (80) |
|  | Unknown/not assessed | 2 (5) | 4 (3) | 12 (8) |
| Pleural effusion on CXR | Pleural effusion present on CXR | 0 (0) | 7 (5) | 5 (4) |
|  | Pleural effusion not present on CXR | 39 (95) | 144 (93) | 125 (88) |
|  | Unknown/not assessed | 2 (5) | 4 (3) | 12 (8) |
| Tuberculin skin test | TST positive | 7 (17) | 10 (6) | 8 (6) |
|  | TST negative | 29 (71) | 133 (86) | 114 (80) |
|  | Unknown/not performed | 5 (12) | 12 (8) | 20 (14) |

**Table S35. Description of IPD from Hamid/2019/PK**

| **Variable** | **Value** | **Confirmed TB (N=0)** | **Unconfirmed TB (N=29)** | **Unlikely TB (N=416)** |
| --- | --- | --- | --- | --- |
|  |  | n (%) OR median (IQR) | n (%) OR median (IQR) | n (%) OR median (IQR) |
| Age | Months | NA (NA,NA) | 66 (30,90) | 66 (42,90) |
|  | Unknown | 0 (NaN) | 0 (0) | 0 (0) |
| Sex | Male | 0 (NaN) | 12 (41) | 222 (53) |
|  | Female | 0 (NaN) | 17 (59) | 194 (47) |
|  | Unknown | 0 (NaN) | 0 (0) | 0 (0) |
| Weight-for-age | z-score | NA (NA,NA) | -2.4 (-3.5,-1.6) | -2.2 (-3.2,-1.5) |
|  | Unknown | 0 (NaN) | 0 (0) | 83 (20) |
| BCG vaccination | Evidence of BCG vaccination | 0 (NaN) | 0 (0) | 0 (0) |
|  | No evidence of BCG vaccination | 0 (NaN) | 0 (0) | 0 (0) |
|  | Unknown | 0 (NaN) | 29 (100) | 416 (100) |
| HIV status | HIV-positive | 0 (NaN) | 0 (0) | 0 (0) |
|  | HIV-negative | 0 (NaN) | 29 (100) | 416 (100) |
|  | Unknown | 0 (NaN) | 0 (0) | 0 (0) |
| Cough duration | No cough | 0 (NaN) | 5 (17) | 175 (42) |
|  | Cough 0-13 days | 0 (NaN) | 1 (3) | 60 (14) |
|  | Cough 14-20 days | 0 (NaN) | 17 (59) | 46 (11) |
|  | Cough 21-27 days | 0 (NaN) | 6 (21) | 67 (16) |
|  | Cough >27 days | 0 (NaN) | 0 (0) | 64 (15) |
|  | Unknown | 0 (NaN) | 0 (0) | 4 (1) |
| Cough ≥2 weeks | Cough ≥2 weeks present | 0 (NaN) | 23 (79) | 177 (43) |
|  | Cough ≥2 weeks not present | 0 (NaN) | 6 (21) | 235 (56) |
|  | Unknown | 0 (NaN) | 0 (0) | 4 (1) |
| Fever duration | No fever | 0 (NaN) | 7 (24) | 152 (37) |
|  | Fever 0-13 days | 0 (NaN) | 11 (38) | 157 (38) |
|  | Fever 14-20 days | 0 (NaN) | 10 (34) | 66 (16) |
|  | Fever 21-27 days | 0 (NaN) | 1 (3) | 27 (6) |
|  | Fever >27 days | 0 (NaN) | 0 (0) | 6 (1) |
|  | Unknown | 0 (NaN) | 0 (0) | 8 (2) |
| Fever ≥1 week | Fever ≥1 week present | 0 (NaN) | 11 (38) | 99 (24) |
|  | Fever ≥1 week not present | 0 (NaN) | 18 (62) | 309 (74) |
|  | Unknown | 0 (NaN) | 0 (0) | 8 (2) |
| Lethargy | Lethargy | 0 (NaN) | 2 (7) | 6 (1) |
|  | No lethargy | 0 (NaN) | 27 (93) | 410 (99) |
|  | Unknown | 0 (NaN) | 0 (0) | 0 (0) |
| Weight loss | Weight loss | 0 (NaN) | 14 (48) | 58 (14) |
|  | No weight loss | 0 (NaN) | 15 (52) | 358 (86) |
|  | Unknown | 0 (NaN) | 0 (0) | 0 (0) |
| Documented TB exposure | Documented TB exposure in previous 12 months | 0 (NaN) | 13 (45) | 16 (4) |
|  | No documented TB exposure in previous 12 months | 0 (NaN) | 16 (55) | 400 (96) |
|  | Unknown | 0 (NaN) | 0 (0) | 0 (0) |
| Night sweats | Night sweats | 0 (NaN) | 2 (7) | 11 (3) |
|  | No night sweats | 0 (NaN) | 27 (93) | 405 (97) |
|  | Unknown | 0 (NaN) | 0 (0) | 0 (0) |
| Haemoptysis | Haemoptysis | 0 (NaN) | 0 (0) | 1 (0) |
|  | No haemoptysis | 0 (NaN) | 29 (100) | 415 (100) |
|  | Unknown | 0 (NaN) | 0 (0) | 0 (0) |
| Objective fever ( ≥38 degrees Celsius) | Objective fever | 0 (NaN) | 0 (0) | 0 (0) |
|  | No objective fever | 0 (NaN) | 0 (0) | 0 (0) |
|  | Unknown | 0 (NaN) | 29 (100) | 416 (100) |
| Tachycardia | Tachycardia | 0 (NaN) | 0 (0) | 0 (0) |
|  | No tachycardia | 0 (NaN) | 0 (0) | 0 (0) |
|  | Unknown | 0 (NaN) | 29 (100) | 416 (100) |
| Tachypnoea | Tachypnoea | 0 (NaN) | 0 (0) | 0 (0) |
|  | No tachypnoea | 0 (NaN) | 0 (0) | 0 (0) |
|  | Unknown | 0 (NaN) | 29 (100) | 416 (100) |
| Peripheral lymphadenopathy | Peripheral lymphadenopathy | 0 (NaN) | 2 (7) | 34 (8) |
|  | No peripheral lymphadenopathy | 0 (NaN) | 27 (93) | 382 (92) |
|  | Unknown | 0 (NaN) | 0 (0) | 0 (0) |

| **Variable** | **Value** | **Confirmed TB (N=0)** | **Unconfirmed TB (N=29)** | **Unlikely TB (N=416)** |
| --- | --- | --- | --- | --- |
|  |  | n (%) OR median (IQR) | n (%) OR median (IQR) | n (%) OR median (IQR) |
| First Xpert MTB/RIF | Xpert positive for Mtb | 0 (NaN) | 0 (0) | 0 (0) |
|  | Xpert negative for Mtb | 0 (NaN) | 7 (24) | 156 (38) |
|  | Unknown/not performed | 0 (NaN) | 22 (76) | 260 (62) |
| Overall CXR read | CXR consistent with TB | 0 (NaN) | 20 (69) | 29 (7) |
|  | CXR not consistent with TB | 0 (NaN) | 4 (14) | 278 (67) |
|  | Unknown/not assessed | 0 (NaN) | 5 (17) | 109 (26) |
| Opacities on CXR | Opacities present on CXR | 0 (NaN) | 0 (0) | 0 (0) |
|  | Opacities not present on CXR | 0 (NaN) | 0 (0) | 0 (0) |
|  | Unknown/not assessed | 0 (NaN) | 29 (100) | 416 (100) |
| Cavities on CXR | Cavities present on CXR | 0 (NaN) | 0 (0) | 0 (0) |
|  | Cavities not present on CXR | 0 (NaN) | 0 (0) | 0 (0) |
|  | Unknown/not assessed | 0 (NaN) | 29 (100) | 416 (100) |
| Miliary infiltrate on CXR | Miliary infiltrate present on CXR | 0 (NaN) | 0 (0) | 0 (0) |
|  | Miliary infiltrate not present on CXR | 0 (NaN) | 0 (0) | 0 (0) |
|  | Unknown/not assessed | 0 (NaN) | 29 (100) | 416 (100) |
| Intrathoracic lymphadenopathy on CXR | Intrathoracic lymphadenopathy present on CXR | 0 (NaN) | 0 (0) | 0 (0) |
|  | Intrathoracic lymphadenopathy not present on CXR | 0 (NaN) | 0 (0) | 0 (0) |
|  | Unknown/not assessed | 0 (NaN) | 29 (100) | 416 (100) |
| Pleural effusion on CXR | Pleural effusion present on CXR | 0 (NaN) | 0 (0) | 0 (0) |
|  | Pleural effusion not present on CXR | 0 (NaN) | 0 (0) | 0 (0) |
|  | Unknown/not assessed | 0 (NaN) | 29 (100) | 416 (100) |
| Tuberculin skin test | TST positive | 0 (NaN) | 10 (34) | 82 (20) |
|  | TST negative | 0 (NaN) | 0 (0) | 4 (1) |
|  | Unknown/not performed | 0 (NaN) | 19 (66) | 330 (79) |

**Table S36. Description of IPD from Zar/2019/ZA**

| **Variable** | **Value** | **Confirmed TB (N=189)** | **Unconfirmed TB (N=274)** | **Unlikely TB (N=303)** |
| --- | --- | --- | --- | --- |
|  |  | n (%) OR median (IQR) | n (%) OR median (IQR) | n (%) OR median (IQR) |
| Age | Months | 32·4 (17,61·6) | 25·1 (11·9,53·9) | 24 (12·4,50·5) |
|  | Unknown | 0 (0) | 0 (0) | 0 (0) |
| Sex | Male | 111 (59) | 155 (57) | 156 (51) |
|  | Female | 78 (41) | 119 (43) | 147 (49) |
|  | Unknown | 0 (0) | 0 (0) | 0 (0) |
| Weight-for-age | z-score | -1.3 (-2.2,-0.2) | -0.9 (-1.7,0) | -0.8 (-1.9,0.1) |
|  | Unknown | 0 (0) | 2 (1) | 1 (0) |
| BCG vaccination | Evidence of BCG vaccination | 163 (86) | 252 (92) | 281 (93) |
|  | No evidence of BCG vaccination | 0 (0) | 2 (1) | 1 (0) |
|  | Unknown | 26 (14) | 20 (7) | 21 (7) |
| HIV status | HIV-positive | 29 (15) | 57 (21) | 51 (17) |
|  | HIV-negative | 160 (85) | 217 (79) | 252 (83) |
|  | Unknown | 0 (0) | 0 (0) | 0 (0) |
| Cough duration | No cough | 33 (17) | 62 (23) | 47 (16) |
|  | Cough 0-13 days | 71 (38) | 109 (40) | 134 (44) |
|  | Cough 14-20 days | 37 (20) | 33 (12) | 47 (16) |
|  | Cough 21-27 days | 16 (8) | 12 (4) | 16 (5) |
|  | Cough >27 days | 27 (14) | 51 (19) | 48 (16) |
|  | Unknown | 5 (3) | 7 (3) | 11 (4) |
| Cough ≥2 weeks | Cough ≥2 weeks present | 80 (42) | 96 (35) | 111 (37) |
|  | Cough ≥2 weeks not present | 104 (55) | 171 (62) | 181 (60) |
|  | Unknown | 5 (3) | 7 (3) | 11 (4) |
| Fever duration | No fever | 72 (38) | 103 (38) | 106 (35) |
|  | Fever 0-13 days | 70 (37) | 118 (43) | 145 (48) |
|  | Fever 14-20 days | 22 (12) | 21 (8) | 17 (6) |
|  | Fever 21-27 days | 5 (3) | 12 (4) | 9 (3) |
|  | Fever >27 days | 15 (8) | 13 (5) | 13 (4) |
|  | Unknown | 5 (3) | 7 (3) | 13 (4) |
| Fever ≥1 week | Fever ≥1 week present | 66 (35) | 85 (31) | 87 (29) |
|  | Fever ≥1 week not present | 118 (62) | 182 (66) | 203 (67) |
|  | Unknown | 5 (3) | 7 (3) | 13 (4) |
| Lethargy | Lethargy | 91 (48) | 124 (45) | 124 (41) |
|  | No lethargy | 93 (49) | 138 (50) | 163 (54) |
|  | Unknown | 5 (3) | 12 (4) | 16 (5) |
| Weight loss | Weight loss | 146 (77) | 182 (66) | 206 (68) |
|  | No weight loss | 41 (22) | 85 (31) | 88 (29) |
|  | Unknown | 2 (1) | 7 (3) | 9 (3) |
| Documented TB exposure | Documented TB exposure in previous 12 months | 113 (60) | 160 (58) | 184 (61) |
|  | No documented TB exposure in previous 12 months | 76 (40) | 114 (42) | 119 (39) |
|  | Unknown | 0 (0) | 0 (0) | 0 (0) |
| Night sweats | Night sweats | 109 (58) | 149 (54) | 131 (43) |
|  | No night sweats | 79 (42) | 121 (44) | 164 (54) |
|  | Unknown | 1 (1) | 4 (1) | 8 (3) |
| Haemoptysis | Haemoptysis | 0 (0) | 0 (0) | 0 (0) |
|  | No haemoptysis | 0 (0) | 0 (0) | 0 (0) |
|  | Unknown | 189 (100) | 274 (100) | 303 (100) |
| Objective fever ( ≥38 degrees Celsius) | Objective fever | 42 (22) | 48 (18) | 52 (17) |
|  | No objective fever | 145 (77) | 220 (80) | 243 (80) |
|  | Unknown | 2 (1) | 6 (2) | 8 (3) |
| Tachycardia | Tachycardia | 51 (27) | 85 (31) | 87 (29) |
|  | No tachycardia | 129 (68) | 171 (62) | 203 (67) |
|  | Unknown | 9 (5) | 18 (7) | 13 (4) |
| Tachypnoea | Tachypnoea | 61 (32) | 92 (34) | 116 (38) |
|  | No tachypnoea | 120 (63) | 166 (61) | 178 (59) |
|  | Unknown | 8 (4) | 16 (6) | 9 (3) |
| Peripheral lymphadenopathy | Peripheral lymphadenopathy | 80 (42) | 107 (39) | 91 (30) |
|  | No peripheral lymphadenopathy | 106 (56) | 157 (57) | 205 (68) |
|  | Unknown | 3 (2) | 10 (4) | 7 (2) |

| **Variable** | **Value** | **Confirmed TB (N=189)** | **Unconfirmed TB (N=274)** | **Unlikely TB (N=303)** |
| --- | --- | --- | --- | --- |
|  |  | n (%) OR median (IQR) | n (%) OR median (IQR) | n (%) OR median (IQR) |
| First Xpert MTB/RIF | Xpert positive for Mtb | 98 (52) | 0 (0) | 0 (0) |
|  | Xpert negative for Mtb | 91 (48) | 271 (99) | 303 (100) |
|  | Unknown/not performed | 0 (0) | 3 (1) | 0 (0) |
| Overall CXR read | CXR consistent with TB | 95 (50) | 104 (38) | 66 (22) |
|  | CXR not consistent with TB | 17 (9) | 76 (28) | 94 (31) |
|  | Unknown/not assessed | 77 (41) | 94 (34) | 143 (47) |
| Opacities on CXR | Opacities present on CXR | 130 (69) | 165 (60) | 194 (64) |
|  | Opacities not present on CXR | 59 (31) | 109 (40) | 108 (36) |
|  | Unknown/not assessed | 0 (0) | 0 (0) | 1 (0) |
| Cavities on CXR | Cavities present on CXR | 21 (11) | 6 (2) | 12 (4) |
|  | Cavities not present on CXR | 168 (89) | 268 (98) | 291 (96) |
|  | Unknown/not assessed | 0 (0) | 0 (0) | 0 (0) |
| Miliary infiltrate on CXR | Miliary infiltrate present on CXR | 21 (11) | 2 (1) | 2 (1) |
|  | Miliary infiltrate not present on CXR | 168 (89) | 272 (99) | 301 (99) |
|  | Unknown/not assessed | 0 (0) | 0 (0) | 0 (0) |
| Intrathoracic lymphadenopathy on CXR | Intrathoracic lymphadenopathy present on CXR | 75 (40) | 94 (34) | 56 (18) |
|  | Intrathoracic lymphadenopathy not present on CXR | 114 (60) | 180 (66) | 246 (81) |
|  | Unknown/not assessed | 0 (0) | 0 (0) | 1 (0) |
| Pleural effusion on CXR | Pleural effusion present on CXR | 44 (23) | 39 (14) | 23 (8) |
|  | Pleural effusion not present on CXR | 145 (77) | 235 (86) | 278 (92) |
|  | Unknown/not assessed | 0 (0) | 0 (0) | 2 (1) |
| Tuberculin skin test | TST positive | 138 (73) | 173 (63) | 42 (14) |
|  | TST negative | 32 (17) | 81 (30) | 230 (76) |
|  | Unknown/not performed | 19 (10) | 20 (7) | 31 (10) |

**Table S37. Description of IPD from Walters/2017/ZA**

| **Variable** | **Value** | **Confirmed TB (N=119)** | **Unconfirmed TB (N=180)** | **Unlikely TB (N=283)** | **TB-status unknown (N=13)** |
| --- | --- | --- | --- | --- | --- |
|  |  | n (%) OR median (IQR) | n (%) OR median (IQR) | n (%) OR median (IQR) | n (%) OR median (IQR) |
| Age | Months | 18·8 (9·7,44·7) | 16·4 (10·5,29·3) | 15·1 (9·1,28·4) | 11·5 (7·1,22·6) |
|  | Unknown | 0 (0) | 0 (0) | 0 (0) | 0 (0) |
| Sex | Male | 53 (45) | 106 (59) | 153 (54) | 8 (62) |
|  | Female | 66 (55) | 74 (41) | 130 (46) | 5 (38) |
|  | Unknown | 0 (0) | 0 (0) | 0 (0) | 0 (0) |
| Weight-for-age | z-score | -1.5 (-2.6,-0.7) | -1.2 (-2.4,-0.2) | -1.2 (-2,-0.3) | -1.6 (-2.3,-0.6) |
|  | Unknown | 0 (0) | 0 (0) | 0 (0) | 0 (0) |
| BCG vaccination | Evidence of BCG vaccination | 108 (91) | 179 (99) | 278 (98) | 13 (100) |
|  | No evidence of BCG vaccination | 11 (9) | 1 (1) | 5 (2) | 0 (0) |
|  | Unknown | 0 (0) | 0 (0) | 0 (0) | 0 (0) |
| HIV status | HIV-positive | 16 (13) | 32 (18) | 21 (7) | 1 (8) |
|  | HIV-negative | 103 (87) | 148 (82) | 262 (93) | 12 (92) |
|  | Unknown | 0 (0) | 0 (0) | 0 (0) | 0 (0) |
| Cough duration | No cough | 18 (15) | 35 (19) | 64 (23) | 13 (100) |
|  | Cough 0-13 days | 43 (36) | 65 (36) | 120 (42) | 0 (0) |
|  | Cough 14-20 days | 24 (20) | 22 (12) | 35 (12) | 0 (0) |
|  | Cough 21-27 days | 8 (7) | 13 (7) | 18 (6) | 0 (0) |
|  | Cough >27 days | 26 (22) | 44 (24) | 41 (14) | 0 (0) |
|  | Unknown | 0 (0) | 1 (1) | 5 (2) | 0 (0) |
| Cough ≥2 weeks | Cough ≥2 weeks present | 58 (49) | 79 (44) | 94 (33) | 0 (0) |
|  | Cough ≥2 weeks not present | 61 (51) | 100 (56) | 184 (65) | 13 (100) |
|  | Unknown | 0 (0) | 1 (1) | 5 (2) | 0 (0) |
| Fever duration | No fever | 45 (38) | 78 (43) | 148 (52) | 4 (31) |
|  | Fever 0-13 days | 26 (22) | 32 (18) | 61 (22) | 2 (15) |
|  | Fever 14-20 days | 3 (3) | 1 (1) | 4 (1) | 2 (15) |
|  | Fever 21-27 days | 2 (2) | 1 (1) | 2 (1) | 0 (0) |
|  | Fever >27 days | 3 (3) | 6 (3) | 5 (2) | 0 (0) |
|  | Unknown | 40 (34) | 62 (34) | 63 (22) | 5 (38) |
| Fever ≥1 week | Fever ≥1 week present | 15 (13) | 16 (9) | 19 (7) | 3 (23) |
|  | Fever ≥1 week not present | 64 (54) | 102 (57) | 201 (71) | 5 (38) |
|  | Unknown | 40 (34) | 62 (34) | 63 (22) | 5 (38) |
| Lethargy | Lethargy | 51 (43) | 80 (44) | 97 (34) | 6 (46) |
|  | No lethargy | 68 (57) | 100 (56) | 186 (66) | 7 (54) |
|  | Unknown | 0 (0) | 0 (0) | 0 (0) | 0 (0) |
| Weight loss | Weight loss | 55 (46) | 83 (46) | 109 (39) | 7 (54) |
|  | No weight loss | 64 (54) | 97 (54) | 174 (61) | 6 (46) |
|  | Unknown | 0 (0) | 0 (0) | 0 (0) | 0 (0) |
| Documented TB exposure | Documented TB exposure in previous 12 months | 61 (51) | 93 (52) | 61 (22) | 5 (38) |
|  | No documented TB exposure in previous 12 months | 58 (49) | 87 (48) | 222 (78) | 8 (62) |
|  | Unknown | 0 (0) | 0 (0) | 0 (0) | 0 (0) |
| Night sweats | Night sweats | 0 (0) | 0 (0) | 0 (0) | 0 (0) |
|  | No night sweats | 0 (0) | 0 (0) | 0 (0) | 0 (0) |
|  | Unknown | 119 (100) | 180 (100) | 283 (100) | 13 (100) |
| Haemoptysis | Haemoptysis | 0 (0) | 0 (0) | 0 (0) | 0 (0) |
|  | No haemoptysis | 0 (0) | 0 (0) | 0 (0) | 0 (0) |
|  | Unknown | 119 (100) | 180 (100) | 283 (100) | 13 (100) |
| Objective fever ( ≥38 degrees Celsius) | Objective fever | 0 (0) | 0 (0) | 0 (0) | 0 (0) |
|  | No objective fever | 0 (0) | 0 (0) | 0 (0) | 0 (0) |
|  | Unknown | 119 (100) | 180 (100) | 283 (100) | 13 (100) |
| Tachycardia | Tachycardia | 0 (0) | 0 (0) | 0 (0) | 0 (0) |
|  | No tachycardia | 0 (0) | 0 (0) | 0 (0) | 0 (0) |
|  | Unknown | 119 (100) | 180 (100) | 283 (100) | 13 (100) |
| Tachypnoea | Tachypnoea | 39 (33) | 61 (34) | 79 (28) | 4 (31) |
|  | No tachypnoea | 78 (66) | 116 (64) | 202 (71) | 9 (69) |
|  | Unknown | 2 (2) | 3 (2) | 2 (1) | 0 (0) |
| Peripheral lymphadenopathy | Peripheral lymphadenopathy | 34 (29) | 51 (28) | 56 (20) | 1 (8) |
|  | No peripheral lymphadenopathy | 85 (71) | 129 (72) | 227 (80) | 12 (92) |
|  | Unknown | 0 (0) | 0 (0) | 0 (0) | 0 (0) |

| **Variable** | **Value** | **Confirmed TB (N=119)** | **Unconfirmed TB (N=180)** | **Unlikely TB (N=283)** | **TB-status unknown (N=13)** |
| --- | --- | --- | --- | --- | --- |
|  |  | n (%) OR median (IQR) | n (%) OR median (IQR) | n (%) OR median (IQR) | n (%) OR median (IQR) |
| First Xpert MTB/RIF | Xpert positive for Mtb | 37 (31) | 0 (0) | 0 (0) | 0 (0) |
|  | Xpert negative for Mtb | 69 (58) | 141 (78) | 233 (82) | 11 (85) |
|  | Unknown/not performed | 13 (11) | 39 (22) | 50 (18) | 2 (15) |
| Overall CXR read | CXR consistent with TB | 83 (70) | 61 (34) | 25 (9) | 2 (15) |
|  | CXR not consistent with TB | 29 (24) | 105 (58) | 235 (83) | 11 (85) |
|  | Unknown/not assessed | 7 (6) | 14 (8) | 23 (8) | 0 (0) |
| Opacities on CXR | Opacities present on CXR | 60 (50) | 74 (41) | 126 (45) | 6 (46) |
|  | Opacities not present on CXR | 59 (50) | 106 (59) | 157 (55) | 7 (54) |
|  | Unknown/not assessed | 0 (0) | 0 (0) | 0 (0) | 0 (0) |
| Cavities on CXR | Cavities present on CXR | 9 (8) | 5 (3) | 6 (2) | 0 (0) |
|  | Cavities not present on CXR | 110 (92) | 175 (97) | 277 (98) | 13 (100) |
|  | Unknown/not assessed | 0 (0) | 0 (0) | 0 (0) | 0 (0) |
| Miliary infiltrate on CXR | Miliary infiltrate present on CXR | 6 (5) | 3 (2) | 0 (0) | 0 (0) |
|  | Miliary infiltrate not present on CXR | 113 (95) | 177 (98) | 283 (100) | 13 (100) |
|  | Unknown/not assessed | 0 (0) | 0 (0) | 0 (0) | 0 (0) |
| Intrathoracic lymphadenopathy on CXR | Intrathoracic lymphadenopathy present on CXR | 69 (58) | 50 (28) | 14 (5) | 1 (8) |
|  | Intrathoracic lymphadenopathy not present on CXR | 50 (42) | 130 (72) | 269 (95) | 12 (92) |
|  | Unknown/not assessed | 0 (0) | 0 (0) | 0 (0) | 0 (0) |
| Pleural effusion on CXR | Pleural effusion present on CXR | 8 (7) | 4 (2) | 7 (2) | 0 (0) |
|  | Pleural effusion not present on CXR | 111 (93) | 176 (98) | 276 (98) | 13 (100) |
|  | Unknown/not assessed | 0 (0) | 0 (0) | 0 (0) | 0 (0) |
| Tuberculin skin test | TST positive | 63 (53) | 48 (27) | 14 (5) | 3 (23) |
|  | TST negative | 26 (22) | 95 (53) | 200 (71) | 8 (62) |
|  | Unknown/not performed | 30 (25) | 37 (21) | 69 (24) | 2 (15) |

**Table S38. Description of IPD from Orikiriza/2018/UG**

| **Variable** | **Value** | **Confirmed TB (N=12)** | **Unconfirmed TB (N=145)** | **Unlikely TB (N=167)** | **TB-status unknown (N=14)** |
| --- | --- | --- | --- | --- | --- |
|  |  | n (%) OR median (IQR) | n (%) OR median (IQR) | n (%) OR median (IQR) | n (%) OR median (IQR) |
| Age | Months | 31 (20·8,66) | 28 (12,60) | 47 (16,72) | 20 (10·5,66) |
|  | Unknown | 0 (0) | 0 (0) | 0 (0) | 0 (0) |
| Sex | Male | 9 (75) | 82 (57) | 91 (54) | 5 (36) |
|  | Female | 3 (25) | 63 (43) | 76 (46) | 9 (64) |
|  | Unknown | 0 (0) | 0 (0) | 0 (0) | 0 (0) |
| Weight-for-age | z-score | -2 (-3,-0.9) | -1.5 (-2.9,-0.4) | -1.2 (-2.6,-0.2) | -1.9 (-2.9,-1.1) |
|  | Unknown | 0 (0) | 0 (0) | 0 (0) | 0 (0) |
| BCG vaccination | Evidence of BCG vaccination | 10 (83) | 123 (85) | 151 (90) | 11 (79) |
|  | No evidence of BCG vaccination | 1 (8) | 4 (3) | 3 (2) | 1 (7) |
|  | Unknown | 1 (8) | 18 (12) | 13 (8) | 2 (14) |
| HIV status | HIV-positive | 5 (42) | 43 (30) | 51 (31) | 2 (14) |
|  | HIV-negative | 6 (50) | 100 (69) | 115 (69) | 12 (86) |
|  | Unknown | 1 (8) | 2 (1) | 1 (1) | 0 (0) |
| Cough duration | No cough | 0 (0) | 0 (0) | 0 (0) | 0 (0) |
|  | Cough 0-13 days | 0 (0) | 0 (0) | 0 (0) | 0 (0) |
|  | Cough 14-20 days | 0 (0) | 0 (0) | 0 (0) | 0 (0) |
|  | Cough 21-27 days | 0 (0) | 0 (0) | 0 (0) | 0 (0) |
|  | Cough >27 days | 0 (0) | 0 (0) | 0 (0) | 0 (0) |
|  | Unknown | 12 (100) | 145 (100) | 167 (100) | 14 (100) |
| Cough ≥2 weeks | Cough ≥2 weeks present | 12 (100) | 129 (89) | 161 (96) | 10 (71) |
|  | Cough ≥2 weeks not present | 0 (0) | 16 (11) | 6 (4) | 4 (29) |
|  | Unknown | 0 (0) | 0 (0) | 0 (0) | 0 (0) |
| Fever duration | No fever | 0 (0) | 0 (0) | 0 (0) | 0 (0) |
|  | Fever 0-13 days | 0 (0) | 0 (0) | 0 (0) | 0 (0) |
|  | Fever 14-20 days | 0 (0) | 0 (0) | 0 (0) | 0 (0) |
|  | Fever 21-27 days | 0 (0) | 0 (0) | 0 (0) | 0 (0) |
|  | Fever >27 days | 0 (0) | 0 (0) | 0 (0) | 0 (0) |
|  | Unknown | 12 (100) | 145 (100) | 167 (100) | 14 (100) |
| Fever ≥1 week | Fever ≥1 week present | 7 (58) | 59 (41) | 54 (32) | 6 (43) |
|  | Fever ≥1 week not present | 5 (42) | 86 (59) | 113 (68) | 8 (57) |
|  | Unknown | 0 (0) | 0 (0) | 0 (0) | 0 (0) |
| Lethargy | Lethargy | 7 (58) | 53 (37) | 49 (29) | 6 (43) |
|  | No lethargy | 5 (42) | 92 (63) | 118 (71) | 8 (57) |
|  | Unknown | 0 (0) | 0 (0) | 0 (0) | 0 (0) |
| Weight loss | Weight loss | 8 (67) | 73 (50) | 90 (54) | 9 (64) |
|  | No weight loss | 4 (33) | 72 (50) | 77 (46) | 5 (36) |
|  | Unknown | 0 (0) | 0 (0) | 0 (0) | 0 (0) |
| Documented TB exposure | Documented TB exposure in previous 12 months | 4 (33) | 58 (40) | 3 (2) | 5 (36) |
|  | No documented TB exposure in previous 12 months | 8 (67) | 86 (59) | 164 (98) | 9 (64) |
|  | Unknown | 0 (0) | 1 (1) | 0 (0) | 0 (0) |
| Night sweats | Night sweats | 3 (25) | 44 (30) | 42 (25) | 2 (14) |
|  | No night sweats | 9 (75) | 100 (69) | 123 (74) | 11 (79) |
|  | Unknown | 0 (0) | 1 (1) | 2 (1) | 1 (7) |
| Haemoptysis | Haemoptysis | 0 (0) | 2 (1) | 3 (2) | 0 (0) |
|  | No haemoptysis | 12 (100) | 142 (98) | 162 (97) | 13 (93) |
|  | Unknown | 0 (0) | 1 (1) | 2 (1) | 1 (7) |
| Objective fever ( ≥38 degrees Celsius) | Objective fever | 2 (17) | 5 (3) | 8 (5) | 0 (0) |
|  | No objective fever | 10 (83) | 140 (97) | 159 (95) | 14 (100) |
|  | Unknown | 0 (0) | 0 (0) | 0 (0) | 0 (0) |
| Tachycardia | Tachycardia | 4 (33) | 16 (11) | 23 (14) | 3 (21) |
|  | No tachycardia | 8 (67) | 127 (88) | 143 (86) | 11 (79) |
|  | Unknown | 0 (0) | 2 (1) | 1 (1) | 0 (0) |
| Tachypnoea | Tachypnoea | 3 (25) | 33 (23) | 38 (23) | 2 (14) |
|  | No tachypnoea | 9 (75) | 111 (77) | 129 (77) | 12 (86) |
|  | Unknown | 0 (0) | 1 (1) | 0 (0) | 0 (0) |
| Peripheral lymphadenopathy | Peripheral lymphadenopathy | 0 (0) | 3 (2) | 4 (2) | 0 (0) |
|  | No peripheral lymphadenopathy | 12 (100) | 139 (96) | 160 (96) | 12 (86) |
|  | Unknown | 0 (0) | 3 (2) | 3 (2) | 2 (14) |

| **Variable** | **Value** | **Confirmed TB (N=12)** | **Unconfirmed TB (N=145)** | **Unlikely TB (N=167)** | **TB-status unknown (N=14)** |
| --- | --- | --- | --- | --- | --- |
|  |  | n (%) OR median (IQR) | n (%) OR median (IQR) | n (%) OR median (IQR) | n (%) OR median (IQR) |
| First Xpert MTB/RIF | Xpert positive for Mtb | 9 (75) | 0 (0) | 0 (0) | 0 (0) |
|  | Xpert negative for Mtb | 1 (8) | 124 (86) | 151 (90) | 10 (71) |
|  | Unknown/not performed | 2 (17) | 21 (14) | 16 (10) | 4 (29) |
| Overall CXR read | CXR consistent with TB | 7 (58) | 84 (58) | 26 (16) | 7 (50) |
|  | CXR not consistent with TB | 5 (42) | 57 (39) | 140 (84) | 4 (29) |
|  | Unknown/not assessed | 0 (0) | 4 (3) | 1 (1) | 3 (21) |
| Opacities on CXR | Opacities present on CXR | 10 (83) | 81 (56) | 61 (37) | 7 (50) |
|  | Opacities not present on CXR | 2 (17) | 60 (41) | 105 (63) | 4 (29) |
|  | Unknown/not assessed | 0 (0) | 4 (3) | 1 (1) | 3 (21) |
| Cavities on CXR | Cavities present on CXR | 1 (8) | 3 (2) | 0 (0) | 0 (0) |
|  | Cavities not present on CXR | 11 (92) | 138 (95) | 166 (99) | 11 (79) |
|  | Unknown/not assessed | 0 (0) | 4 (3) | 1 (1) | 3 (21) |
| Miliary infiltrate on CXR | Miliary infiltrate present on CXR | 0 (0) | 2 (1) | 0 (0) | 0 (0) |
|  | Miliary infiltrate not present on CXR | 12 (100) | 139 (96) | 166 (99) | 11 (79) |
|  | Unknown/not assessed | 0 (0) | 4 (3) | 1 (1) | 3 (21) |
| Intrathoracic lymphadenopathy on CXR | Intrathoracic lymphadenopathy present on CXR | 1 (8) | 14 (10) | 7 (4) | 1 (7) |
|  | Intrathoracic lymphadenopathy not present on CXR | 11 (92) | 131 (90) | 160 (96) | 13 (93) |
|  | Unknown/not assessed | 0 (0) | 0 (0) | 0 (0) | 0 (0) |
| Pleural effusion on CXR | Pleural effusion present on CXR | 1 (8) | 1 (1) | 3 (2) | 0 (0) |
|  | Pleural effusion not present on CXR | 11 (92) | 140 (97) | 163 (98) | 11 (79) |
|  | Unknown/not assessed | 0 (0) | 4 (3) | 1 (1) | 3 (21) |
| Tuberculin skin test | TST positive | 7 (58) | 72 (50) | 1 (1) | 4 (29) |
|  | TST negative | 5 (42) | 71 (49) | 162 (97) | 8 (57) |
|  | Unknown/not performed | 0 (0) | 2 (1) | 4 (2) | 2 (14) |

**Table S39. Description of IPD from Orikiriza/2022/UG**

| **Variable** | **Value** | **Confirmed TB (N=12)** | **Unconfirmed TB (N=58)** | **Unlikely TB (N=125)** | **TB-status unknown (N=22)** |
| --- | --- | --- | --- | --- | --- |
|  |  | n (%) OR median (IQR) | n (%) OR median (IQR) | n (%) OR median (IQR) | n (%) OR median (IQR) |
| Age | Months | 10·6 (5·1,31·2) | 17·8 (12·3,37·6) | 15 (9,22·9) | 23·3 (10·9,39·8) |
|  | Unknown | 0 (0) | 0 (0) | 0 (0) | 0 (0) |
| Sex | Male | 6 (50) | 24 (41) | 68 (54) | 12 (55) |
|  | Female | 6 (50) | 34 (59) | 57 (46) | 10 (45) |
|  | Unknown | 0 (0) | 0 (0) | 0 (0) | 0 (0) |
| Weight-for-age | z-score | -4.9 (-5.8,-2.9) | -3.6 (-5,-2.6) | -4.2 (-5.2,-2.8) | -4.2 (-5.2,-2.3) |
|  | Unknown | 0 (0) | 0 (0) | 0 (0) | 1 (5) |
| BCG vaccination | Evidence of BCG vaccination | 12 (100) | 53 (91) | 118 (94) | 19 (86) |
|  | No evidence of BCG vaccination | 0 (0) | 5 (9) | 7 (6) | 3 (14) |
|  | Unknown | 0 (0) | 0 (0) | 0 (0) | 0 (0) |
| HIV status | HIV-positive | 4 (33) | 17 (29) | 37 (30) | 12 (55) |
|  | HIV-negative | 8 (67) | 41 (71) | 82 (66) | 10 (45) |
|  | Unknown | 0 (0) | 0 (0) | 6 (5) | 0 (0) |
| Cough duration | No cough | 0 (0) | 0 (0) | 0 (0) | 0 (0) |
|  | Cough 0-13 days | 0 (0) | 0 (0) | 0 (0) | 0 (0) |
|  | Cough 14-20 days | 0 (0) | 0 (0) | 0 (0) | 0 (0) |
|  | Cough 21-27 days | 0 (0) | 0 (0) | 0 (0) | 0 (0) |
|  | Cough >27 days | 0 (0) | 0 (0) | 0 (0) | 0 (0) |
|  | Unknown | 12 (100) | 58 (100) | 125 (100) | 22 (100) |
| Cough ≥2 weeks | Cough ≥2 weeks present | 2 (17) | 7 (12) | 15 (12) | 2 (9) |
|  | Cough ≥2 weeks not present | 9 (75) | 50 (86) | 102 (82) | 20 (91) |
|  | Unknown | 1 (8) | 1 (2) | 8 (6) | 0 (0) |
| Fever duration | No fever | 0 (0) | 0 (0) | 0 (0) | 0 (0) |
|  | Fever 0-13 days | 0 (0) | 0 (0) | 0 (0) | 0 (0) |
|  | Fever 14-20 days | 0 (0) | 0 (0) | 0 (0) | 0 (0) |
|  | Fever 21-27 days | 0 (0) | 0 (0) | 0 (0) | 0 (0) |
|  | Fever >27 days | 0 (0) | 0 (0) | 0 (0) | 0 (0) |
|  | Unknown | 12 (100) | 58 (100) | 125 (100) | 22 (100) |
| Fever ≥1 week | Fever ≥1 week present | 3 (25) | 11 (19) | 28 (22) | 6 (27) |
|  | Fever ≥1 week not present | 8 (67) | 43 (74) | 81 (65) | 14 (64) |
|  | Unknown | 1 (8) | 4 (7) | 16 (13) | 2 (9) |
| Lethargy | Lethargy | 12 (100) | 55 (95) | 120 (96) | 21 (95) |
|  | No lethargy | 0 (0) | 3 (5) | 5 (4) | 1 (5) |
|  | Unknown | 0 (0) | 0 (0) | 0 (0) | 0 (0) |
| Weight loss | Weight loss | 11 (92) | 48 (83) | 106 (85) | 16 (73) |
|  | No weight loss | 1 (8) | 10 (17) | 19 (15) | 6 (27) |
|  | Unknown | 0 (0) | 0 (0) | 0 (0) | 0 (0) |
| Documented TB exposure | Documented TB exposure in previous 12 months | 3 (25) | 15 (26) | 0 (0) | 0 (0) |
|  | No documented TB exposure in previous 12 months | 9 (75) | 40 (69) | 121 (97) | 22 (100) |
|  | Unknown | 0 (0) | 3 (5) | 4 (3) | 0 (0) |
| Night sweats | Night sweats | 4 (33) | 24 (41) | 34 (27) | 5 (23) |
|  | No night sweats | 8 (67) | 34 (59) | 91 (73) | 17 (77) |
|  | Unknown | 0 (0) | 0 (0) | 0 (0) | 0 (0) |
| Haemoptysis | Haemoptysis | 0 (0) | 0 (0) | 0 (0) | 0 (0) |
|  | No haemoptysis | 0 (0) | 0 (0) | 0 (0) | 0 (0) |
|  | Unknown | 12 (100) | 58 (100) | 125 (100) | 22 (100) |
| Objective fever ( ≥38 degrees Celsius) | Objective fever | 2 (17) | 10 (17) | 27 (22) | 7 (32) |
|  | No objective fever | 10 (83) | 48 (83) | 98 (78) | 14 (64) |
|  | Unknown | 0 (0) | 0 (0) | 0 (0) | 1 (5) |
| Tachycardia | Tachycardia | 9 (75) | 25 (43) | 44 (35) | 15 (68) |
|  | No tachycardia | 3 (25) | 33 (57) | 80 (64) | 6 (27) |
|  | Unknown | 0 (0) | 0 (0) | 1 (1) | 1 (5) |
| Tachypnoea | Tachypnoea | 6 (50) | 15 (26) | 20 (16) | 8 (36) |
|  | No tachypnoea | 6 (50) | 43 (74) | 105 (84) | 13 (59) |
|  | Unknown | 0 (0) | 0 (0) | 0 (0) | 1 (5) |
| Peripheral lymphadenopathy | Peripheral lymphadenopathy | 5 (42) | 15 (26) | 25 (20) | 6 (27) |
|  | No peripheral lymphadenopathy | 7 (58) | 43 (74) | 100 (80) | 16 (73) |
|  | Unknown | 0 (0) | 0 (0) | 0 (0) | 0 (0) |

| **Variable** | **Value** | **Confirmed TB (N=12)** | **Unconfirmed TB (N=58)** | **Unlikely TB (N=125)** | **TB-status unknown (N=22)** |
| --- | --- | --- | --- | --- | --- |
|  |  | n (%) OR median (IQR) | n (%) OR median (IQR) | n (%) OR median (IQR) | n (%) OR median (IQR) |
| First Xpert MTB/RIF | Xpert positive for Mtb | 8 (67) | 0 (0) | 0 (0) | 0 (0) |
|  | Xpert negative for Mtb | 4 (33) | 58 (100) | 124 (99) | 17 (77) |
|  | Unknown/not performed | 0 (0) | 0 (0) | 1 (1) | 5 (23) |
| Overall CXR read | CXR consistent with TB | 4 (33) | 10 (17) | 4 (3) | 1 (5) |
|  | CXR not consistent with TB | 6 (50) | 48 (83) | 121 (97) | 6 (27) |
|  | Unknown/not assessed | 2 (17) | 0 (0) | 0 (0) | 15 (68) |
| Opacities on CXR | Opacities present on CXR | 6 (50) | 38 (66) | 57 (46) | 4 (18) |
|  | Opacities not present on CXR | 4 (33) | 11 (19) | 30 (24) | 1 (5) |
|  | Unknown/not assessed | 2 (17) | 9 (16) | 38 (30) | 17 (77) |
| Cavities on CXR | Cavities present on CXR | 0 (0) | 2 (3) | 1 (1) | 0 (0) |
|  | Cavities not present on CXR | 10 (83) | 47 (81) | 86 (69) | 5 (23) |
|  | Unknown/not assessed | 2 (17) | 9 (16) | 38 (30) | 17 (77) |
| Miliary infiltrate on CXR | Miliary infiltrate present on CXR | 2 (17) | 0 (0) | 0 (0) | 1 (5) |
|  | Miliary infiltrate not present on CXR | 8 (67) | 49 (84) | 87 (70) | 4 (18) |
|  | Unknown/not assessed | 2 (17) | 9 (16) | 38 (30) | 17 (77) |
| Intrathoracic lymphadenopathy on CXR | Intrathoracic lymphadenopathy present on CXR | 1 (8) | 6 (10) | 3 (2) | 0 (0) |
|  | Intrathoracic lymphadenopathy not present on CXR | 9 (75) | 43 (74) | 84 (67) | 5 (23) |
|  | Unknown/not assessed | 2 (17) | 9 (16) | 38 (30) | 17 (77) |
| Pleural effusion on CXR | Pleural effusion present on CXR | 0 (0) | 2 (3) | 0 (0) | 0 (0) |
|  | Pleural effusion not present on CXR | 10 (83) | 47 (81) | 87 (70) | 5 (23) |
|  | Unknown/not assessed | 2 (17) | 9 (16) | 38 (30) | 17 (77) |
| Tuberculin skin test | TST positive | 4 (33) | 3 (5) | 0 (0) | 0 (0) |
|  | TST negative | 6 (50) | 52 (90) | 117 (94) | 11 (50) |
|  | Unknown/not performed | 2 (17) | 3 (5) | 8 (6) | 11 (50) |

**Table S40. Description of IPD from Giang/2015/VN**

| **Variable** | **Value** | **Confirmed TB (N=20)** | **Unconfirmed TB (N=77)** | **Unlikely TB (N=16)** |
| --- | --- | --- | --- | --- |
|  |  | n (%) OR median (IQR) | n (%) OR median (IQR) | n (%) OR median (IQR) |
| Age | Months | 13 (7·8,23·8) | 14 (9,20) | 18 (9·8,44·5) |
|  | Unknown | 0 (0) | 0 (0) | 0 (0) |
| Sex | Male | 14 (70) | 55 (71) | 9 (56) |
|  | Female | 6 (30) | 22 (29) | 7 (44) |
|  | Unknown | 0 (0) | 0 (0) | 0 (0) |
| Weight-for-age | z-score | -0.9 (-1.7,-0.1) | -0.7 (-1.7,0.2) | -1.3 (-2,-0.9) |
|  | Unknown | 0 (0) | 0 (0) | 0 (0) |
| BCG vaccination | Evidence of BCG vaccination | 16 (80) | 71 (92) | 16 (100) |
|  | No evidence of BCG vaccination | 3 (15) | 6 (8) | 0 (0) |
|  | Unknown | 1 (5) | 0 (0) | 0 (0) |
| HIV status | HIV-positive | 0 (0) | 0 (0) | 0 (0) |
|  | HIV-negative | 20 (100) | 77 (100) | 16 (100) |
|  | Unknown | 0 (0) | 0 (0) | 0 (0) |
| Cough duration | No cough | 0 (0) | 0 (0) | 0 (0) |
|  | Cough 0-13 days | 0 (0) | 0 (0) | 0 (0) |
|  | Cough 14-20 days | 0 (0) | 0 (0) | 0 (0) |
|  | Cough 21-27 days | 0 (0) | 0 (0) | 0 (0) |
|  | Cough >27 days | 0 (0) | 0 (0) | 0 (0) |
|  | Unknown | 20 (100) | 77 (100) | 16 (100) |
| Cough ≥2 weeks | Cough ≥2 weeks present | 14 (70) | 65 (84) | 14 (88) |
|  | Cough ≥2 weeks not present | 6 (30) | 12 (16) | 2 (12) |
|  | Unknown | 0 (0) | 0 (0) | 0 (0) |
| Fever duration | No fever | 0 (0) | 0 (0) | 0 (0) |
|  | Fever 0-13 days | 0 (0) | 0 (0) | 0 (0) |
|  | Fever 14-20 days | 0 (0) | 0 (0) | 0 (0) |
|  | Fever 21-27 days | 0 (0) | 0 (0) | 0 (0) |
|  | Fever >27 days | 0 (0) | 0 (0) | 0 (0) |
|  | Unknown | 20 (100) | 77 (100) | 16 (100) |
| Fever ≥1 week | Fever ≥1 week present | 11 (55) | 43 (56) | 9 (56) |
|  | Fever ≥1 week not present | 9 (45) | 34 (44) | 7 (44) |
|  | Unknown | 0 (0) | 0 (0) | 0 (0) |
| Lethargy | Lethargy | 7 (35) | 27 (35) | 5 (31) |
|  | No lethargy | 13 (65) | 50 (65) | 11 (69) |
|  | Unknown | 0 (0) | 0 (0) | 0 (0) |
| Weight loss | Weight loss | 9 (45) | 33 (43) | 5 (31) |
|  | No weight loss | 11 (55) | 44 (57) | 11 (69) |
|  | Unknown | 0 (0) | 0 (0) | 0 (0) |
| Documented TB exposure | Documented TB exposure in previous 12 months | 6 (30) | 9 (12) | 1 (6) |
|  | No documented TB exposure in previous 12 months | 14 (70) | 68 (88) | 15 (94) |
|  | Unknown | 0 (0) | 0 (0) | 0 (0) |
| Night sweats | Night sweats | 14 (70) | 60 (78) | 13 (81) |
|  | No night sweats | 6 (30) | 17 (22) | 3 (19) |
|  | Unknown | 0 (0) | 0 (0) | 0 (0) |
| Haemoptysis | Haemoptysis | 1 (5) | 0 (0) | 0 (0) |
|  | No haemoptysis | 19 (95) | 77 (100) | 16 (100) |
|  | Unknown | 0 (0) | 0 (0) | 0 (0) |
| Objective fever ( ≥38 degrees Celsius) | Objective fever | 0 (0) | 0 (0) | 0 (0) |
|  | No objective fever | 0 (0) | 0 (0) | 0 (0) |
|  | Unknown | 20 (100) | 77 (100) | 16 (100) |
| Tachycardia | Tachycardia | 0 (0) | 0 (0) | 0 (0) |
|  | No tachycardia | 0 (0) | 0 (0) | 0 (0) |
|  | Unknown | 20 (100) | 77 (100) | 16 (100) |
| Tachypnoea | Tachypnoea | 0 (0) | 0 (0) | 0 (0) |
|  | No tachypnoea | 0 (0) | 0 (0) | 0 (0) |
|  | Unknown | 20 (100) | 77 (100) | 16 (100) |
| Peripheral lymphadenopathy | Peripheral lymphadenopathy | 0 (0) | 1 (1) | 0 (0) |
|  | No peripheral lymphadenopathy | 20 (100) | 76 (99) | 16 (100) |
|  | Unknown | 0 (0) | 0 (0) | 0 (0) |

| **Variable** | **Value** | **Confirmed TB (N=20)** | **Unconfirmed TB (N=77)** | **Unlikely TB (N=16)** |
| --- | --- | --- | --- | --- |
|  |  | n (%) OR median (IQR) | n (%) OR median (IQR) | n (%) OR median (IQR) |
| First Xpert MTB/RIF | Xpert positive for Mtb | 7 (35) | 0 (0) | 0 (0) |
|  | Xpert negative for Mtb | 13 (65) | 76 (99) | 16 (100) |
|  | Unknown/not performed | 0 (0) | 1 (1) | 0 (0) |
| Overall CXR read | CXR consistent with TB | 19 (95) | 76 (99) | 15 (94) |
|  | CXR not consistent with TB | 0 (0) | 1 (1) | 0 (0) |
|  | Unknown/not assessed | 1 (5) | 0 (0) | 1 (6) |
| Opacities on CXR | Opacities present on CXR | 0 (0) | 0 (0) | 0 (0) |
|  | Opacities not present on CXR | 0 (0) | 0 (0) | 0 (0) |
|  | Unknown/not assessed | 20 (100) | 77 (100) | 16 (100) |
| Cavities on CXR | Cavities present on CXR | 0 (0) | 0 (0) | 0 (0) |
|  | Cavities not present on CXR | 0 (0) | 0 (0) | 0 (0) |
|  | Unknown/not assessed | 20 (100) | 77 (100) | 16 (100) |
| Miliary infiltrate on CXR | Miliary infiltrate present on CXR | 0 (0) | 0 (0) | 0 (0) |
|  | Miliary infiltrate not present on CXR | 0 (0) | 0 (0) | 0 (0) |
|  | Unknown/not assessed | 20 (100) | 77 (100) | 16 (100) |
| Intrathoracic lymphadenopathy on CXR | Intrathoracic lymphadenopathy present on CXR | 0 (0) | 0 (0) | 0 (0) |
|  | Intrathoracic lymphadenopathy not present on CXR | 0 (0) | 0 (0) | 0 (0) |
|  | Unknown/not assessed | 20 (100) | 77 (100) | 16 (100) |
| Pleural effusion on CXR | Pleural effusion present on CXR | 0 (0) | 0 (0) | 0 (0) |
|  | Pleural effusion not present on CXR | 0 (0) | 0 (0) | 0 (0) |
|  | Unknown/not assessed | 20 (100) | 77 (100) | 16 (100) |
| Tuberculin skin test | TST positive | 18 (90) | 60 (78) | 9 (56) |
|  | TST negative | 1 (5) | 12 (16) | 5 (31) |
|  | Unknown/not performed | 1 (5) | 5 (6) | 2 (12) |

### Appendix J: Missingness in IPD received

**Figure S2. Missingness in IPD received** (note variables names per Table S1)

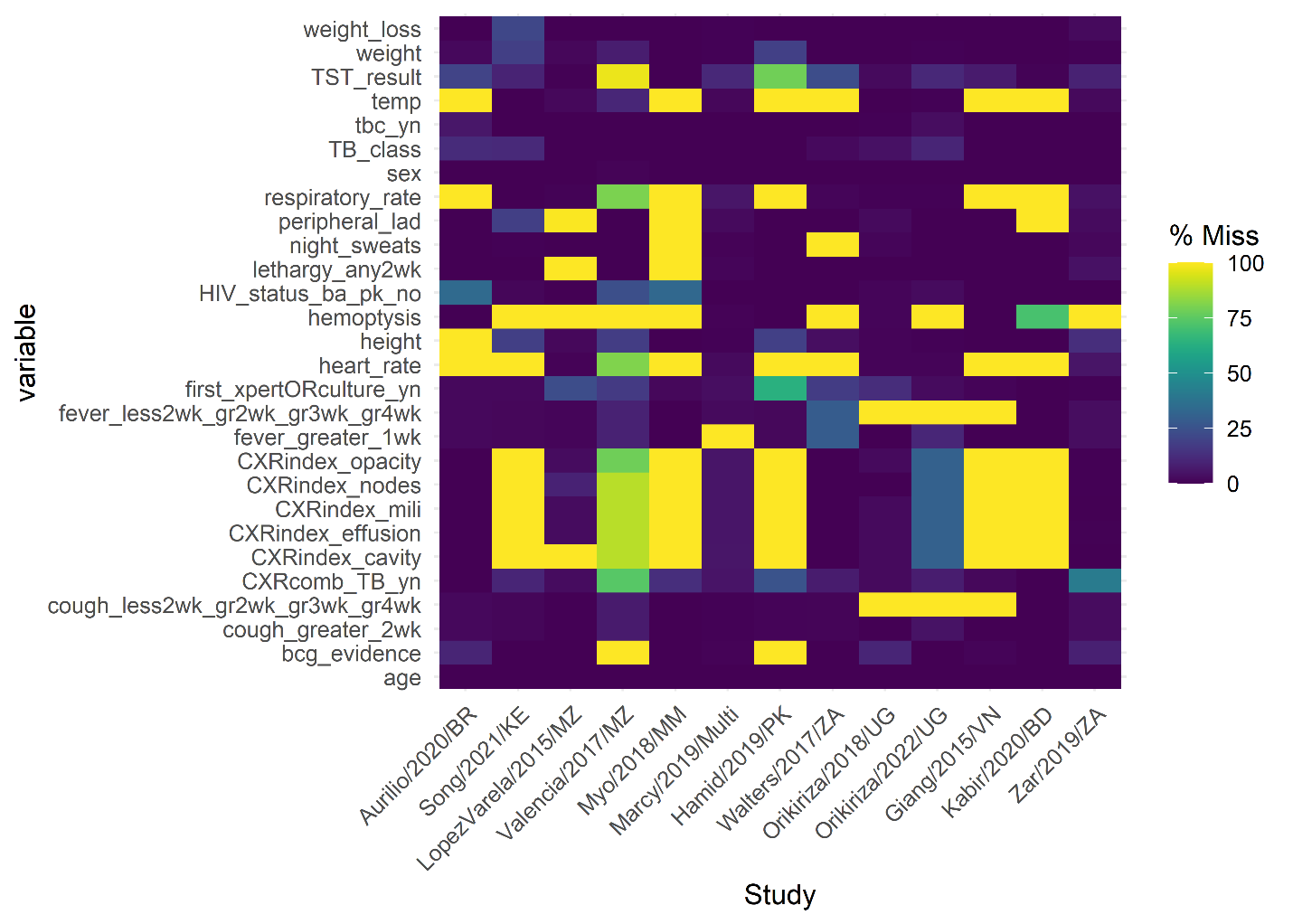

### Appendix K: Generate additional variables

After imputation, additional variables were computed from requested variables as follows:

- **Temperature >38°C**
  - Objective temperature recorded as greater than 38°C
- **Tachycardia**
  - Children <2 months old, heart rate >160
  - Children 2-12 months old, heart rate >150
  - Children 12 months – 5 years old, heart rate >140
  - Children >5 years old, heart rate >120
- **Tachypnoea**
  - Children <2 months old, respiratory rate >60
  - Children 2-12 months old, respiratory rate >50
  - Children 12 months – 5 years old, respiratory rate >40
  - Children >5 years old, respiratory rate >30
- **Weight-for-age Z-score**
  - Determined from sex, age, and weight as per WHO Child Growth Standards
  - Implemented in function “addWGSR” in R package *zscorer*
- **Weight-for-height Z-score**
  - Determined from sex, weight, and height as per WHO Child Growth Standards
  - Implemented in function “addWGSR” in R package *zscorer*
- **Body-mass-index-for-height Z-score**
  - Determined from sex, weight, height, and age as per WHO Child Growth Standards
  - Implemented in function “addWGSR” in R package *zscorer*
- **Severely acutely malnourished**
  - Children <5 years old, weight-for-height Z-score <-3
  - Children ≥5 years old, body-mass-index-for-height Z-score <-3

### Appendix L: Study quality assessment

Table S41: Modified Newcastle-Ottawa Scale for cohort studies^21^

| **Selection** | **Kabir/2020/BD** | **Aurilio/2020/BR** | **Song/2021/KE** | **LopezVarela/2015/MZ** | **Valencia/2017/MZ** | **Myo/2018/MM** | **Marcy/2019/Multi** | **Hamid/2019/PK** | **Zar/2019/ZA** | **Walters/2017/ZA** | **Orikiriza/2018/UG** | **Orikiriza/2022/UG** | **Giang/2015/VN** |
| --- | --- | --- | --- | --- | --- | --- | --- | --- | --- | --- | --- | --- | --- |
| 1) Representativeness of the exposed cohort |  |  |  |  |  |  |  |  |  |  |  |  |  |
| a) truly representative of the target population in the community* |  |  |  |  |  |  |  |  |  |  |  |  |  |
| b) somewhat representative of the target population in the community* | **✓** | **✓** |  | **✓** | **✓** | **✓** | **✓** | **✓** | **✓** | **✓** | **✓** |  | **✓** |
| c) selected group of users |  |  | **✓** |  |  |  |  |  |  |  |  | **✓** |  |
| d) no description of the derivation of the cohort |  |  |  |  |  |  |  |  |  |  |  |  |  |
| 2) Selection of the non-exposed cohort |  |  |  |  |  |  |  |  |  |  |  |  |  |
| a) drawn from the same community as the exposed cohort* | **✓** | **✓** | **✓** | **✓** | **✓** | **✓** | **✓** | **✓** | **✓** | **✓** | **✓** | **✓** | **✓** |
| b) drawn from a different source |  |  |  |  |  |  |  |  |  |  |  |  |  |
| c) no description of the derivation of the non-exposed cohort |  |  |  |  |  |  |  |  |  |  |  |  |  |
| 3) Ascertainment of TB reference classification |  |  |  |  |  |  |  |  |  |  |  |  |  |
| a) secure record* |  |  |  |  |  |  |  |  |  |  |  |  |  |
| b) structured interview* | **✓** | **✓** | **✓** | **✓** | **✓** | **✓** | **✓** | **✓** | **✓** | **✓** | **✓** | **✓** | **✓** |
| c) written self-report |  |  |  |  |  |  |  |  |  |  |  |  |  |
| d) no description |  |  |  |  |  |  |  |  |  |  |  |  |  |
| **Comparability** |  |  |  |  |  |  |  |  |  |  |  |  |  |
| 1) Comparability of cohorts on the basis of the design or analysis | N/A | N/A | N/A | N/A | N/A | N/A | N/A | N/A | N/A | N/A | N/A | N/A | N/A |
| **Outcome** |  |  |  |  |  |  |  |  |  |  |  |  |  |
| 1) Assessment of reference classification |  |  |  |  |  |  |  |  |  |  |  |  |  |
| a) independent assessment blinded to TB reference classification* | **✓** | **✓** | **✓** |  |  | **✓** | **✓** |  | **✓** | **✓** | **✓** | **✓** |  |
| b) record linkage* |  |  |  | **✓** | **✓** |  |  | **✓** |  |  |  |  | **✓** |
| c) self-report |  |  |  |  |  |  |  |  |  |  |  |  |  |
| d) no description |  |  |  |  |  |  |  |  |  |  |  |  |  |
| 2) Follow-up of cohorts to establish reference classification |  |  |  |  |  |  |  |  |  |  |  |  |  |
| a) Follow-up for all participants* |  | **✓** | **✓** |  |  | **✓** | **✓** | **✓** | **✓** | **✓** | **✓** | **✓** | **✓** |
| b) Follow-up for only participants treated for TB | **✓** |  |  | **✓** | **✓** |  |  |  |  |  |  |  |  |
| c) follow-up information not used to establish reference classification |  |  |  |  |  |  |  |  |  |  |  |  |  |
| 3) Adequacy of follow-up of cohorts | N/A | N/A | N/A | N/A | N/A | N/A | N/A | N/A | N/A | N/A | N/A | N/A | N/A |
| **Total** (out of maximum score 5) | **4** | **5** | **4** | **4** | **4** | **5** | **5** | **5** | **5** | **5** | **5** | **4** | **5** |

### Appendix M: Existing algorithms and modifications to make maximal use of IPD

**Table S42. Modifications to Marais et al. criteria^22^**

| **Algorithm** | **Variable in data** | **Differences** |
| --- | --- | --- |
| Persistent, nonremitting cough > 2 weeks | Cough duration | Cannot specify cough characteristic (persistent and nonremitting) |
| Objective weight loss (documented failure to thrive) during the preceding 3 months | Weight loss | Definition of weight loss was not specific to failure to thrive |
| Reported fatigue | Lethargy |  |

**Figure S3. The Union’s desk guide^23^**

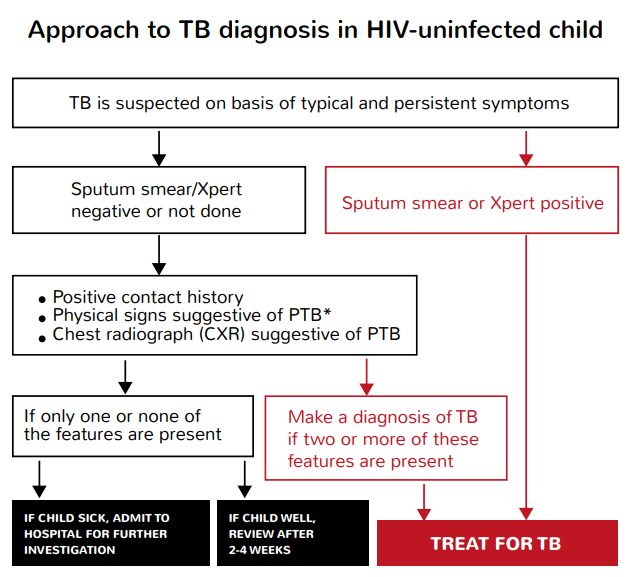

**Table S43. Modifications to The Union’s desk guide**

| **Algorithm** | **Variables in data** | **Differences** |
| --- | --- | --- |
| **Strict Symptom Criteria** |  |  |
| *Persistent, non-remitting cough or wheeze for more than 2 weeks not responding to standard therapy* | Cough duration | Cannot specify cough characteristic (persistent and nonremitting) |
| *Documented loss of weight or failure to thrive during past 3 months especially if not responding to food and/or micronutrient supplementation, or severe malnutrition* | Weight loss  Weight/Height/Age | Definition of weight loss was not specific to failure to thrive  Use weight/height/age to determine if severely acutely malnourished |
| *Fatigue/reduced playfulness* | Lethargy |  |
| *Persistent fever >10 days* | Fever duration | Evaluated as fever >7 days |
| **TB contact in the preceding year** | Documented TB exposure | Some studies defined documented TB exposure as within the previous 24 months |
| **HIV** | HIV status |  |
| **Physical signs** |  |  |
| *Weight loss or poor weight gain, evidence of growth faltering* | Weight loss | Definition of weight loss was not specific to failure to thrive |
| *Fever* | Temperature (C) |  |
| *Increased respiratory rate* | Respiratory rate (per min) |  |
| *Signs of respiratory distress* | N/A | N/A |
| *Auscultation and percussion* | N/A | N/A |
| **CXR** |  |  |
| *Enlarged hilar lymph nodes* | Intrathoracic lymphadenopathy on CXR |  |
| *Opacification in lung tissue* | Opacities on CXR |  |
| *Miliary mottling* | Miliary infiltrate on CXR |  |
| *Cavitation* | Cavities on CXR |  |
| *Pleural or pericardial effusion* | Pleural effusion on CXR | Did not evaluate pericardial effusion |
| *Marked abnormality on CXR in child with no signs of respiratory distress (no fast breathing or chest indrawing) is supportive of TB* | N/A | N/A |
| **Sputum Xpert** | First Xpert MTB/RIF |  |
| **Sputum smear** | N/A | N/A |

**Figure S4. Stegen-Toledo Score^24^**

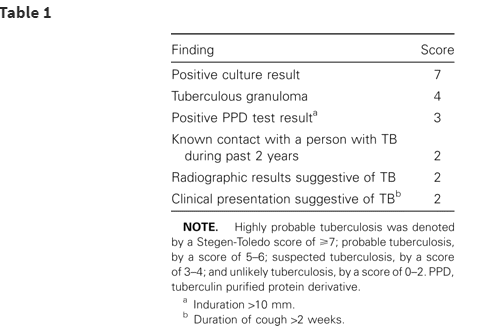

**Table S44. Modifications to Stegen-Toledo Score** (using cutoff of 5 points to classify TB)

| **Algorithm** | **Variables in data** | **Differences** |
| --- | --- | --- |
| Positive culture result | First Xpert MTB/RIF | Used Xpert rather than culture given practical advantage of Xpert |
| Tuberculosis granuloma | N/A | N/A |
| Positive PPD test result | TST result |  |
| Known contact with a person with TB during past 2 years | Documented TB exposure | Some studies defined documented TB exposure as within the previous 12 months |
| Radiological results suggestive of TB | CXR consistent with TB |  |
| Clinical presentation suggestive of TB (defined as duration of cough >2 weeks) | Cough duration |  |

**Figure S5. Uganda National Tuberculosis and Leprosy Control Programme (NTLP) algorithm^25^**

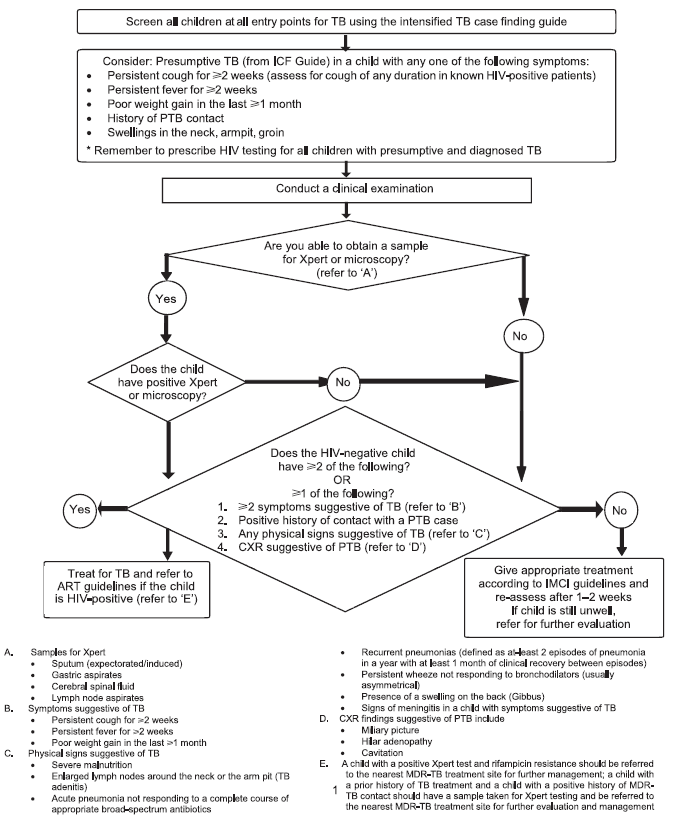

**Table S45. Modifications to Uganda NTLP Algorithm**

| **Algorithm** | **Variables in data** | **Differences** |
| --- | --- | --- |
| **Xpert or microscopy** | First Xpert MTB/RIF | Did not evaluate microscopy |
| **Symptoms suggestive of TB** (*≥2* of the following) |  |  |
| *Persistent cough ≥2 wks* | Cough duration |  |
| *Persistent fever for ≥2 wks* | Fever duration |  |
| *Poor weight gain in the last ≥1 month* | Weight loss |  |
| **CXR findings suggestive of PTB** |  |  |
| *Miliary picture* | Miliary infiltrate on CXR |  |
| *Hilar adenopathy* | Intrathoracic lymphadenopathy on CXR |  |
| *Cavitation* | Cavities on CXR |  |
| **Physical signs suggestive of TB** |  |  |
| *Severe malnutrition* | Weight/Height/Age | Use weight/height/age to determine if severely acutely malnourished |
| *Enlarged lymph nodes around neck or arm pit* | Peripheral lymphadenopathy |  |
| *Acute pneumonia not responding to complete course of appropriate antibiotics* | N/A | N/A |
| *Recurrent pneumonias* | N/A | N/A |
| *Persistent wheeze not responding to bronchodilators* | N/A | N/A |
| *Persistence of swelling on the back (Gibbus)* | N/A | N/A |
| *Signs of meningitis in child with symptoms suggestive of TB* | N/A | N/A |

**Figure S6. Brazilian Ministry of Health Score^26^**

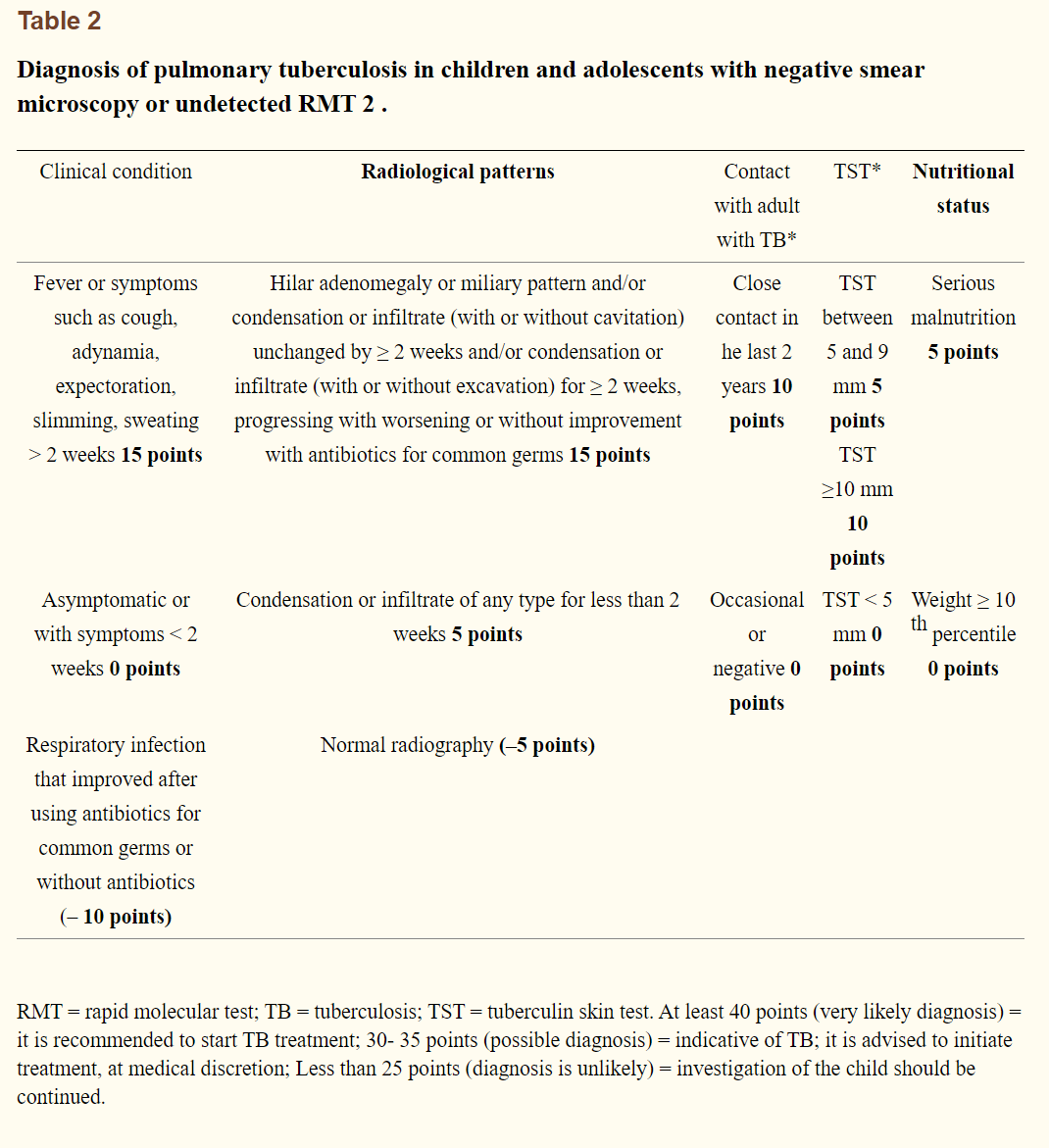

**Table S46. Modifications to Brazilian Ministry of Health Score** (using cutoff of 30 points to classify TB)

| **Algorithm** | **Variables in data** | **Differences** |
| --- | --- | --- |
| Fever *≥2 wks* | Fever duration |  |
| Cough *≥2 wks* | Cough duration |  |
| Adynamia *≥2 wks* | Lethargy | Duration not specified in data |
| Expectoration *≥2 wks* | N/A | N/A |
| Slimming *≥2 wks* | Weight loss | Duration not specified in data |
| Sweating *≥2 wks* | Night sweats | Duration not specified in data |
| Respiratory infection that improved after using antibiotics for common germs or without antibiotics | N/A |  |
| Hilar adenomegaly or miliary pattern and/or condensation or infiltrate (with or without cavitation) unchanged by ≥ 2 weeks and/or condensation or infiltrate (with or without excavation) for ≥ 2 weeks, progressing with worsening or without improvement with antibiotics for common germs | Intrathoracic lymphadenopathy on CXR  Miliary infiltrate on CXR  Opacities on CXR  Cavities on CXR | CXR abnormalities consistent with TB of unknown duration |
| Condensation or infiltrate of any type for less than 2 weeks | N/A | No data on duration of CXR abnormalities consistent with TB |
| Normal radiography | N/A | No data to indicate CXR |
| Close contact in the last 2 years (with adult with TB) | Documented TB exposure | Some studies defined documented TB exposure as within the previous 12 months |
| TST diameter | TST result | TST diameter not specified in data, only whether result was positive or not |
| Serious malnutrition (weight <10^th^ percentile) | Weight/age | Weight and age used to compute weight-for-age z-score |

**Figure S7. Keith-Edwards Score^27^**

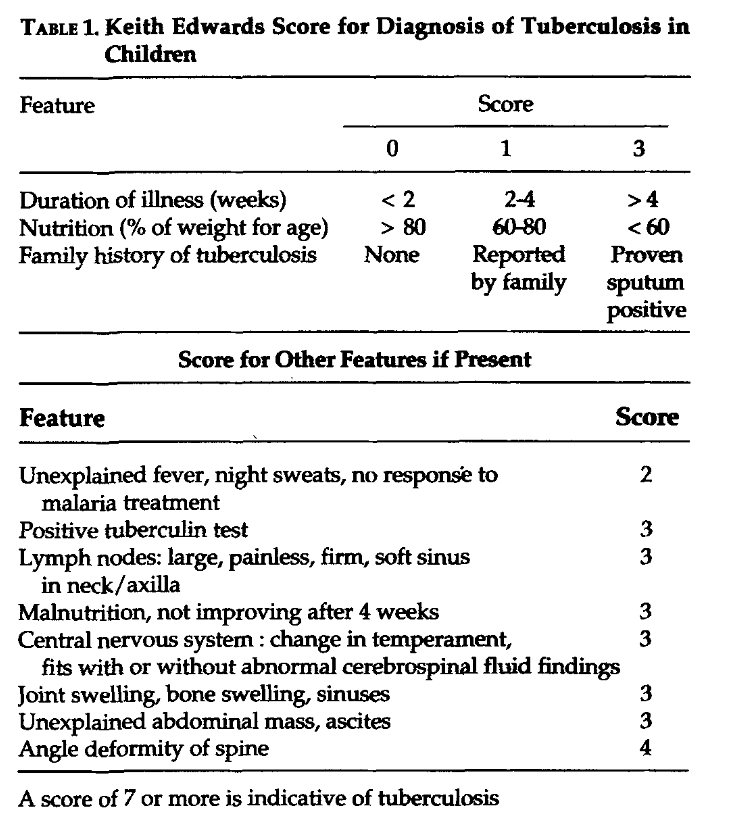

**Table S47. Modifications to Keith-Edwards Score**

| **Algorithm** | **Variables in Data** | **Differences** |
| --- | --- | --- |
| Duration of illness | Cough duration  Fever duration | Used the max of cough or fever duration to represent duration of illness |
| Nutrition (% of weight for age) | Weight/age | Weight and age used to compute weight-for-age z-score |
| Family history of tuberculosis | Documented TB exposure | Data unavailable on whether TB exposure was bacteriologically-confirmed |
| Fever | Temperature (C) |  |
| Night sweats | Night sweats |  |
| No response to malaria treatment | N/A | N/A |
| Lymph nodes: large, painless, firm, soft sinus in neck/axilla | Peripheral lymphadenopathy |  |
| Malnutrition, not improving after 4 weeks | Weight loss | Cannot specify whether malnutrition did not improve after 4 weeks |
| Central nervous system: change in temperament, fits with or without abnormal cerebrospinal fluid findings | Lethargy | Unable to evaluate fits or abnormal cerebrospinal fluid findings |
| Joint swelling, bone swelling, sinuses | N/A | N/A |
| Unexplained abdominal mass, ascites | N/A | N/A |
| Angle deformity of spine | N/A | N/A |

**Figure S8. Gunasekera et al. algorithm^28^**

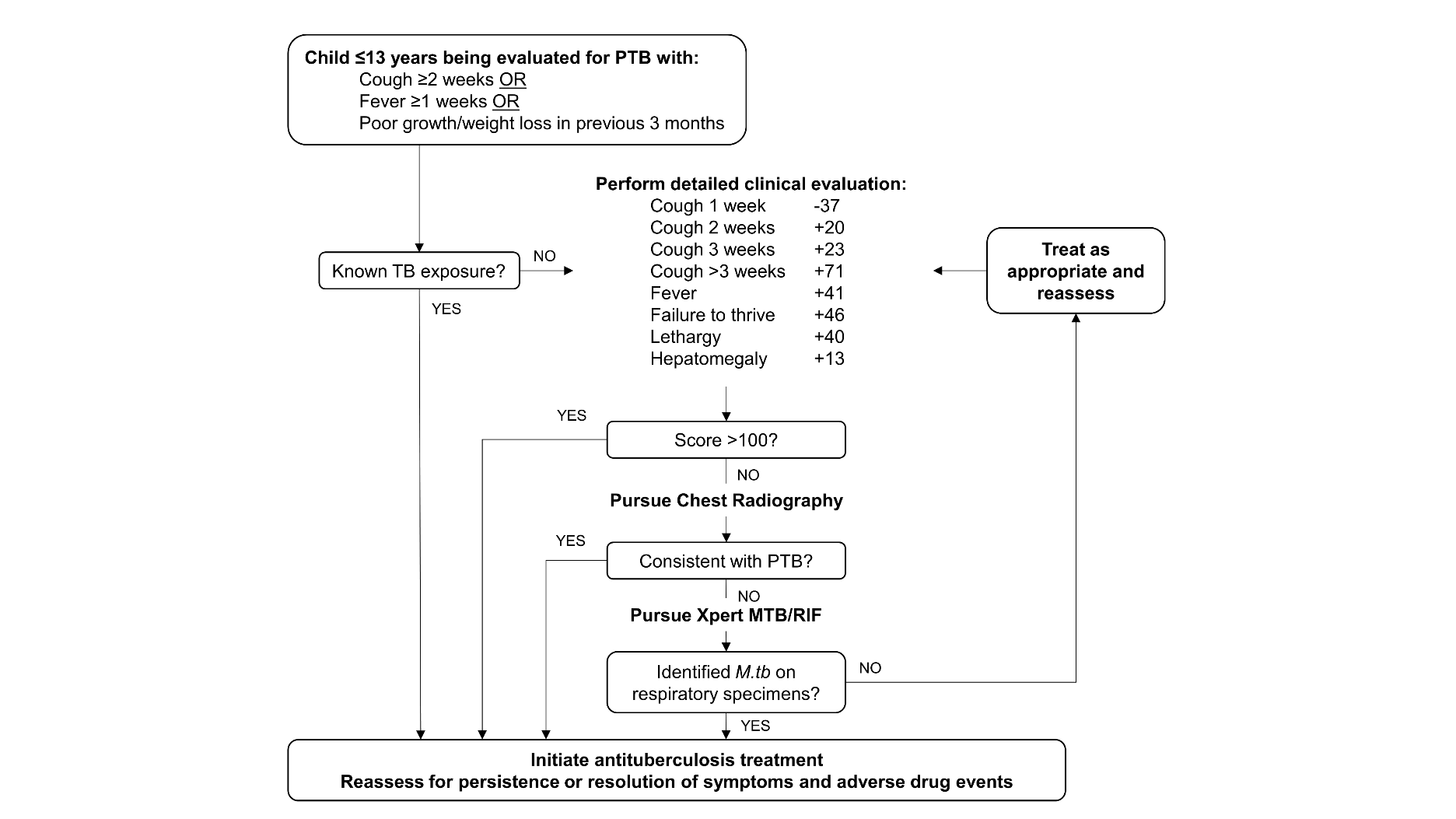

**Table S48. Modifications to Gunasekera et al. Algorithm.** Given that we had access to the Walters/2017/ZA data from HIV-negative children used to develop this algorithm, we refit the logistic regression model using a complete case analysis of variables available in the IPD before any imputation using the composite TB reference standard; thus, this algorithm is modified from the originally stated algorithm (we do not include hepatomegaly and fever is defined as ≥1 week). This model had an AUC of 0·85. The model parameter coefficients were scaled to produce a score such that a sum of the scores resulted in classification of TB with a sensitivity of 90% -- this resulted in an algorithm with a sensitivity of 91% and a specificity of 49%.

|  | OR | 2·50% | 97·5% | p-value | Scaled score |
| --- | --- | --- | --- | --- | --- |
| No cough | -- | -- | -- | -- | -- |
| Cough < 2 weeks | 0·81 | 0·37 | 1·78 | 0·60 | -22 |
| Cough 2 weeks | 1·14 | 0·40 | 3·18 | 0·81 | 13 |
| Cough 3 weeks | 1·56 | 0·46 | 5·25 | 0·47 | 46 |
| Cough >3 weeks | 3·60 | 1·49 | 9·01 | 0·01 | 132 |
| No fever or fever <1 week | -- | -- | -- | -- | -- |
| Fever ≥1 week | 2·12 | 0·87 | 5·33 | 0·10 | 78 |
| No weight loss | -- | -- | -- | -- | -- |
| Weight loss | 1·98 | 1·06 | 3·76 | 0·03 | 71 |
| No lethargy | -- | -- | -- | -- | -- |
| Lethargy | 1·43 | 0·71 | 2·88 | 0·32 | 37 |
| No history of documented TB contact | -- | -- | -- | -- | -- |
| History of documented TB contact | 6·64 | 3·53 | 12·99 | 0·00 | 195 |
| CXR not consistent with TB | -- | -- | -- | -- | -- |
| CXR consistent with TB | 11·02 | 5·39 | 23·90 | 0·00 | 248 |
| Xpert negative for Mtb | -- | -- | -- | -- | -- |
| Xpert positive for Mtb | 13927274·15 | 0·00 | Inf | 0·98 | 1698 |

**Figure S9. Marcy et al. Algorithm^14^**

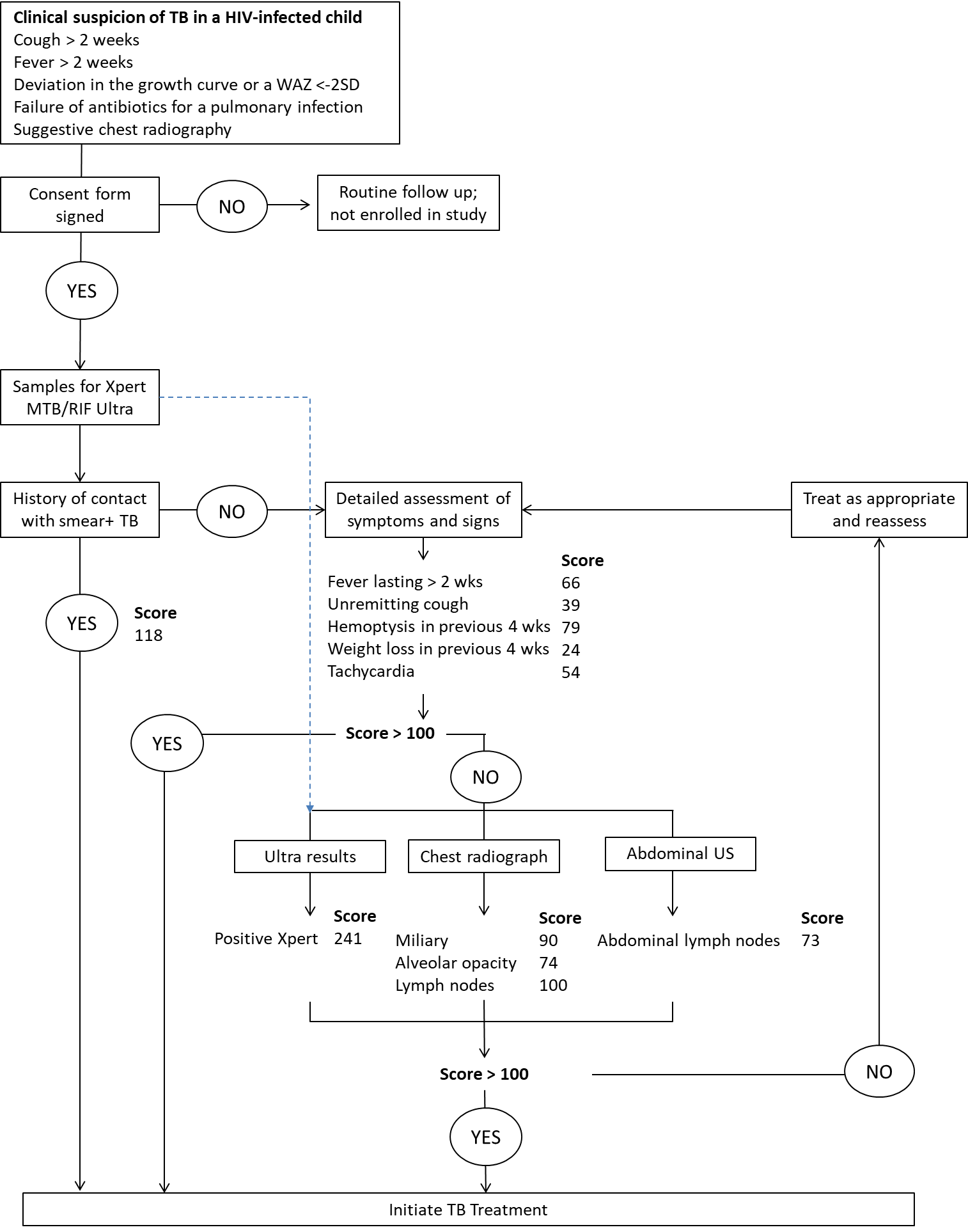

**Table S49. Modifications to Marcy et al. Algorithm**. Given that we had access to the Marcy/2019/Multi data from HIV-positive children used to develop this algorithm, we refit the logistic regression model using a complete case analysis of variables available in the IPD before any imputation using the composite TB reference standard; thus, this algorithm is modified from the originally stated algorithm (we use cough ≥2 weeks rather than remitting cough, and we do not include abdominal ultrasound results). This model had an AUC of 0·80. The model parameter coefficients were scaled to produce a score such that a sum of the scores resulted in classification of TB with a sensitivity of 90% -- this resulted in an algorithm with a sensitivity of 91% and a specificity of 40%.

|  | **odds-ratio** | **2·50%** | **97·5%** | **p-value** | **Scaled score** |
| --- | --- | --- | --- | --- | --- |
| No cough or cough <2 weeks | -- | -- | -- | -- | -- |
| Cough ≥2 weeks | 1·11 | 0·52 | 2·37 | 0·78 | 9 |
| No fever or fever <1 week | -- | -- | -- | -- | -- |
| Fever ≥1 week | 2·94 | 1·72 | 5·39 | 0·00 | 95 |
| No weight loss | -- | -- | -- | -- | -- |
| Weight loss | 1·79 | 1·01 | 3·37 | 0·05 | 52 |
| No haemoptysis | -- | -- | -- | -- | -- |
| Haemoptysis | 3·29 | 0·62 | 93·23 | 0·23 | 105 |
| No tachycardia | -- | -- | -- | -- | -- |
| Tachycardia | 2·03 | 0·91 | 5·12 | 0·09 | 62 |
| No history of documented TB contact | -- | -- | -- | -- | -- |
| History of documented TB contact | 1·71 | 0·60 | 5·59 | 0·33 | 47 |
| Miliary infiltrate not present on CXR | -- | -- | -- | -- | -- |
| Miliary infiltrate present on CXR | 2·56 | 0·77 | 10·36 | 0·14 | 83 |
| Opacities not present on CXR | -- | -- | -- | -- | -- |
| Opacities present on CXR | 2·36 | 1·32 | 4·53 | 0·00 | 76 |
| Intrathoracic lymphadenopathy not present on CXR | -- | -- | -- | -- | -- |
| Intrathoracic lymphadenopathy present on CXR | 5·41 | 2·84 | 11·83 | 0·00 | 149 |
| Xpert negative for Mtb | -- | -- | -- | -- | -- |
| Xpert positive for Mtb | 29·18 | 3·40 | Inf | 0·03 | 298 |

### Appendix N: Performance of existing algorithms against reference classification of all TB

**Figure S10. Performance of Marais et al. criteria.** Study-level and pooled estimates of the **(A)** sensitivity and **(B)** specificity of classifying TB (composite reference standard: bacteriologically-confirmed pulmonary TB and unconfirmed pulmonary TB).

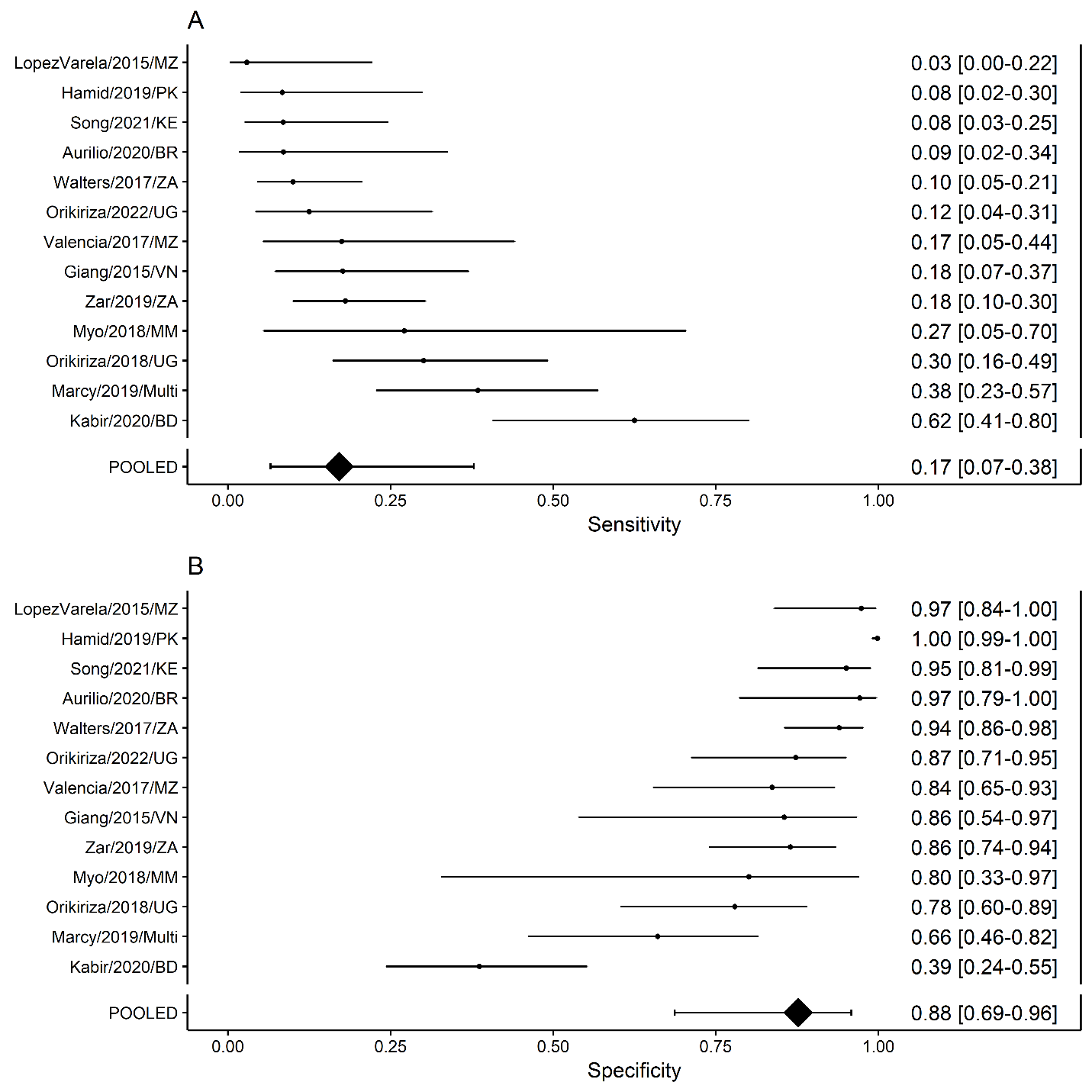

**Figure S11. Performance of Stegen-Toledo Score** (using cutoff of 5 points to classify TB). Study-level and pooled estimates of the (A) sensitivity and (B) specificity of classifying TB (composite reference standard: bacteriologically-confirmed pulmonary TB and unconfirmed pulmonary TB).

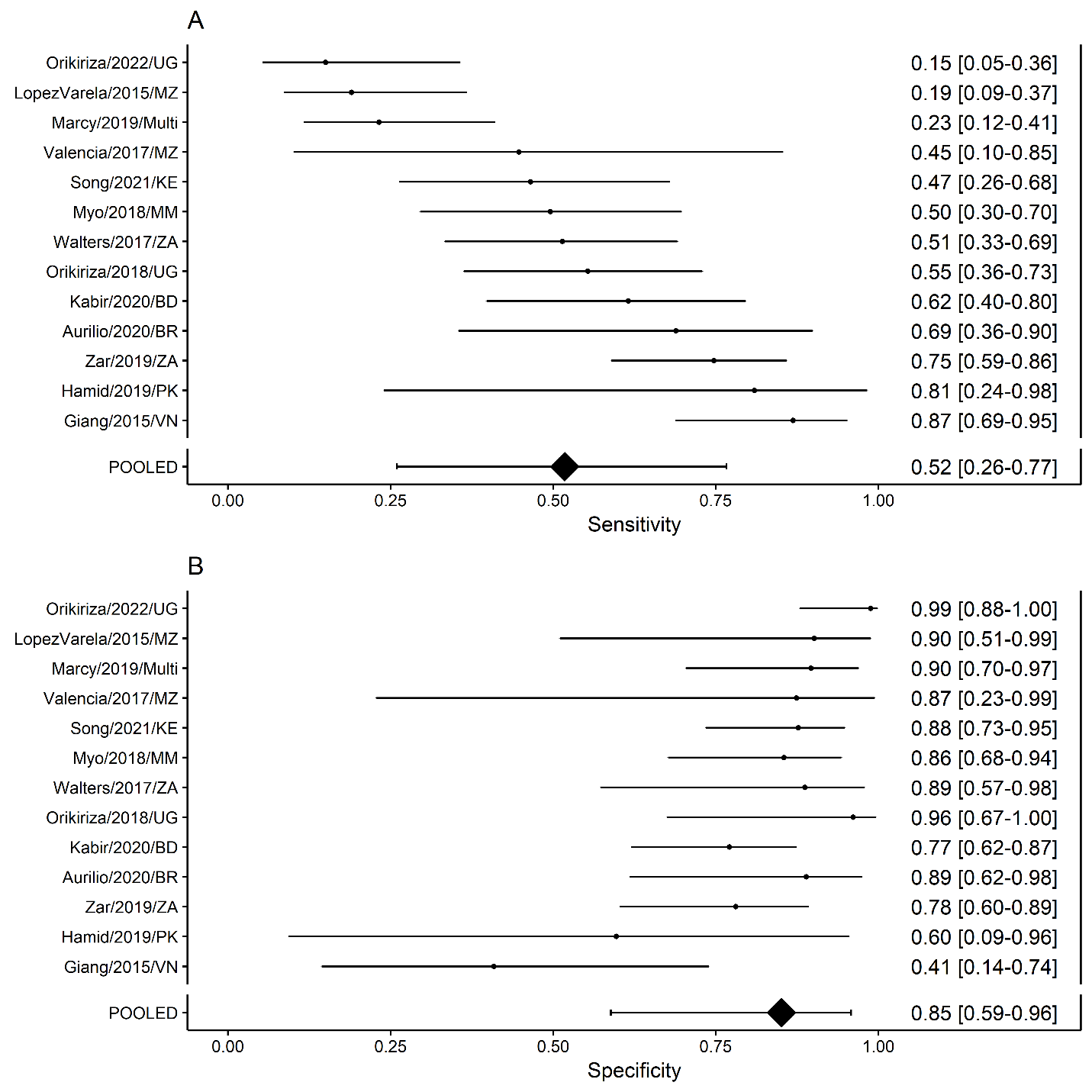

**Figure S12. Performance of Uganda NTLP Algorithm**. Study-level and pooled estimates of the **(A)** sensitivity and **(B)** specificity of classifying TB (composite reference standard: bacteriologically-confirmed pulmonary TB and unconfirmed pulmonary TB).

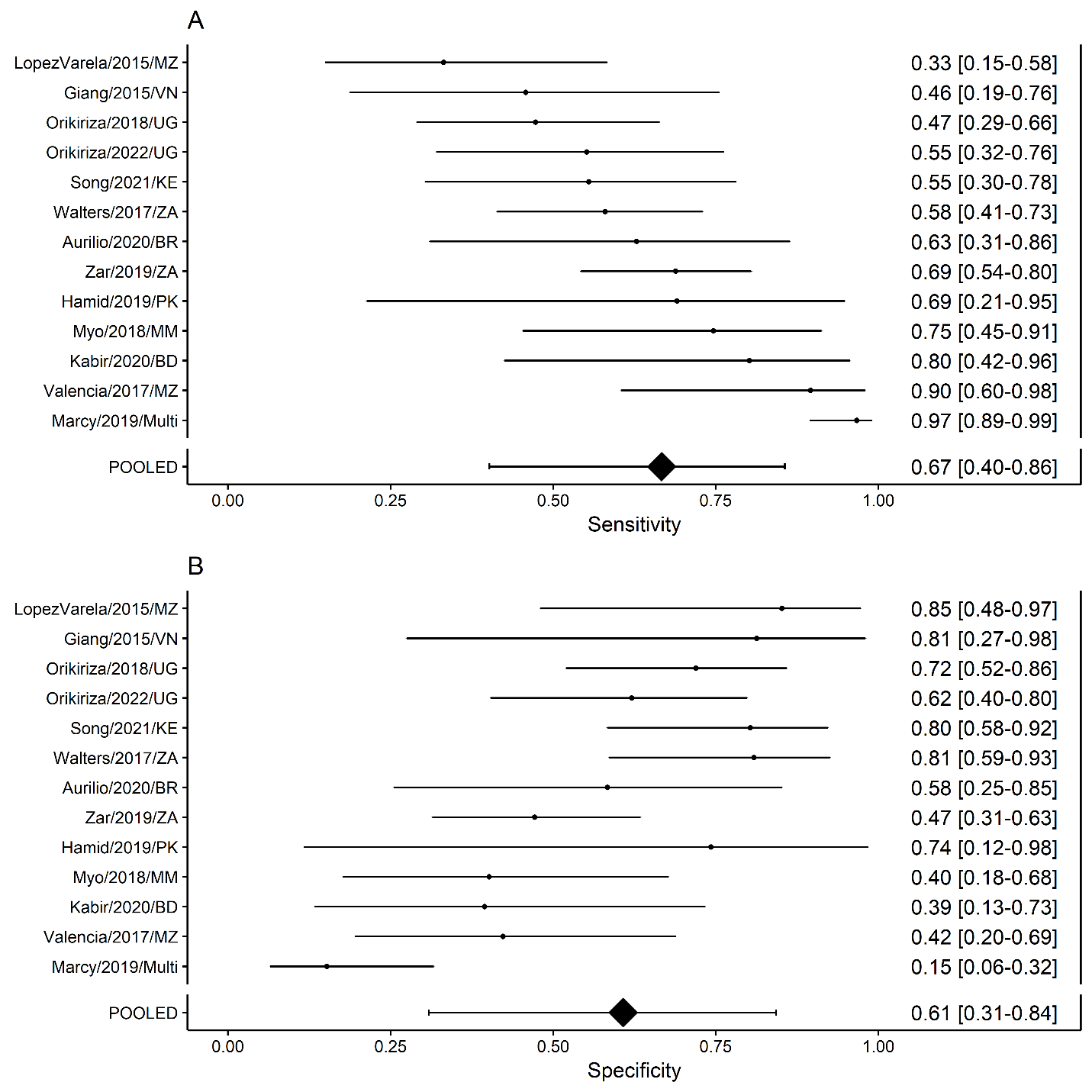

**Figure S13. Performance of The Union’s Desk Guide.** Study-level and pooled estimates of the **(A)** sensitivity and **(B)** specificity of classifying TB (composite reference standard: bacteriologically-confirmed pulmonary TB and unconfirmed pulmonary TB).

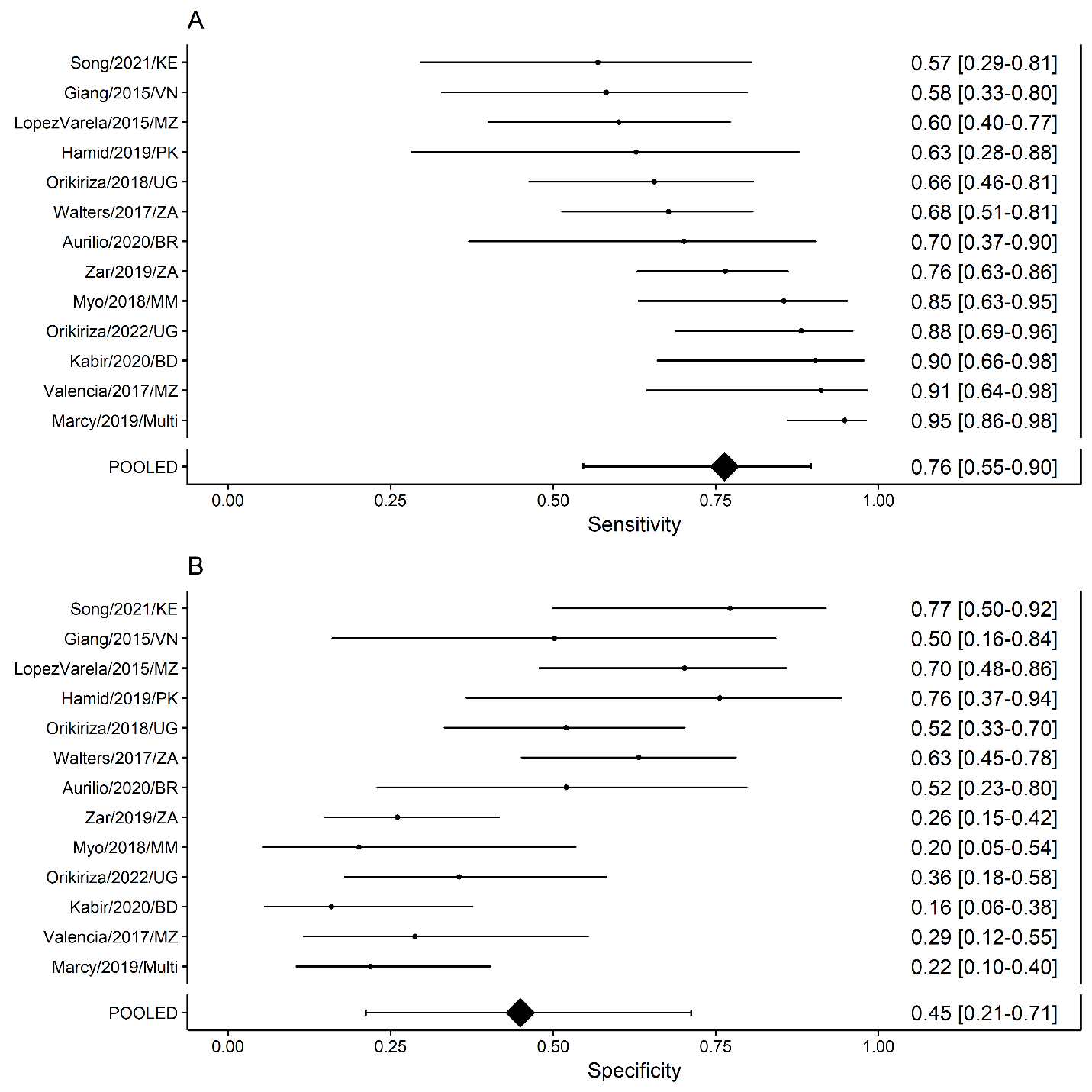

**Figure S14. Performance of Brazilian Ministry of Health Score (**using cutoff of 30 points to classify TB) Study-level and pooled estimates of the **(A)** sensitivity and **(B)** specificity of classifying TB (composite reference standard: bacteriologically-confirmed pulmonary TB and unconfirmed pulmonary TB).

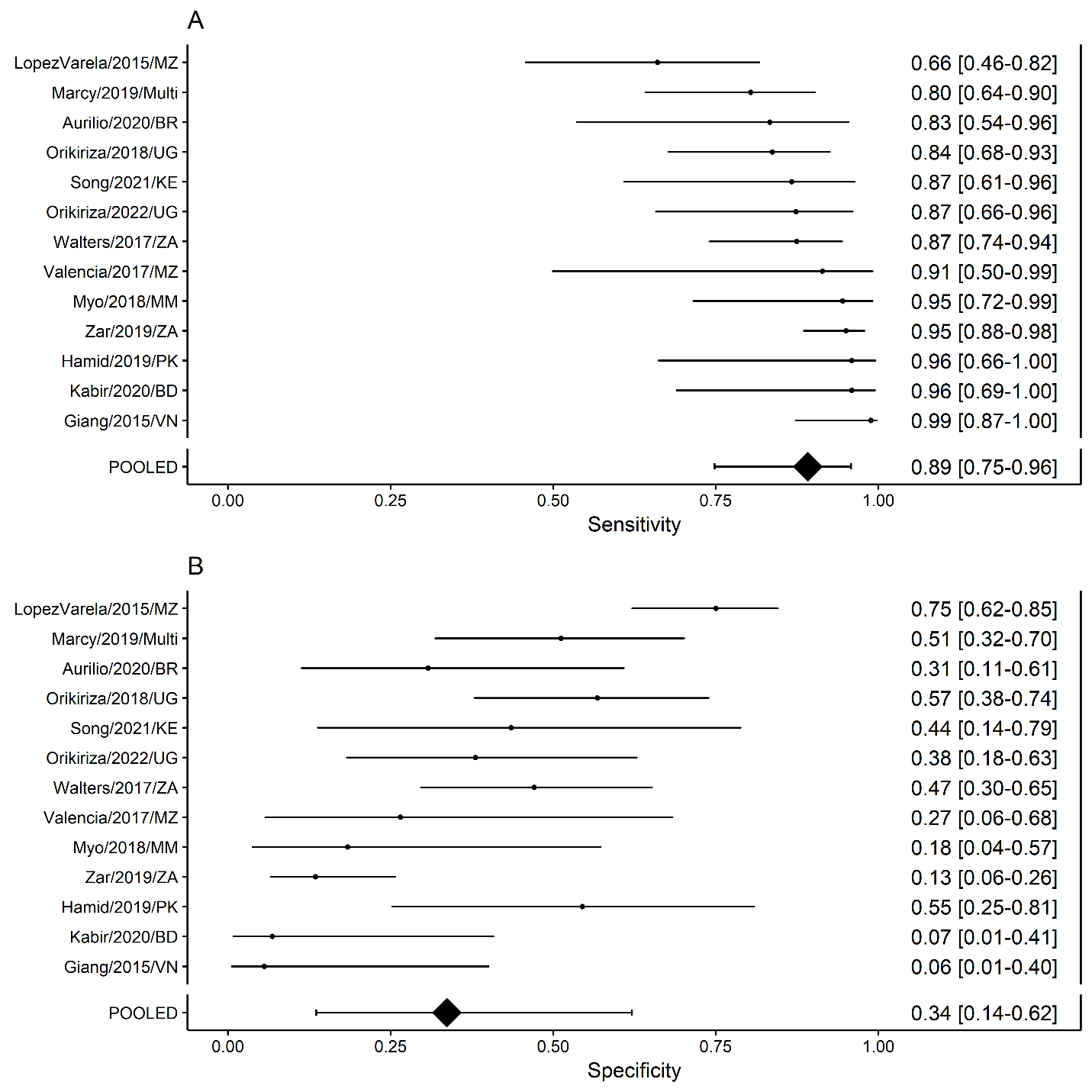

**Figure S15. Performance of Keith-Edwards Score.** Study-level and pooled estimates of the **(A)** sensitivity and **(B)** specificity of classifying TB (composite reference standard: bacteriologically-confirmed pulmonary TB and unconfirmed pulmonary TB).

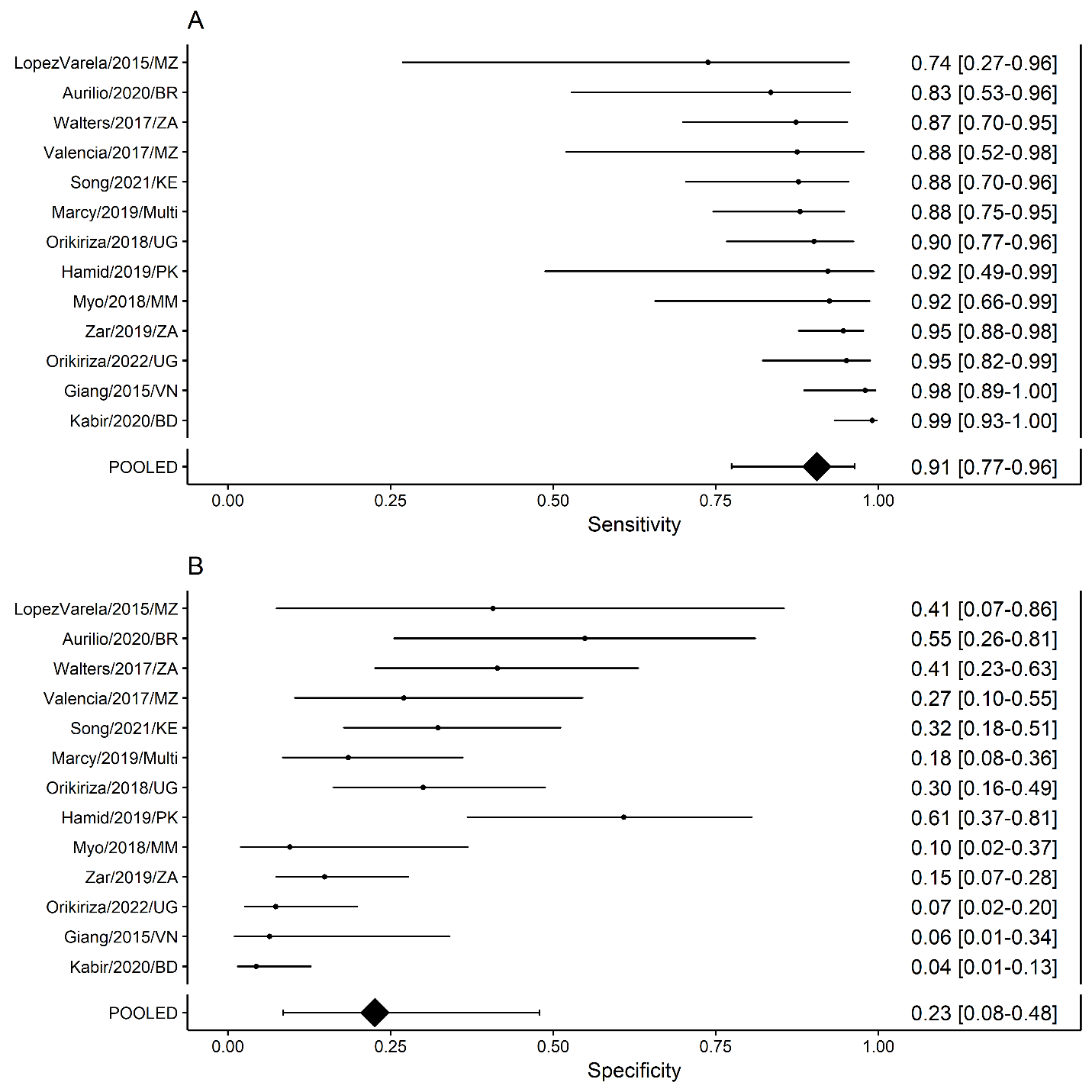

**Figure S16. Performance of Marcy et al. Algorithm.** Performance estimates of the Marcy et al. Algorithm were derived from only HIV-positive children in the IPD that excludes data form the Marcy/2019/Multi cohort (from which the algorithm was developed). Study-level and pooled estimates of the **(A)** sensitivity and **(B)** specificity of classifying TB (composite reference standard: bacteriologically-confirmed pulmonary TB and unconfirmed pulmonary TB).

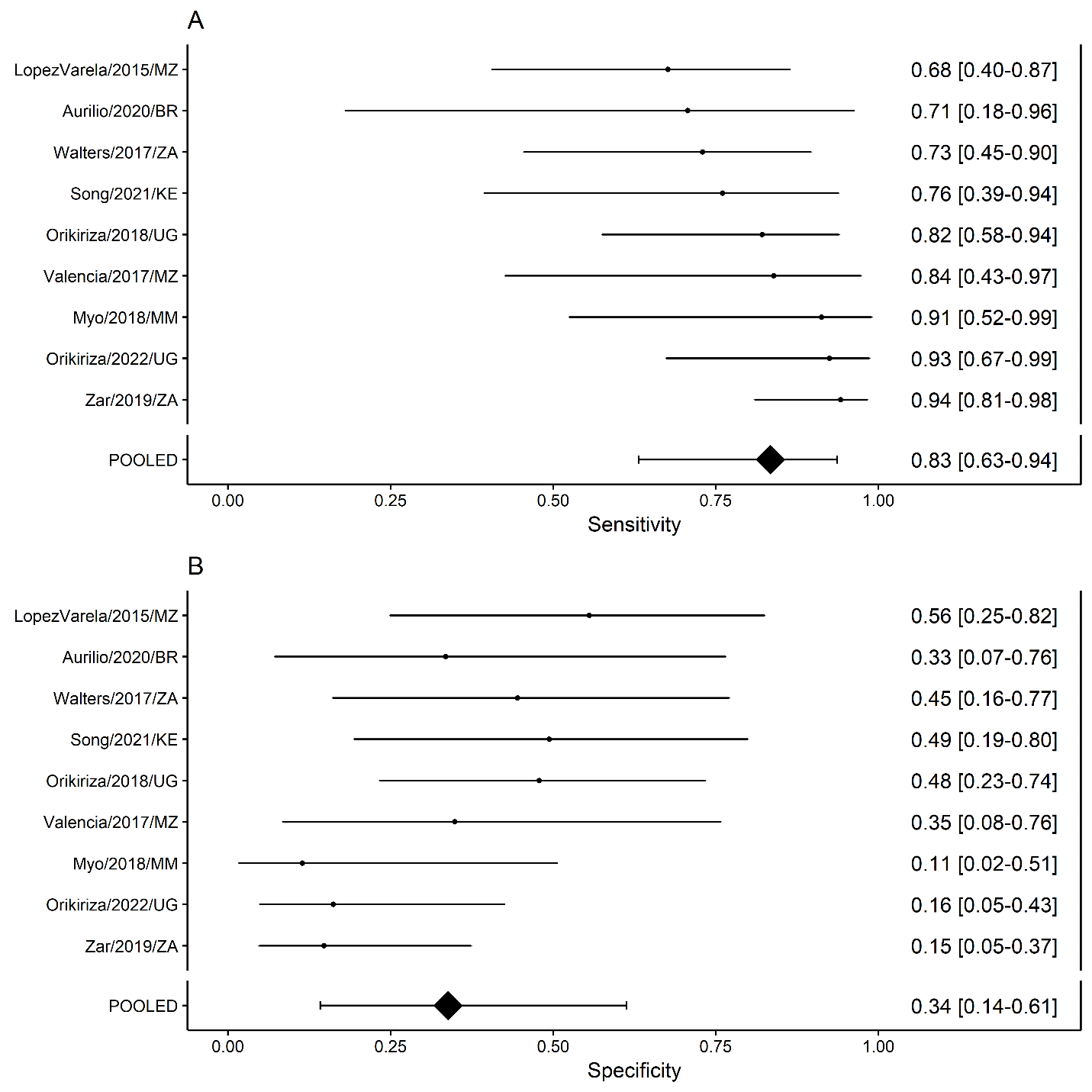

**Figure S17. Performance of Gunasekera et al. Algorithm.** Performance estimates of the Gunasekera et al. Algorithm were derived from only HIV-negative children in the IPD that excludes data from the Walter/2017/ZA population (from which the algorithm was developed). Study-level and pooled estimates of the **(A)** sensitivity and **(B)** specificity of classifying TB (composite reference standard: bacteriologically-confirmed pulmonary TB and unconfirmed pulmonary TB).

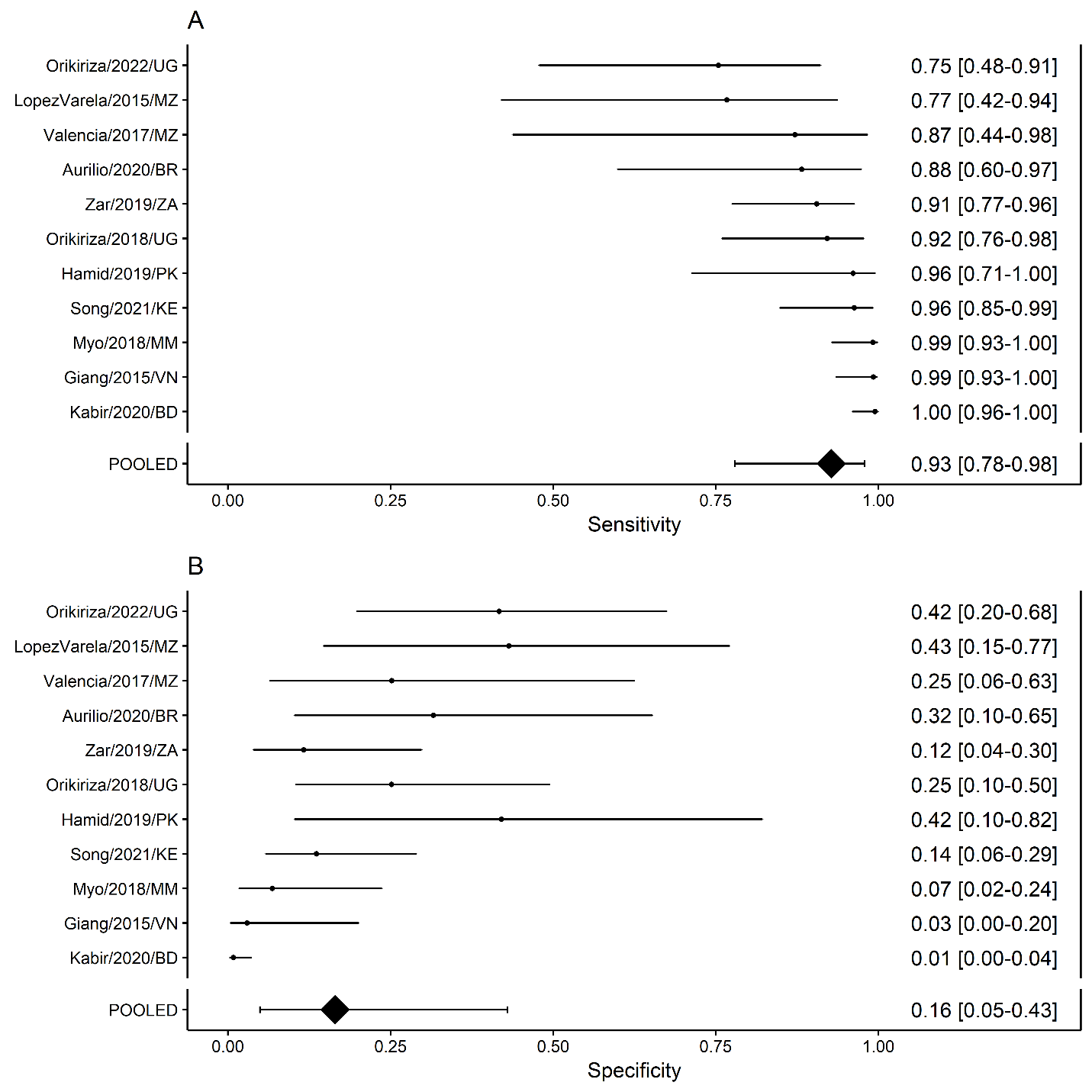

### Appendix O: Performance of existing algorithms against reference classification of bacteriologically-confirmed TB

**Figure S18. Performance of existing algorithms at classifying confirmed TB (excluding data from children with unconfirmed TB).** Retrospective estimates of the pooled **(A)** sensitivity and **(B)** specificity of eight algorithms to guide decisions to treat children with presumptive pulmonary TB, had they been used to evaluate the children for whom we have IPD records. The reference classification of pulmonary TB included bacteriologically-confirmed pulmonary TB only (children with unconfirmed TB are excluded from this analysis).

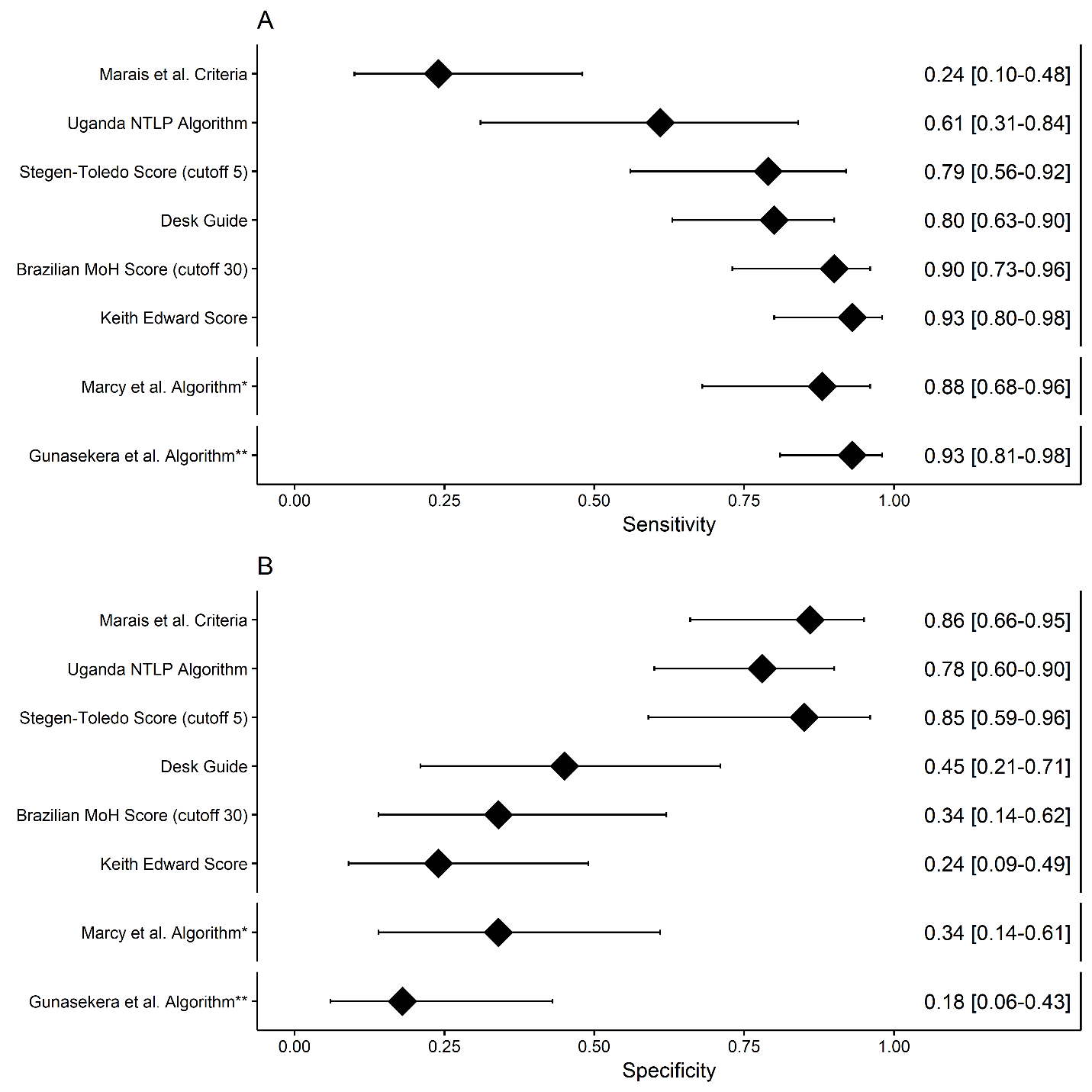

### Appendix P: Calibration and discrimination of prediction model to classify TB

**Figures S19. Forest plot depicting discrimination and calibration of prediction model including chest x-ray features to classify TB.** Study-level and pooled estimates of the **(A)** discrimination (c-statistic) and **(B)** calibration (O:E slope) of the prediction model developed from the IPD in classifying TB using an internal-external cross-validation framework (reference standard: bacteriologically-confirmed pulmonary TB and unconfirmed pulmonary TB). c-Statistic – concordance statistic, O:E – observed: expected slope, IPD – individual participant data, TB – tuberculosis, BD – Bangladesh, BR – Brazil, KE – Kenya, MM – Myanmar, Multi – Multi-country study (includes Burkina Faso, Cameroon, Vietnam, and Cambodia), MZ – Mozambique, PK – Pakistan, UG – Uganda, VN – Vietnam, ZA – South Africa.

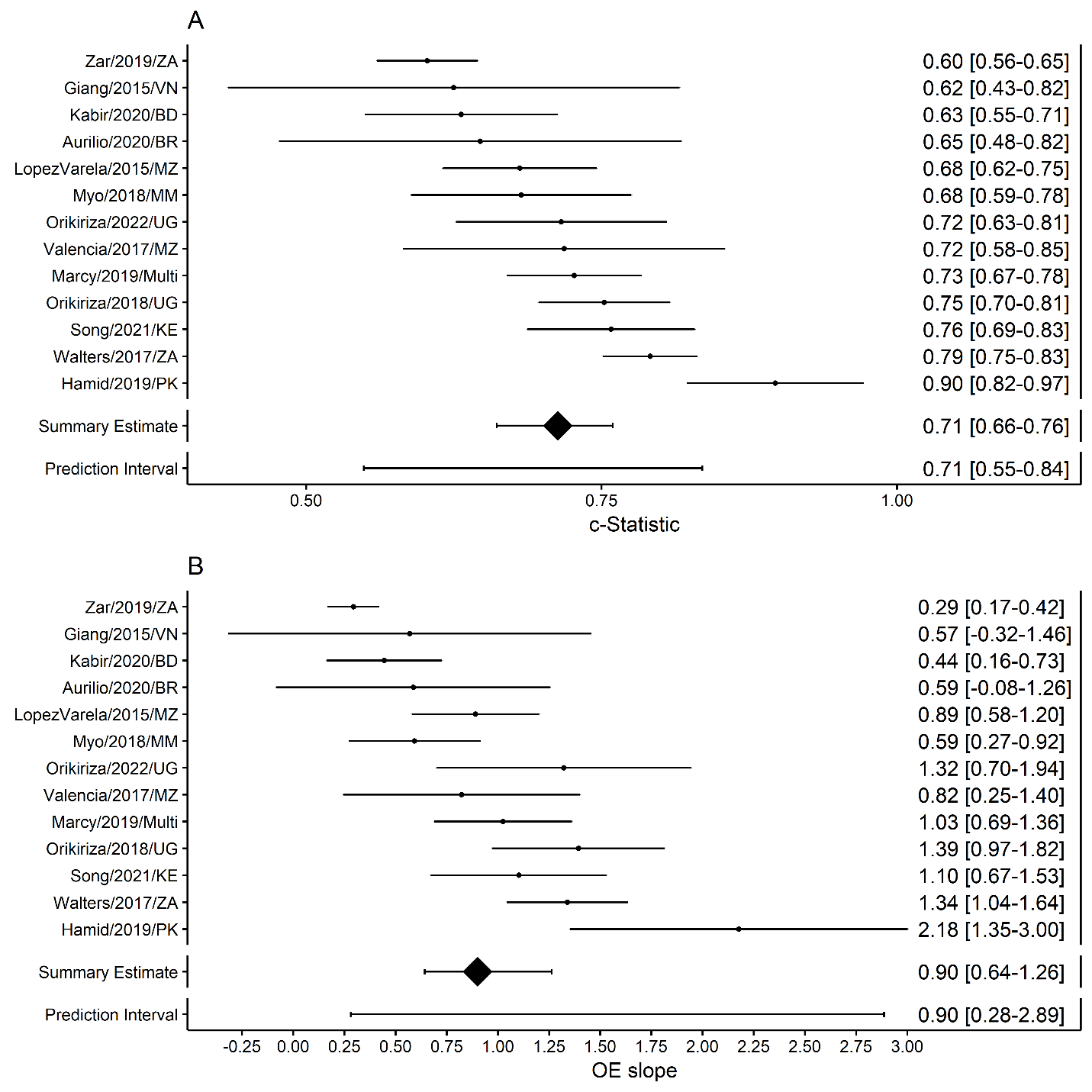

### Appendix Q: Logistic regression model, calibration, and discrimination of prediction model developed from IPD without CXR features

**Table S50. Estimates of logistic regression prediction model developed from IPD without CXR features.** Parameter estimate with 95% confidence intervals, odds ratio estimate with 95% confidence interval, and p-values for each parameter included in the logistic regression prediction model. The estimate provided for each predictor is computed against a reference that reflects the absence of that feature. The model parameter estimates account for potential clustering at the study-level as well as uncertainty introduced by missing data. *P-value calculated using Rubin’s rules for multiple imputed data. TB – tuberculosis, IPD – individual participant data, OR – odds ratio, CI – confidence interval, CXR – chest x-ray.

| **Predictors** | **Coefficient** | **Coefficient 95% CI** | **OR** | **OR 95% CI** | **P-value*** |
| --- | --- | --- | --- | --- | --- |
| Intercept | -1·36 | -1·94, -0·78 | -- | -- | -- |
| Cough duration ≥ 2 weeks  (Absence is no cough or cough <2 weeks) | 0·22 | -0·03, 0·47 | 1·25 | 0·97, 1·6 | 0·75 |
| Fever duration ≥ 2 weeks  (Absence is no fever or fever <2 weeks) | 0·46 | 0·18, 0·73 | 1·58 | 1·2, 2·07 | 0·21 |
| Lethargy | 0·2 | -0·01, 0·41 | 1·22 | 0·99, 1·51 | 0·75 |
| Weight loss | 0·24 | 0·01, 0·48 | 1·28 | 1·01, 1·62 | 0·68 |
| History of documented TB exposure | 1·33 | 0·81, 1·84 | 3·76 | 2·24, 6·31 | <0·001 |
| Haemoptysis | 0·4 | -0·27, 1·06 | 1·49 | 0·76, 2·89 | 0·70 |
| Night sweats | 0·28 | 0·12, 0·45 | 1·33 | 1·12, 1·57 | 0·43 |
| Peripheral lymphadenopathy | 0·32 | 0·12, 0·52 | 1·38 | 1·13, 1·68 | 0·40 |
| Temperature ≥38 degrees Celsius | 0·01 | -0·22, 0·23 | 1·01 | 0·8, 1·26 | >0·999 |
| Tachycardia | 0·19 | -0·08, 0·47 | 1·21 | 0·92, 1·6 | 0·83 |
| Tachypnoea | 0·07 | -0·18, 0·33 | 1·08 | 0·84, 1·39 | 0.97 |

**Figures S20. Forest plot depicting discrimination and calibration of prediction model without chest x-ray features to classify TB.** Study-level and pooled estimates of the **(A)** discrimination (c-statistic) and **(B)** calibration (O:E slope) of the prediction model developed from the IPD in classifying TB using an internal-external cross-validation framework (reference standard: bacteriologically-confirmed pulmonary TB and unconfirmed pulmonary TB). c-Statistic – concordance statistic, O:E – observed: expected slope, IPD – individual participant data, TB – tuberculosis, BD – Bangladesh, BR – Brazil, KE – Kenya, MM – Myanmar, Multi – Multi-country study (includes Burkina Faso, Cameroon, Vietnam, and Cambodia), MZ – Mozambique, PK – Pakistan, UG – Uganda, VN – Vietnam, ZA – South Africa.

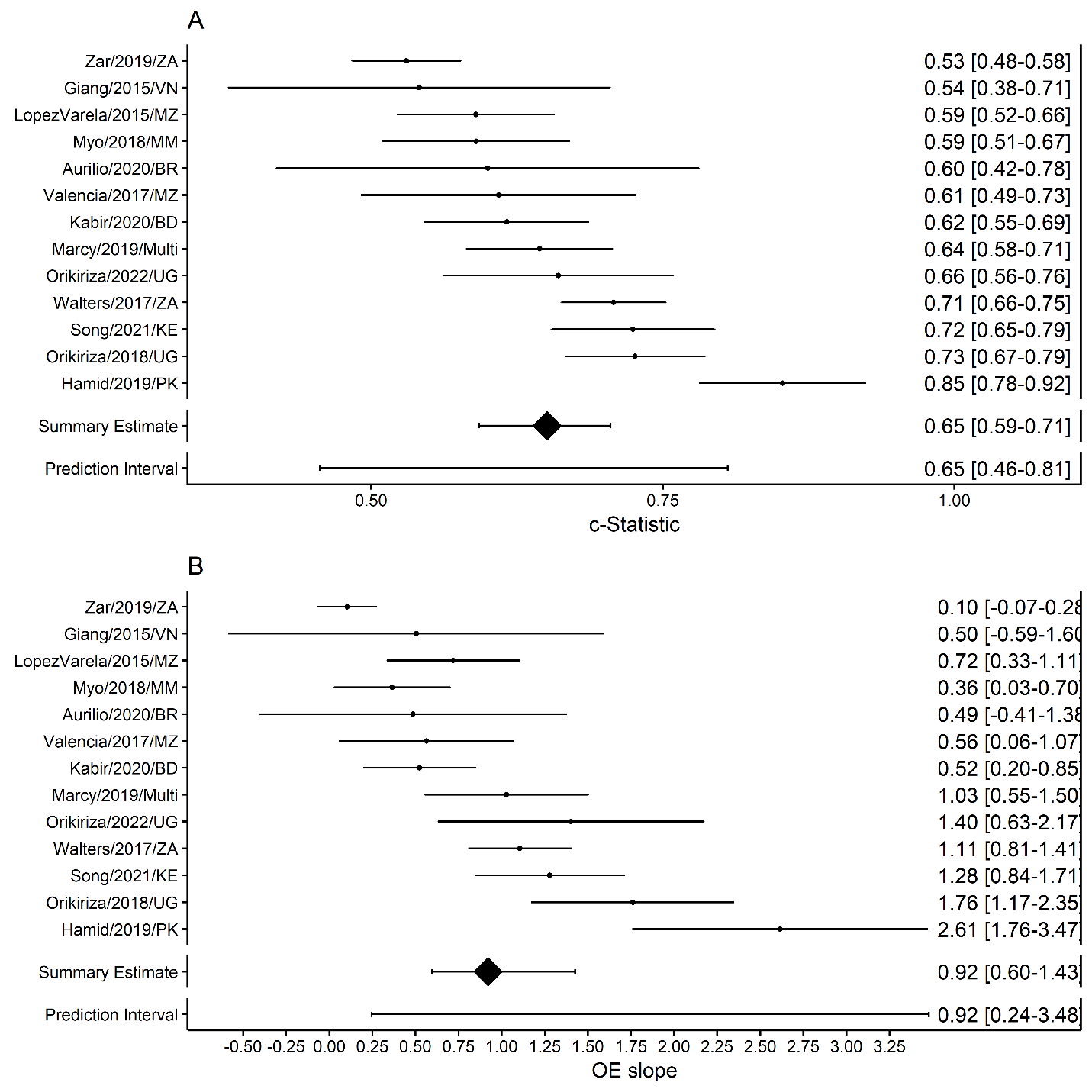

### Appendix R: Prediction model fit and scaled scores at different sensitivity thresholds

**Table S51. OR and 95% CI from prediction model developed from IPD and corresponding scaled scores.**

| **Predictors** | **Coefficient** | **Coefficient 95% CI** | **OR** | **OR 95% CI** | **P-value*** | **Score at 90% sens.** | **Score at 85% sens.** | **Score at 80% sens.** | **Score at 75% sens.** | **Score at 70% sens.** |
| --- | --- | --- | --- | --- | --- | --- | --- | --- | --- | --- |
| Intercept | -1·92 | -2·58, -1·25 | -- | -- | -- | -- | -- | -- | -- | -- |
| Cough duration ≥ 2 weeks  (Absence is no cough or cough <2 weeks) | 0·17 | -0·09, 0·43 | 1·18 | 0·91, 1·54 | 0·86 | 3 | 2 | 2 | 1 | 1 |
| Fever duration ≥ 2 weeks  (Absence is no fever or fever <2 weeks) | 0·45 | 0·16, 0·74 | 1·57 | 1·18, 2·09 | 0·25 | 7 | 5 | 4 | 4 | 3 |
| Lethargy | 0·25 | 0·02, 0·48 | 1·28 | 1·02, 1·62 | 0·66 | 4 | 3 | 2 | 2 | 2 |
| Weight loss | 0·22 | -0·03, 0·48 | 1·25 | 0·97, 1·61 | 0·75 | 3 | 3 | 2 | 2 | 2 |
| History of documented TB exposure | 1·43 | 0·87, 2 | 4·19 | 2·38, 7·38 | <0·001 | 22 | 17 | 14 | 12 | 10 |
| Haemoptysis | 0·34 | -0·37, 1·05 | 1·4 | 0·69, 2·86 | 0·79 | 5 | 4 | 3 | 3 | 2 |
| Night sweats | 0·2 | 0·02, 0·38 | 1·22 | 1·02, 1·47 | 0·71 | 3 | 2 | 2 | 2 | 1 |
| Peripheral lymphadenopathy | 0·35 | 0·13, 0·57 | 1·42 | 1·14, 1·77 | 0·35 | 5 | 4 | 3 | 3 | 2 |
| Temperature ≥38 degrees Celsius | 0 | -0·25, 0·26 | 1 | 0·78, 1·3 | >0·999 | 0 | 0 | 0 | 0 | 0 |
| Tachycardia | 0·15 | -0·13, 0·42 | 1·16 | 0·88, 1·53 | 0·90 | 2 | 2 | 1 | 1 | 1 |
| Tachypnoea | -0·05 | -0·27, 0·16 | 0·95 | 0·77, 1·18 | 0·98 | -1 | -1 | -1 | 0 | 0 |
| Cavities on CXR | 0·47 | -0·11, 1·05 | 1·6 | 0·9, 2·85 | 0·53 | 7 | 6 | 5 | 4 | 3 |
| Intrathoracic lymphadenopathy on CXR | 1·46 | 1, 1·92 | 4·32 | 2·73, 6·85 | <0·001 | 23 | 17 | 14 | 12 | 10 |
| Opacities on CXR | 0·43 | 0·02, 0·84 | 1·54 | 1·02, 2·32 | 0·45 | 7 | 5 | 4 | 4 | 3 |
| Miliary infiltrate on CXR | 1·27 | 0·57, 1·97 | 3·56 | 1·76, 7·19 | <0·001 | 20 | 15 | 12 | 10 | 9 |
| Pleural effusion on CXR | 0·64 | 0·2, 1·09 | 1·9 | 1·22, 2·96 | 0·13 | 10 | 8 | 6 | 5 | 4 |

**Table S52. OR and 95% CI from prediction model without chest x-ray features developed from IPD and corresponding scaled scores.**

| **Predictors** | **Coefficient** | **Coefficient 95% CI** | **OR** | **OR 95% CI** | **P-value*** | **Score at 90% sens.** | **Score at 85% sens.** | **Score at 80% sens.** | **Score at 75% sens.** | **Score at 70% sens.** |
| --- | --- | --- | --- | --- | --- | --- | --- | --- | --- | --- |
| Intercept | -1·36 | -1·94, -0·78 | -- | -- | -- | -- | -- | -- | -- | -- |
| Cough duration ≥ 2 weeks  (Absence is no cough or cough <2 weeks) | 0·22 | -0·03, 0·47 | 1·25 | 0·97, 1·6 | 0·75 | 6 | 5 | 4 | 3 | 3 |
| Fever duration ≥ 2 weeks  (Absence is no fever or fever <2 weeks) | 0·46 | 0·18, 0·73 | 1·58 | 1·2, 2·07 | 0·21 | 13 | 10 | 8 | 7 | 6 |
| Lethargy | 0·2 | -0·01, 0·41 | 1·22 | 0·99, 1·51 | 0·75 | 6 | 4 | 4 | 3 | 3 |
| Weight loss | 0·24 | 0·01, 0·48 | 1·28 | 1·01, 1·62 | 0·68 | 7 | 5 | 4 | 4 | 3 |
| History of documented TB exposure | 1·33 | 0·81, 1·84 | 3·76 | 2·24, 6·31 | <0·001 | 39 | 29 | 24 | 20 | 17 |
| Haemoptysis | 0·4 | -0·27, 1·06 | 1·49 | 0·76, 2·89 | 0·70 | 12 | 9 | 7 | 6 | 5 |
| Night sweats | 0·28 | 0·12, 0·45 | 1·33 | 1·12, 1·57 | 0·43 | 8 | 6 | 5 | 4 | 4 |
| Peripheral lymphadenopathy | 0·32 | 0·12, 0·52 | 1·38 | 1·13, 1·68 | 0·40 | 9 | 7 | 6 | 5 | 4 |
| Temperature ≥38 degrees Celsius | 0·01 | -0·22, 0·23 | 1·01 | 0·8, 1·26 | >0·999 | 0 | 0 | 0 | 0 | 0 |
| Tachycardia | 0·19 | -0·08, 0·47 | 1·21 | 0·92, 1·6 | 0·83 | 6 | 4 | 3 | 3 | 2 |
| Tachypnoea | 0·07 | -0·18, 0·33 | 1·08 | 0·84, 1·39 | 0·97 | 2 | 2 | 1 | 1 | 1 |

### Appendix S: Performance of scores from prediction model at different sensitivity thresholds

**Figure S21. (A) sensitivity and (B) specificity of score developed from prediction model to classify TB with 90% sensitivity.**

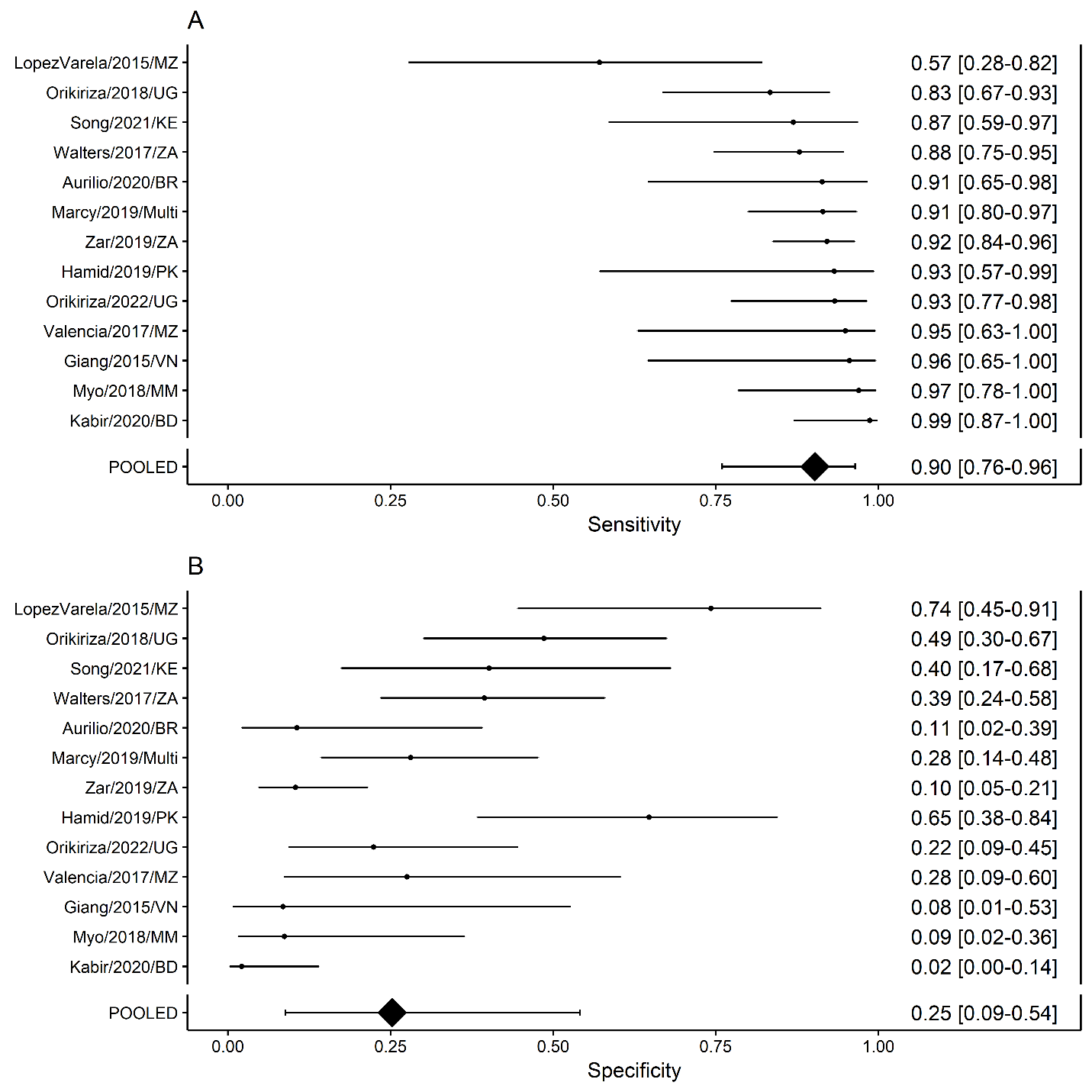

**Figure S22. (A) sensitivity and (B) specificity of score developed from prediction model to classify TB with 85% sensitivity.** Presented in the main text.

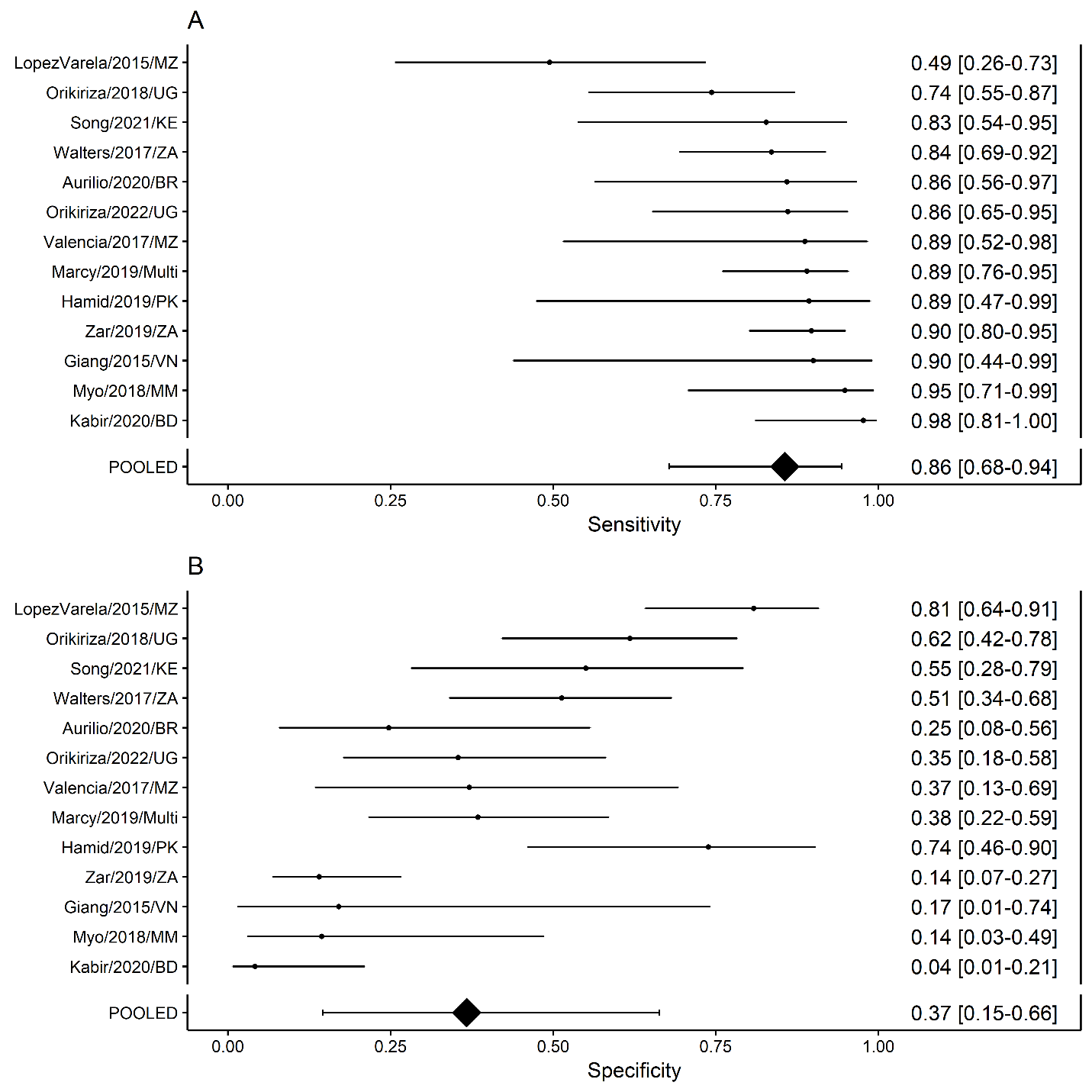

**Figure S23. (A) sensitivity and (B) specificity of score developed from prediction model to classify TB with 80% sensitivity.**

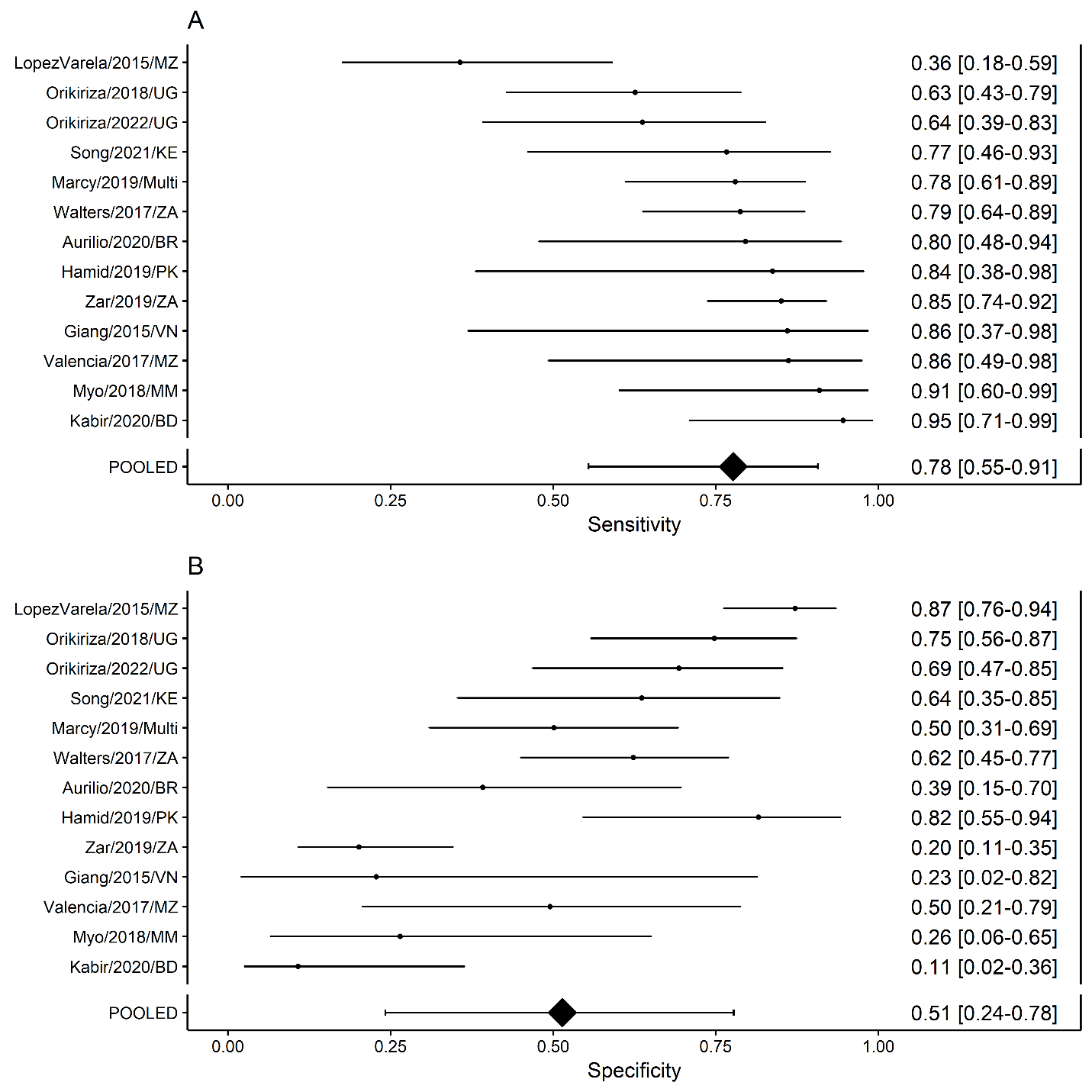

**Figure S24. (A) sensitivity and (B) specificity of score developed from prediction model to classify TB with 75% sensitivity.**

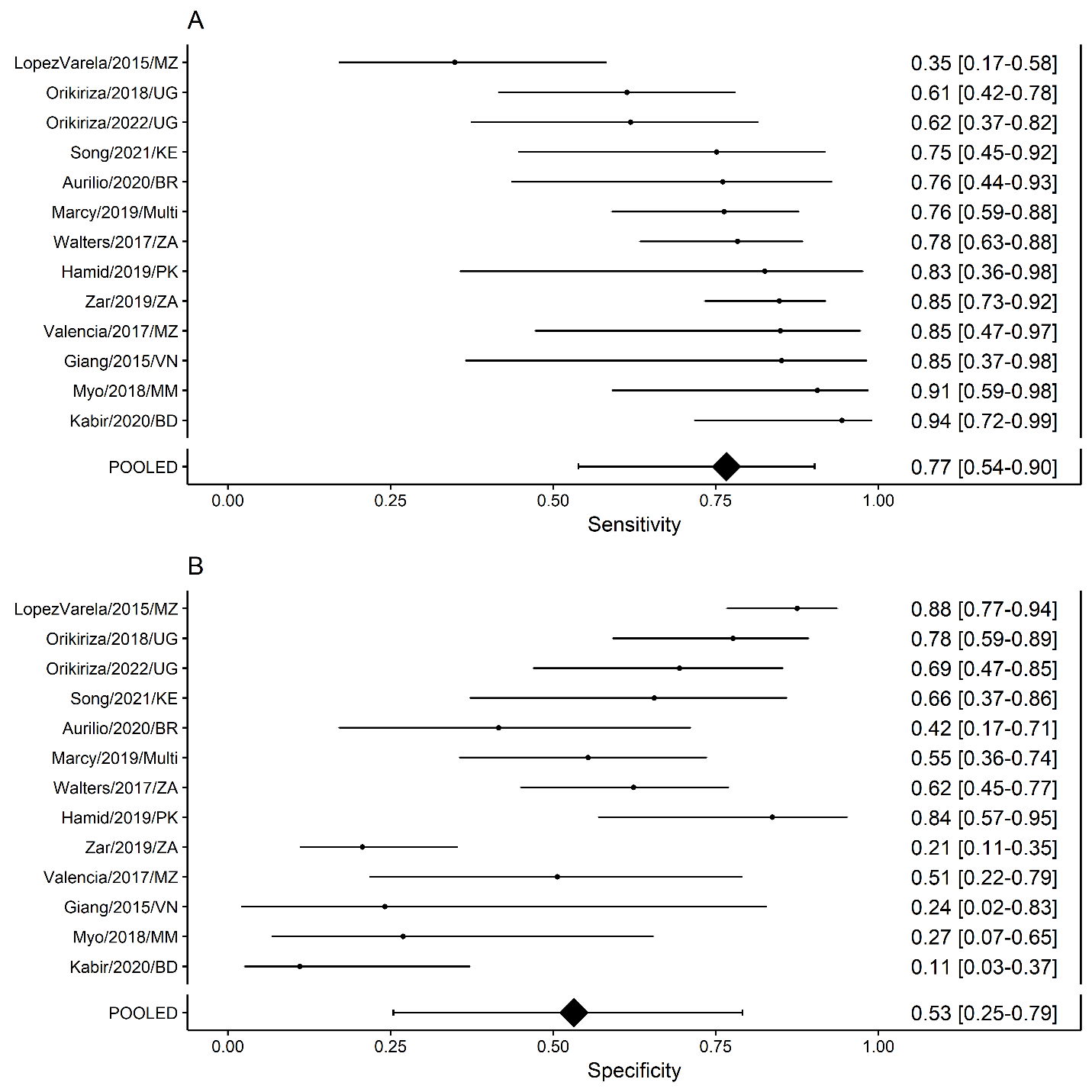

**Figure S25. (A) sensitivity and (B) specificity of score developed from prediction model to classify TB with 70% sensitivity.**

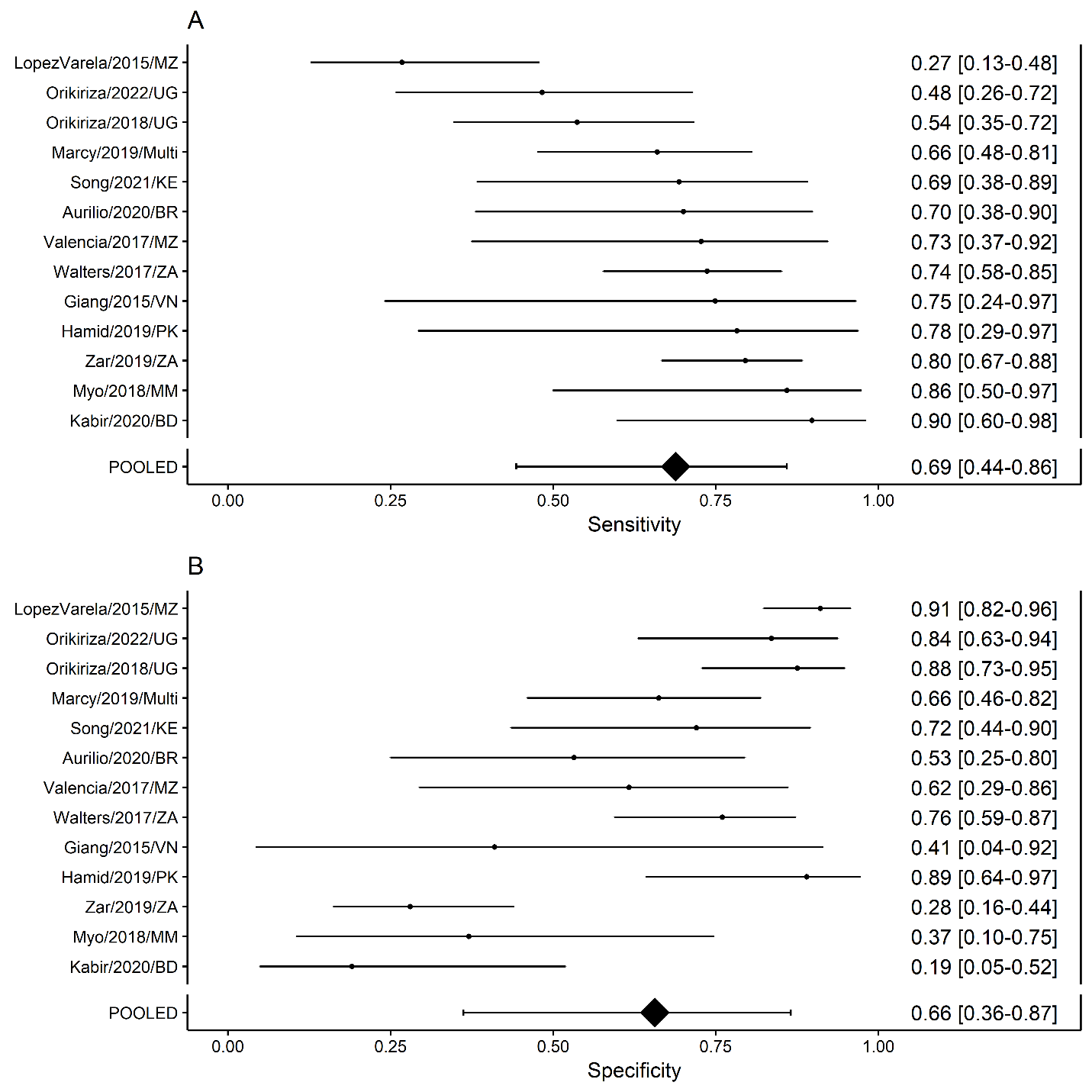

### Appendix T: Performance of scores to classify TB with 85% sensitivity using confirmed TB reference standard

**Figure S26.** **(A) sensitivity and (B) specificity of score developed from prediction model to classify bacteriologically-confirmed TB with 85% sensitivity.** Analysis excludes data from children with unconfirmed TB.

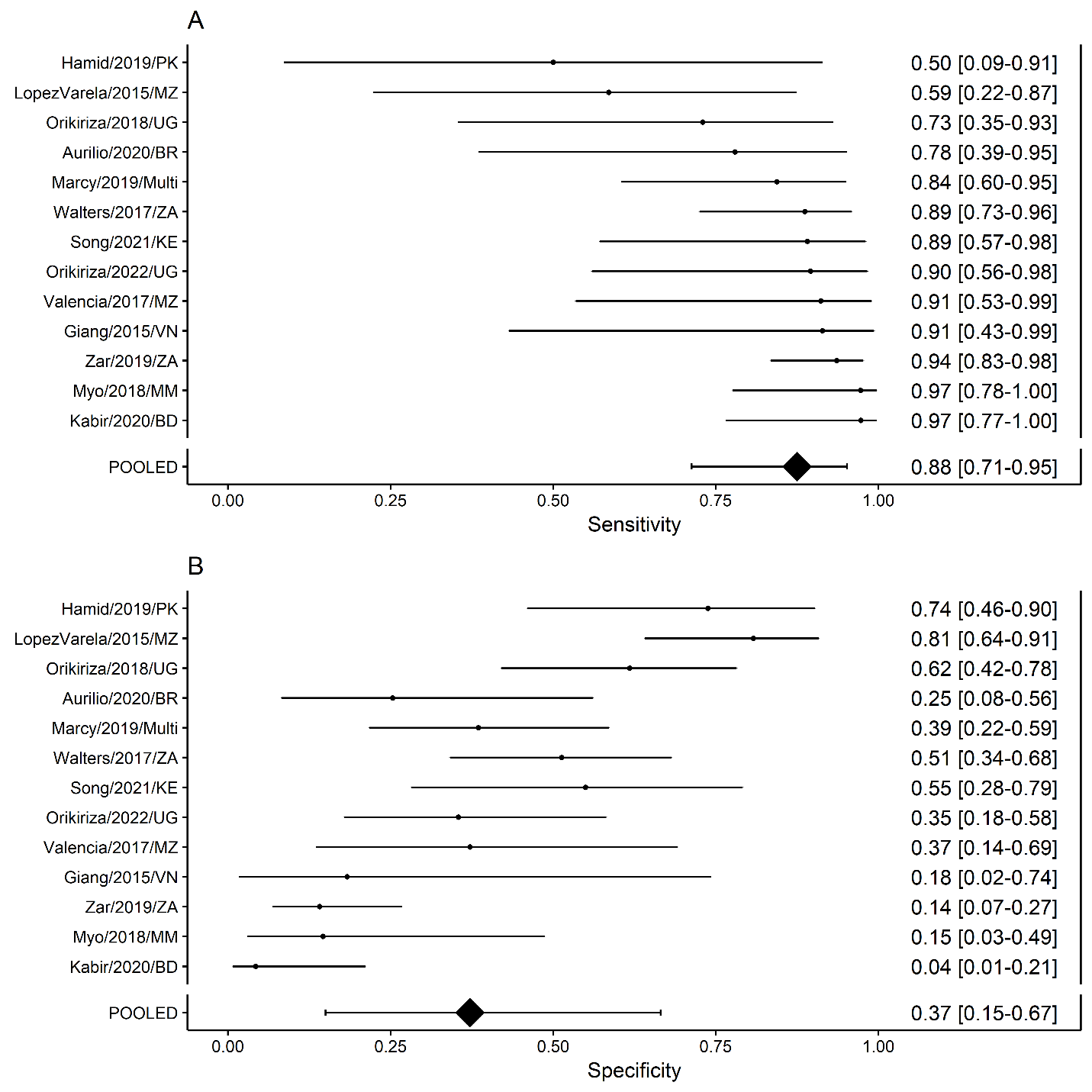

### Appendix U: Performance of scores to classify TB with 85% sensitivity (without chest X-ray features)

**Figure S27. (A) sensitivity and (B) specificity of score developed from prediction model without chest x-ray to classify TB using composite reference standard with 85% sensitivity.**

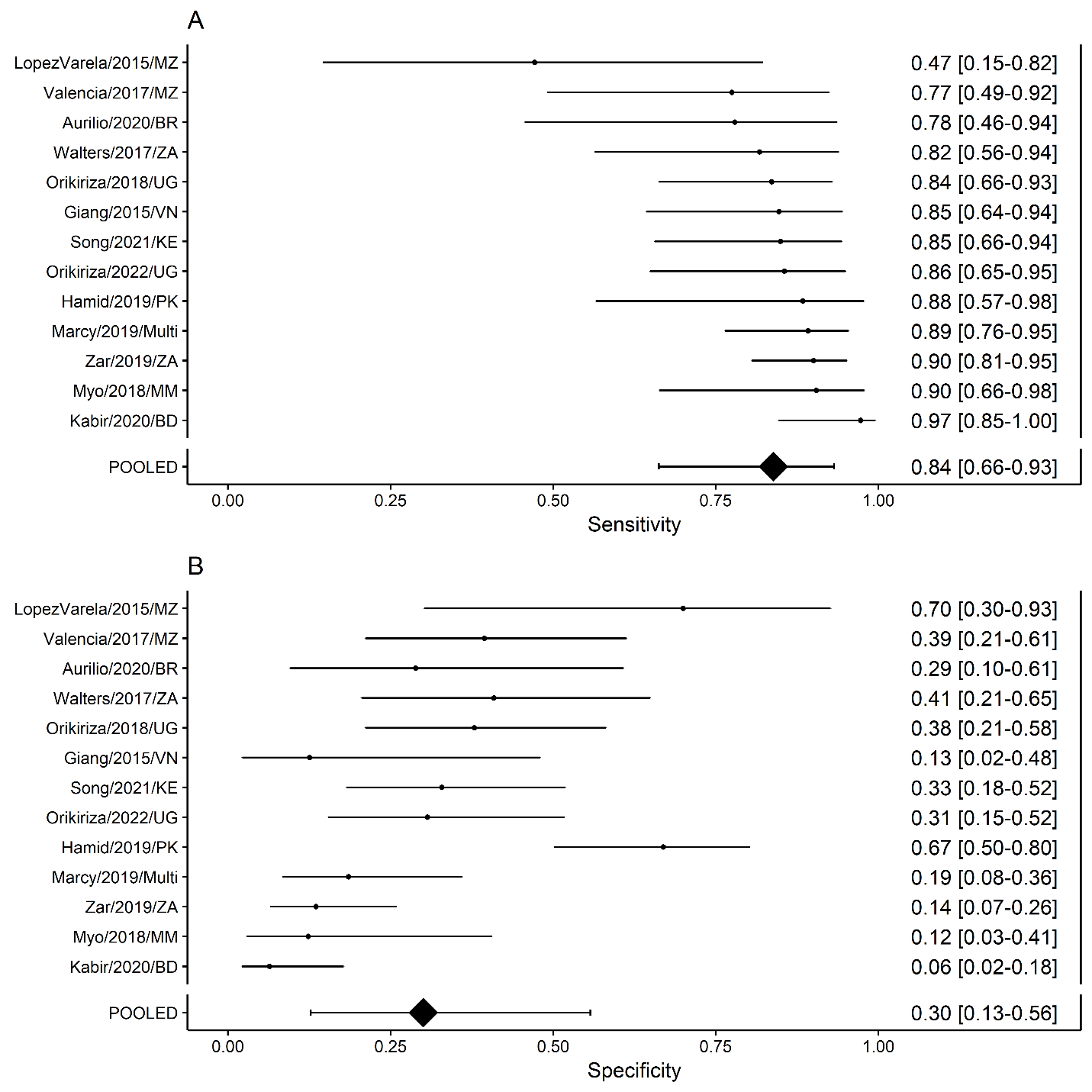

**Figure S28.** **(A) sensitivity and (B) specificity of score developed from prediction model without chest x-ray to classify bacteriologically-confirmed TB with 85% sensitivity.** Analysis excludes data from children with unconfirmed TB.

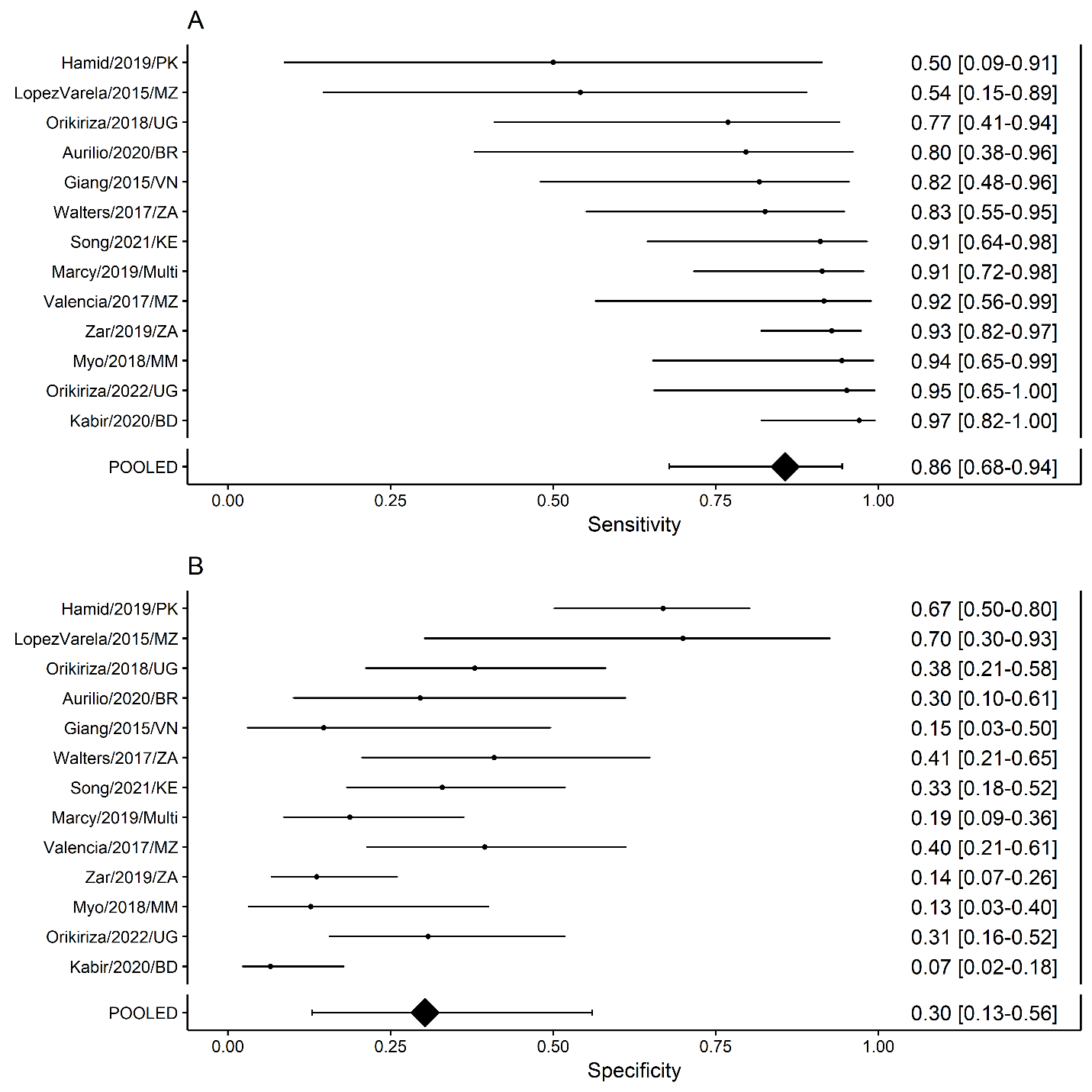

### Appendix V: Inspire algorithm without CXR features

**Figure S29. Treatment-decision algorithm derived from prediction model without CXR features.** Scores associated with features from clinical history and physical exam and chest X-ray translate to risk of TB and are scaled from the prediction model developed from the IPD. Guidance on the practical use of this algorithm is outlined in the WHO operational handbook.^29^ WHO – World Health Organization, TB – tuberculosis, IPD – individual participant data, HIV – human immunodeficiency virus, mWRD – molecular WHO-recommended rapid diagnostic test, CLHIV – children living with HIV, LF-LAM – lateral flow urine lipoarabinomannan assay, CXR – chest X-ray.

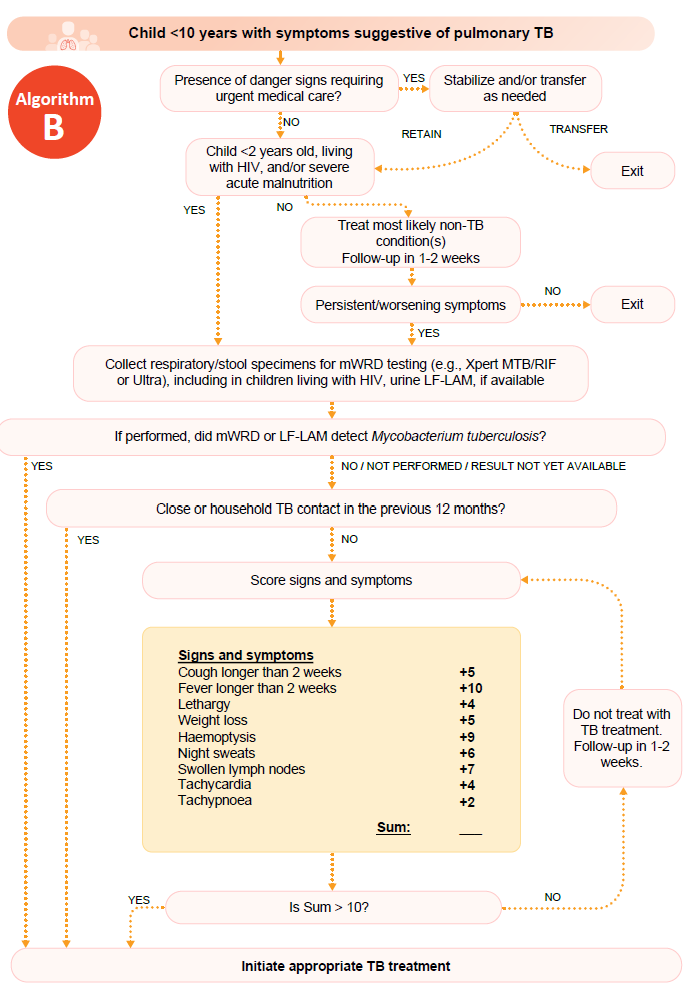

### Appendix W: TRIPOD Checklist

**Table S53. TRIPOD Checklist^30^** Page numbers preceded with an “S” refer to the pages within the supplement.

| Section/Topic | **Item** |  | **Checklist Item** | **Page** |
| --- | --- | --- | --- | --- |
| Title and abstract | | | | |
| Title | **1** | **D;V** | **Identify the study as developing and/or validating a multivariable prediction model, the target population, and the outcome to be predicted.** | Title page |
| Abstract | **2** | **D;V** | **Provide a summary of objectives, study design, setting, participants, sample size, predictors, outcome, statistical analysis, results, and conclusions.** | 1 |
| Introduction | | | | |
| Background and objectives | **3a** | **D;V** | **Explain the medical context (including whether diagnostic or prognostic) and rationale for developing or validating the multivariable prediction model, including references to existing models.** | 5-6 |
|  | **3b** | **D;V** | **Specify the objectives, including whether the study describes the development or validation of the model or both.** | 6 |
| Methods | | | | |
| Source of data | **4a** | **D;V** | **Describe the study design or source of data (e.g., randomized trial, cohort, or registry data), separately for the development and validation data sets, if applicable.** | 6-7, S12-S65 |
|  | **4b** | **D;V** | **Specify the key study dates, including start of accrual; end of accrual; and, if applicable, end of follow-up.** | S12-S24 |
| Participants | **5a** | **D;V** | **Specify key elements of the study setting (e.g., primary care, secondary care, general population) including number and location of centres.** | 6-7, S12-S24 |
|  | **5b** | **D;V** | **Describe eligibility criteria for participants.** | 6-7, S12-S24 |
|  | **5c** | **D;V** | **Give details of treatments received, if relevant.** | NA |
| Outcome | **6a** | **D;V** | **Clearly define the outcome that is predicted by the prediction model, including how and when assessed.** | 6-9, S12-S24 |
|  | **6b** | **D;V** | **Report any actions to blind assessment of the outcome to be predicted.** | S12-S24 |
| Predictors | **7a** | **D;V** | **Clearly define all predictors used in developing or validating the multivariable prediction model, including how and when they were measured.** | S3-S6, S12-S37, S65 |
|  | **7b** | **D;V** | **Report any actions to blind assessment of predictors for the outcome and other predictors.** | NA |
| Sample size | **8** | **D;V** | **Explain how the study size was arrived at.** | NA |
| Missing data | **9** | **D;V** | **Describe how missing data were handled (e.g., complete-case analysis, single imputation, multiple imputation) with details of any imputation method.** | 7, S7-S8 |
| Statistical analysis methods | **10a** | **D** | **Describe how predictors were handled in the analyses.** | 7-8 |
|  | **10b** | **D** | **Specify type of model, all model-building procedures (including any predictor selection), and method for internal validation.** | 7-8 |
|  | **10c** | **V** | **For validation, describe how the predictions were calculated.** | NA |
|  | **10d** | **D;V** | **Specify all measures used to assess model performance and, if relevant, to compare multiple models.** | 7-8 |
|  | **10e** | **V** | **Describe any model updating (e.g., recalibration) arising from the validation, if done.** | NA |
| Risk groups | **11** | **D;V** | **Provide details on how risk groups were created, if done.** | NA |
| Development vs. validation | **12** | **V** | **For validation, identify any differences from the development data in setting, eligibility criteria, outcome, and predictors.** | NA |
| Results | | | | |
| Participants | **13a** | **D;V** | **Describe the flow of participants through the study, including the number of participants with and without the outcome and, if applicable, a summary of the follow-up time. A diagram may be helpful.** | 9-10, S12-S24 |
|  | **13b** | **D;V** | **Describe the characteristics of the participants (basic demographics, clinical features, available predictors), including the number of participants with missing data for predictors and outcome.** | 9-10, S38-S63 |
|  | **13c** | **V** | **For validation, show a comparison with the development data of the distribution of important variables (demographics, predictors and outcome).** | NA |
| Model development | **14a** | **D** | **Specify the number of participants and outcome events in each analysis.** | 9-10, 23-24 |
|  | **14b** | **D** | **If done, report the unadjusted association between each candidate predictor and outcome.** | NA |
| Model specification | **15a** | **D** | **Present the full prediction model to allow predictions for individuals (i.e., all regression coefficients, and model intercept or baseline survival at a given time point).** | 25 |
|  | **15b** | **D** | **Explain how to the use the prediction model.** | 10-11 |
| Model performance | **16** | **D;V** | **Report performance measures (with CIs) for the prediction model.** | 10-11, 25 |
| Model-updating | **17** | **V** | **If done, report the results from any model updating (i.e., model specification, model performance).** | NA |
| Discussion | | | | |
| Limitations | **18** | **D;V** | **Discuss any limitations of the study (such as nonrepresentative sample, few events per predictor, missing data).** | 14-15 |
| Interpretation | **19a** | **V** | **For validation, discuss the results with reference to performance in the development data, and any other validation data.** | NA |
|  | **19b** | **D;V** | **Give an overall interpretation of the results, considering objectives, limitations, results from similar studies, and other relevant evidence.** | 12-16 |
| Implications | **20** | **D;V** | **Discuss the potential clinical use of the model and implications for future research.** | 16 |
| Other information | | | | |
| Supplementary information | **21** | **D;V** | **Provide information about the availability of supplementary resources, such as study protocol, Web calculator, and data sets.** | 21 |
| Funding | **22** | **D;V** | **Give the source of funding and the role of the funders for the present study.** | 19-21, S12-S24 |

### Appendix X: Supplementary references

1.	van Buuren S, Groothuis-Oudshoorn K. mice: Multivariate imputation by chained equations in R. *J Stat Softw* 2011; **45**(3): 1-67.

2.	Robitzsch A, Grund S. miceadds: some additional multiple imputation functions, especially for 'mice'. R package version 3.13-12, <https://CRAN.R-project.org/package=miceadds>. 2022.

3.	Reitsma JB, Glas AS, Rutjes AWS, Scholten RJPM, Bossuyt PM, Zwinderman AH. Bivariate analysis of sensitivity and specificity produces informative summary measures in diagnostic reviews. *J Clin Epidemiol* 2005; **58**(10): 982-90.

4.	Heymans MW, Eekhout I. Applied missing data analysis with SPSS and (R)Studio; 2019.

21.	Wells GA, Shea B, O'Connell D, et al. The Newcastle-Ottawa Scale (NOS) for assessing the quality of nonrandomised studies in meta-analyses <https://www.ohri.ca/programs/clinical_epidemiology/oxford.asp>.

22.	Marais BJ, Gie RP, Hesseling AC, et al. A refined symptom-based approach to diagnose pulmonary tuberculosis in children. *Pediatrics* 2006; **118**(5): e1350-9.

23.	Graham S. The Union's desk guide for diagnosis and management of TB in children. 3 ed. Paris, France: International Union Against Tuberculosis and Lung Disease; 2016.

24.	Montenegro SH, Gilman RH, Sheen P, et al. Improved detection of Mycobacterium tuberculosis in Peruvian children by use of a heminested IS6110 polymerase chain reaction assay. *Clin Infect Dis* 2003; **36**(1): 16-23.

25.	Zawedde-Muyanja S, Nakanwagi A, Dongo JP, et al. Decentralisation of child tuberculosis services increases case finding and uptake of preventive therapy in Uganda. *Int J Tuberc Lung Dis* 2018; **22**(11): 1314-21.

26.	Brasil. Ministério da Saúde. Secretaria de Vigilância em Saúde. Departamento de Vigilância das Doenças Transmissíveis. Manual de recomendações para o controle da tuberculose no Brasil. 2. ed: Brasília: Ministério da Saúde; 2019.

27.	Edwards K. The diagnosis of childhood tuberculosis. *P N G Med J* 1987; **30**(2): 169-78.

28.	Gunasekera KS, Walters E, van der Zalm MM, et al. Development of a treatment-decision algorithm for human immunodeficiency virus-uninfected children evaluated for pulmonary tuberculosis. *Clin Infect Dis* 2021; **73**(4): e904-e12.

29.	World Health Organization. WHO operational handbook on tuberculosis. Module 5: management of tuberculosis in children and adolescents. Geneva: World Health Organization; 2022.

30.	Collins GS, Reitsma JB, Altman DG, Moons KGM. Transparent reporting of a multivariable prediction model for individual prognosis or diagnosis (TRIPOD): the TRIPOD statement. *Br Med J* 2015; **350**: g7594.
